# Emulating a vaccine target trial under unmeasured confounding: single and double negative control methods

**DOI:** 10.64898/2026.10.21.26366144

**Authors:** Beau Schaeffer, Tyler Chervo, Eric P. Elkin, William P. Hanage, Joshua R. Nugent, Ilan N. Rubin, Joy Shi, Jacek Skarbinski, Christopher B. Boyer

## Abstract

Estimating vaccine effectiveness (VE) for symptom-driven outcomes in observational settings remains challenging due to unmeasured differences in health-seeking behavior between vaccine groups. This challenge persists even with target trial emulation, which primarily addresses design-related biases. The test negative design, a popular alternative design for estimating VE, can reduce bias due to unmeasured confounding by health-seeking but often deviates from the design principles of trial emulation. Here, we emulate a target trial to estimate relative VE of COVID-19 bivalent booster in a cohort of Kaiser Permanente of Northern California members and provide evidence for residual unmeasured confounding through negative controls. We then propose two approaches for estimating VE that operate within the target trial framework but allow for unmeasured confounding. The first uses test-negative illness as a negative control outcome and corrects for bias under the assumption of equi-confounding on the multiplicative scale. The second relaxes equi-confounding but requires an additional (valid) negative control. The two approaches yield similar estimates of 1-year relative VE (11.1% vs. 15.9%) but are both qualitatively different from traditional methods (VE: *−*14.5%). These approaches illustrate the flexibility of the target trial framework which can retain sound design principles while accommodating alternative identification strategies.

## Introduction

Vaccine effectiveness (VE) studies increasingly use target trial emulation (TTE), a popular framework for comparative effectiveness analyses of observational data, when randomized trials are not feasible [1, 2]. TTE resolves biases due to inappropriate study design (e.g., those involving selection and immortal time), but does not necessarily address issues arising from data limitations, such as insufficient information on sources of confounding [3]. This is of particular concern when evaluating VE, because health-seeking behavior, which is often unmeasured or hard to fully characterize, typically varies between treatment groups. Health-seeking behaviors can affect both underlying infection risk, through associated behaviors like masking or hand-washing, as well as ascertainment, through the decision to pursue testing services when sick. These behaviors may also vary over time, as testing guidance, supply, or other factors such as perceived risk change over the course of a respiratory illness season. The difficulty in fully characterizing these behaviors thus leads to a pernicious source of residual confounding in observational studies of VE that is not resolvable by TTE alone.

The test negative study design (TND) is a popular alternative design for evaluation of VE. The TND addresses unmeasured confounding by health-seeking behavior by using additional information from individuals who presented for testing with similar symptoms, but tested negative for the focal pathogen targeted by vaccination. If the vaccine has no effect on this “test-negative illness”, then any residual differences in testing negative by vaccination status, after accounting for measured covariates and sampling variability, likely reflect residual unmeasured confounding [4, 5]. In this case, testing negative functions as a negative control outcome (NCO), which can be used to de-bias estimates of VE, provided certain conditions hold [6]. Under outcome dependent sampling among the tested, the TND odds ratio recovers VE in the underlying cohort.

However, the TND also has several notable limitations. Because the TND does not explicitly emulate a target trial (or often deviates from the target trial it is implicitly emulating), it may be affected by design-related biases [7] (for a counterexample of a TND explicitly embedded within an RCT see, for example, Jewell, Dufault, Cutcher, Simmons, and Anders [8]). When vaccination occurs over the course of a respiratory illness season, one design-related bias is the potential for immortal time due to an unclear “time zero” among the unvaccinated (i.e., misclassification) [9]. Another design-related bias is the potential for immortal time due to selection as eligibility in the TND may be assessed based on data collected at time of testing rather than at an unambiguous time zero [9]. Finally, because data on the full underlying cohort is typically not available, the TND yields only relative effect estimates rather than absolute effects that may be important for risk communication and public health decision-making.

To overcome these limitations, we propose two bias correction approaches for VE studies that operate within the target trial framework but relax the assumption of no unmeasured confounding. The first approach (abbreviated throughout as EQC) uses test-negative information as an NCO to de-bias VE estimates as in the TND, but embedded within a full cohort rather than confined to data from tested individuals only. This allows for explicit emulation of a target trial, and therefore mitigation of design-related biases, prior to utilization of test-negative information. Under “equi-confounding” [6], the resulting difference-in-differences estimator is related to the OR estimator in the TND but also allows for recovery of absolute effect measures such as risk/cumulative incidence differences in the full cohort. The EQC approach accounts for unmeasured health-seeking behavior (a canonical source of equi-confounding) but can also encompass other sources of unmeasured confounding provided they affect test-negative and test-positive illness equivalently on the multiplicative scale.

The second approach, which we refer to as the proximal causal inference approach (hereafter, abbreviated as PCI), builds on the EQC approach embedded within a TTE but identifies an additional variable, a negative control exposure (NCE), to further relax the unmeasured confounding assumption [10, 11]. The NCE must be a priori assumed to have no direct causal effect on the outcome, but related to treatment through the unmeasured confounder. In our emulation of a COVID-19 bivalent booster trial below, we use prior year vaccination for a different pathogen (influenza) as an NCE. Assuming access to a valid NCE as well as the validity of testing negative as an NCO, the PCI approach recovers VE under less restrictive (and therefore more general) unmeasured confounding using a two-step estimation procedure.

In what follows, as a motivating example, we first specify and emulate a target trial in a cohort of previously vaccinated Kaiser Permanente Northern California members to estimate the relative VE for the mRNA COVID-19 bivalent booster vaccine (BA.4/5 formulation). We demonstrate potential failures of a standard matching and standardization approach that assumes no unmeasured confounding. We then discuss and emulate trials under the EQC and PCI strategies described above. We contrast these three approaches against a TND to illustrate how concerns of unmeasured confounding by health seeking behavior may or may not be mitigated across these different approaches.

## Emulating a Trial for COVID-19 Bivalent Booster

### Specification of the Target Trial

We designed this observational analysis to emulate a target trial of bivalent booster vaccination versus no bivalent booster vaccination among those with up-to-date vaccination history on medically attended COVID-19.We first specify the protocol of this target trial, and then describe how we emulate the target trial using observational data.

Full details of the target trial protocol are found in Table 2. In brief, target trial enrollment period was from 09-01-2022 to 11-30-2022. Individuals were eligible if they had at least 11 months of prior Kaiser Permanente Northern California (KPNC) membership, had a primary series of 2 COVID-19 vaccinations and one booster, all prior COVID-19 vaccinations were either Pfizer or Moderna (the predominant vaccinations in use at KPNC), had at least one prior COVID-19 lab test in 2022, did not have a positive nucleic acid amplification test (NAAT) or COVID-19 diagnosis code in the prior 90 days, and were 18 years of age or older. Eligible individuals would be randomized to receive one of two treatment strategies: (1) bivalent booster at baseline or (2) no booster vaccination at baseline. The outcome of interest would be medically attended COVID-19 confirmed by a positive polymerase-chain-reaction (PCR) test for SARS-CoV-2. Individuals would be followed from treatment assignment until development of the outcome event, end of follow-up at 52 weeks, disenrollment from the health plan, or death, whichever comes first. We present per-protocol (PP) effects in our main analysis, censoring individuals at deviations from their assigned treatment strategy (any vaccination for individuals assigned to the no booster arm and any additional vaccination for individuals assigned to the booster arm) and retain intention-to-treat (ITT) effects in Appendix S2.3. Risks were obtained using a standard Kaplan-Meier estimator and used to construct weekly risk differences (RD) and risk ratios (RR).

### Emulation of the Target Trial

We emulated the target trial described above using electronic health records from KPNC. KPNC is a health system that serves over 4.5 million members in Northern and Central California providing outpatient and inpatient care across 266 medical offices and 21 hospitals. Members receive most of their clinical services, including laboratory testing, outpatient, and inpatient care in KPNC facilities [12]. For treatment assignment, individuals were classified as treated if they received a bivalent booster during a given week during the enrollment period, and had not been previously matched or developed the study outcome as of that week. For each treated individual, another individual was 1:1 matched and classified as untreated if they had not previously received a bivalent booster, not been previously matched, and did not have a COVID-19 infection as of that week. Exact matching was performed on sex, race, KPNC service area, receipt of any prior COVID-19 infection, and Charlson comorbidity index; nearest neighbor matching was performed on age, neighborhood deprivation index, BMI, total COVID-19 test count, and weeks since most recent COVID-19 infection or vaccination. Receipt of prior year influenza vaccination, our candidate NCE, was not included as a matching factor but does appear as a covariate in our models. From treatment assignment, individuals were followed until treatment deviation (PP analysis only), development of the event, or end of follow-up at 52 weeks, whichever came first (Figure 1).

**Figure 1:**
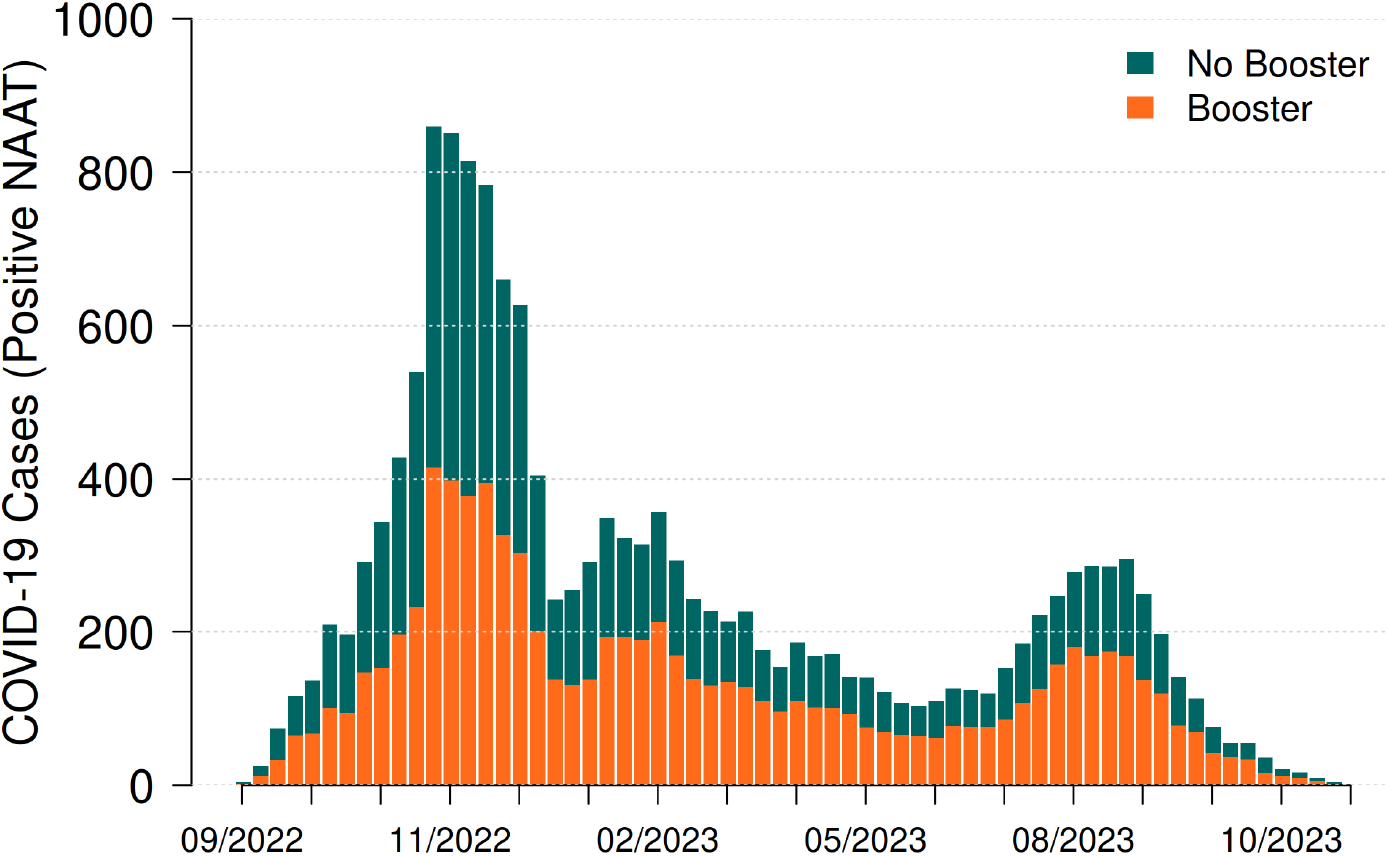
COVID-19 cases among eligible KPNC members over the study period.

### Confounding Control via Measured Covariate Adjustment

We first attempted to estimate VE using a conventional measured covariate adjustment approach (specifically matching followed by regression adjustment and standardization) which assumes that all sources of confounding between treatment and outcome are measured and appropriately modeled (Figure S9a). This is the most common approach in target trial emulations. In lieu of randomization, this approach assumes conditional exchangeability of treatment arms within levels of the matching factors listed above. Weekly estimates of risk were estimated via parametric g-computation using a pooled logistic regression model and Kaplan-Meier-type algorithm. We included a product term between treatment and time, and modeled time flexibly using natural splines with knot points at 10, 20, 30, 40, and 50 weeks. Nonparametric bootstrapping with 200 replicates was used to calculate weekly percentile-based 95% confidence intervals for risk estimates in each group. A summary hazard ratio was also estimated using a Cox proportional hazards model, with robust standard errors derived from the Lin-Wei sandwich variance estimator to account for matching. VE was calculated in all approaches using (1*−CIR*) for pooled logistic estimates and (1*−HR*) for Cox estimates.

### Evidence for Residual Unmeasured Confounding

The measured covariate adjustment approach yielded a summary HR of 1.14 (1.11, 1.18), indicating an apparent increased hazard of medically attended COVID-19 for the bivalent booster group relative to the no booster group (Table 1, Figure 2B). Pooled logistic results provide a more nuanced picture of risk over the year following vaccination, showing apparent waning of protection after roughly 29 weeks (Figure 2A, Table S11). As evidence for residual unmeasured confounding, we used two negative controls. First, using prior year flu vaccination as an NCE, we note an increased hazard of COVID-19 among individuals who received prior year flu vaccination compared to those who did not (Table 1) after matching and adjustment for measured covariates, implying potential residual excess risk of testing positive among the vaccinated (here, flu). Second, using those who tested negative for SARS-CoV-2 as an NCO, we observe an increased hazard of testing negative among the vaccinated (Figure 2B, see also cumulative incidence curves in Figure S10) after matching and adjustment for measured covariates. Finally, using both negative control variables, we observe an increased hazard of testing negative among individuals who received prior year flu vaccination (Figure 2B). Taken together, these results are compelling evidence that testing behaviors remain imbalanced between the boosted and unboosted populations despite fine-scale adjustment for 11 measured covariates.

**Table 1:** Effectiveness of the bivalent COVID-19 booster against a positive SARS-CoV-2 test over 52 weeks under four analytic approaches, with negative-control diagnostics (per-protocol).

| Approach | Test Positive:<br>Bivalent Booster |  | Test Positive:<br>Prior Year Flu Vax<br>(NCE) |  | Test Negative:<br>Bivalent Booster<br>(NCO) |  | VE (%) | 95% CI |
| --- | --- | --- | --- | --- | --- | --- | --- | --- |
|  | HR | 95% CI | HR | 95% CI | HR | 95% CI |  |  |
| STD | 1.14 | (1.11, 1.18) | 1.26 | (1.22, 1.31) | 1.26 | (1.23, 1.28) | −14.5 | (−18.3, −10.8) |
| TND | 0.90 | (0.87, 0.94) | 1.03 | (0.99, 1.08) | — | — | 9.8 | (6.0, 13.5) |
| EQC | 0.89 | (0.86, 0.92) | 1.04 | (1.01, 1.09) | — | — | 11.1 | (7.6, 14.5) |
| PCI | 0.84 | (0.76, 0.90) | — | — | — | — | 15.9 | (10.2, 23.9) |

*Note.* Entries are hazard ratios (HR) with 95% confidence intervals from Cox proportional hazards models; vaccine effectiveness is $VE = 100 \times (1 - HR)$ computed from the test-positive/booster HR and represents average relative vaccine effectiveness over 52 weeks. Analyses are per-protocol and adjusted for the baseline covariates in Table S8; treated and comparator individuals were matched on calendar week. *Test Positive: Bivalent Booster* is the target association from which VE is derived. *Test Positive: Prior Year Flu Vax (NCE)* is the association between the negative-control exposure (prior-season influenza vaccination) and a positive test, expected to be null ( $HR = 1$ ) absent residual confounding; departures from 1 signal uncontrolled confounding. *Test Negative: Bivalent Booster (NCO)* is the association between the booster and the negative-control outcome (a negative test), expected to be null under a valid NCO. “—” denotes a quantity not separately estimated under that approach because it is used internally for de-biasing: the NCO is the de-biasing proxy in TND and EQC, while the NCO and NCE form the proximal bridge in PCI.
*Abbreviations:* STD, standard covariate-adjusted analysis (no negative-control correction); TND, test-negative design; EQC, equi-confounding; PCI, proximal causal inference; CI, confidence interval; HR, hazard ratio; VE, vaccine effectiveness; NCE, negative control exposure; NCO, negative control outcome.

**Table 2:**
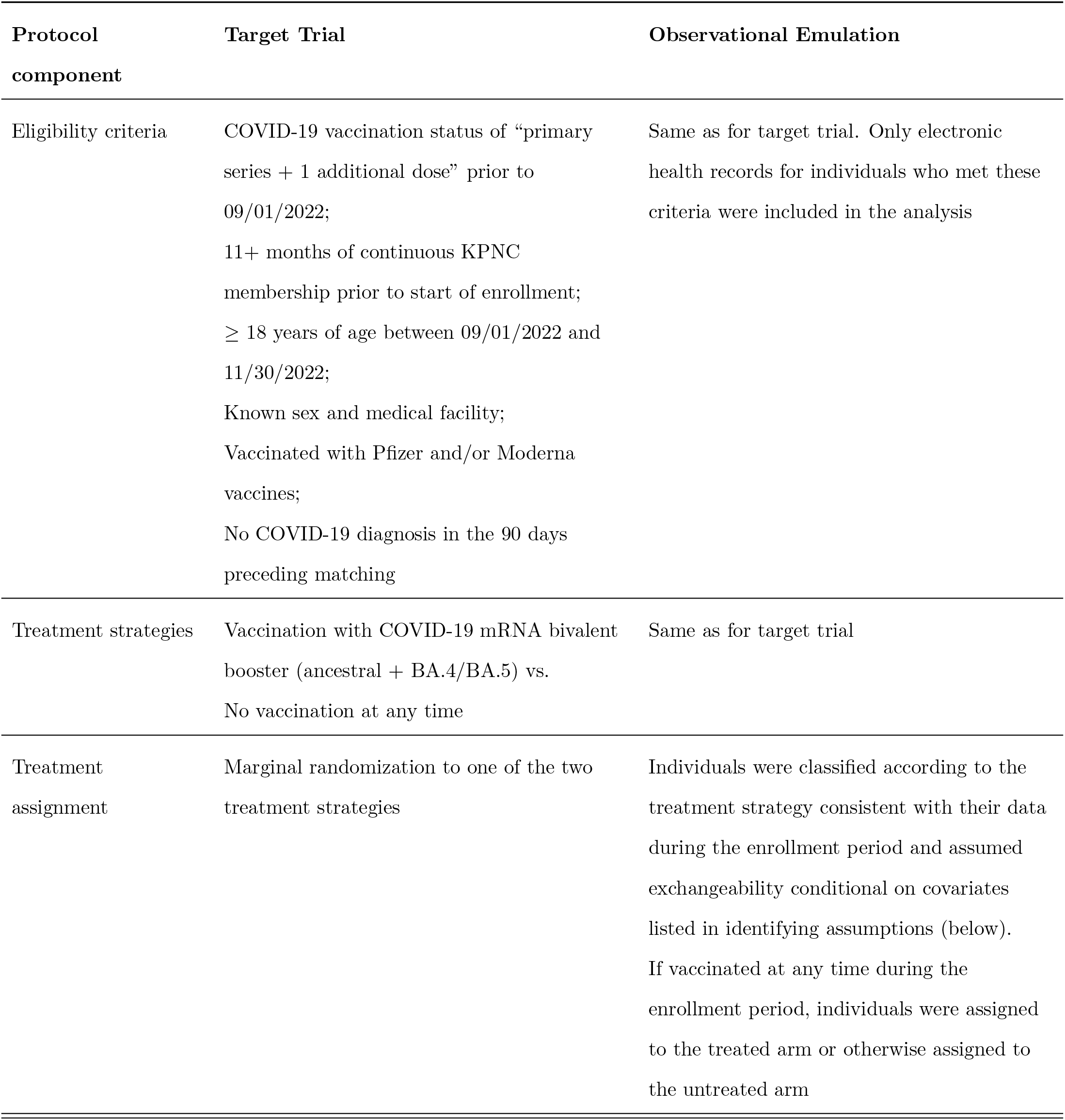
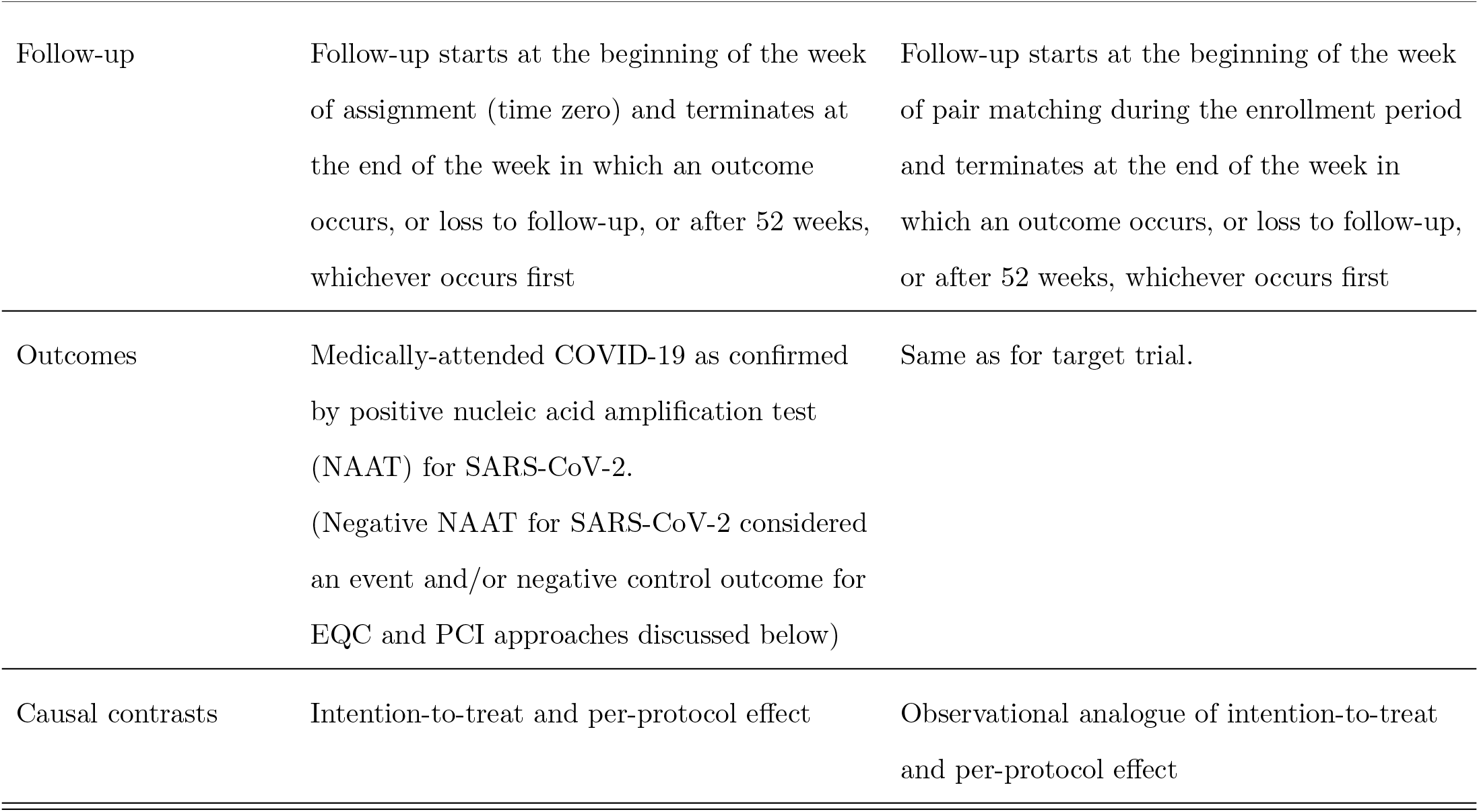
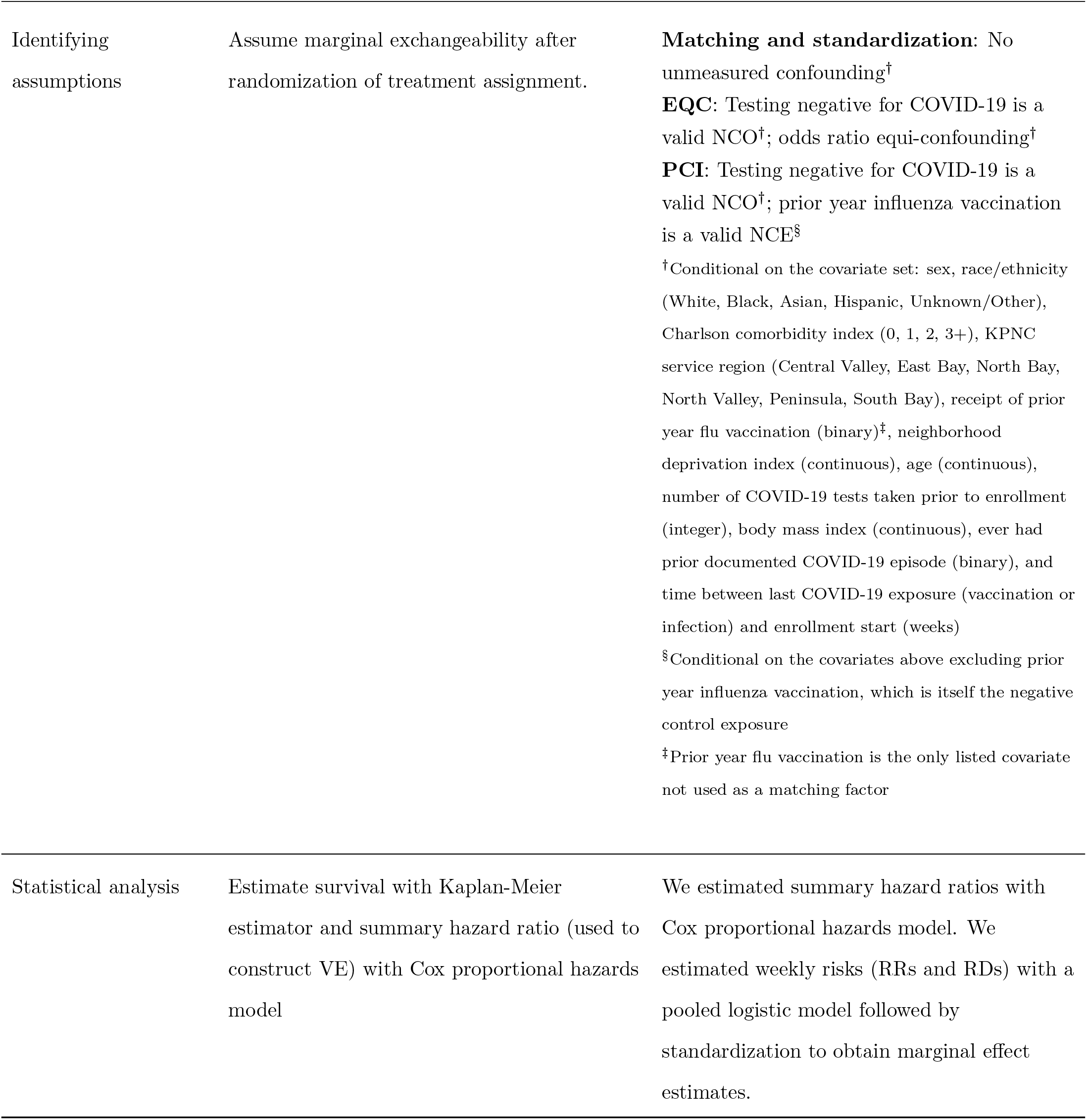
TTE specification and emulation.

| Protocol component | Target Trial | Observational Emulation |
| --- | --- | --- |
| Eligibility criteria | COVID-19 vaccination status of “primary series + 1 additional dose” prior to 09/01/2022;<br>11+ months of continuous KPNC membership prior to start of enrollment;<br>$\geq 18$ years of age between 09/01/2022 and 11/30/2022;<br>Known sex and medical facility;<br>Vaccinated with Pfizer and/or Moderna vaccines;<br>No COVID-19 diagnosis in the 90 days preceding matching | Same as for target trial. Only electronic health records for individuals who met these criteria were included in the analysis |
| Treatment strategies | Vaccination with COVID-19 mRNA bivalent booster (ancestral + BA.4/BA.5) vs.<br>No vaccination at any time | Same as for target trial |
| Treatment assignment | Marginal randomization to one of the two treatment strategies | Individuals were classified according to the treatment strategy consistent with their data during the enrollment period and assumed exchangeability conditional on covariates listed in identifying assumptions (below).<br>If vaccinated at any time during the enrollment period, individuals were assigned to the treated arm or otherwise assigned to the untreated arm |

| **Protocol component** | **Target Trial** | **Observational Emulation** |
| --- | --- | --- |
| Follow-up | Follow-up starts at the beginning of the week of assignment (time zero) and terminates at the end of the week in which an outcome occurs, or loss to follow-up, or after 52 weeks, whichever occurs first | Follow-up starts at the beginning of the week of pair matching during the enrollment period and terminates at the end of the week in which an outcome occurs, or loss to follow-up, or after 52 weeks, whichever occurs first |
| Outcomes | Medically-attended COVID-19 as confirmed by positive nucleic acid amplification test (NAAT) for SARS-CoV-2.<br>(Negative NAAT for SARS-CoV-2 considered an event and/or negative control outcome for EQC and PCI approaches discussed below) | Same as for target trial. |
| Causal contrasts | Intention-to-treat and per-protocol effect | Observational analogue of intention-to-treat and per-protocol effect |
| Identifying assumptions | Assume marginal exchangeability after randomization of treatment assignment. | **Matching and standardization:** No unmeasured confounding<sup>†</sup><br>**EQC:** Testing negative for COVID-19 is a valid NCO<sup>†</sup>; odds ratio equi-confounding<sup>†</sup><br>**PCI:** Testing negative for COVID-19 is a valid NCO<sup>†</sup>; prior year influenza vaccination is a valid NCE<sup>§</sup><br><sup>†</sup>Conditional on the covariate set: sex, race/ethnicity (White, Black, Asian, Hispanic, Unknown/Other), Charlson comorbidity index (0, 1, 2, 3+), KPNC service region (Central Valley, East Bay, North Bay, North Valley, Peninsula, South Bay), receipt of prior year flu vaccination (binary)<sup>‡</sup>, neighborhood deprivation index (continuous), age (continuous), number of COVID-19 tests taken prior to enrollment (integer), body mass index (continuous), ever had prior documented COVID-19 episode (binary), and time between last COVID-19 exposure (vaccination or infection) and enrollment start (weeks)<br><sup>§</sup>Conditional on the covariates above excluding prior year influenza vaccination, which is itself the negative control exposure<br><sup>‡</sup>Prior year flu vaccination is the only listed covariate not used as a matching factor |
| Statistical analysis | Estimate survival with Kaplan-Meier estimator and summary hazard ratio (used to construct VE) with Cox proportional hazards model | We estimated summary hazard ratios with Cox proportional hazards model. We estimated weekly risks (RRs and RDs) with a pooled logistic model followed by standardization to obtain marginal effect estimates. |

**Figure 2:**
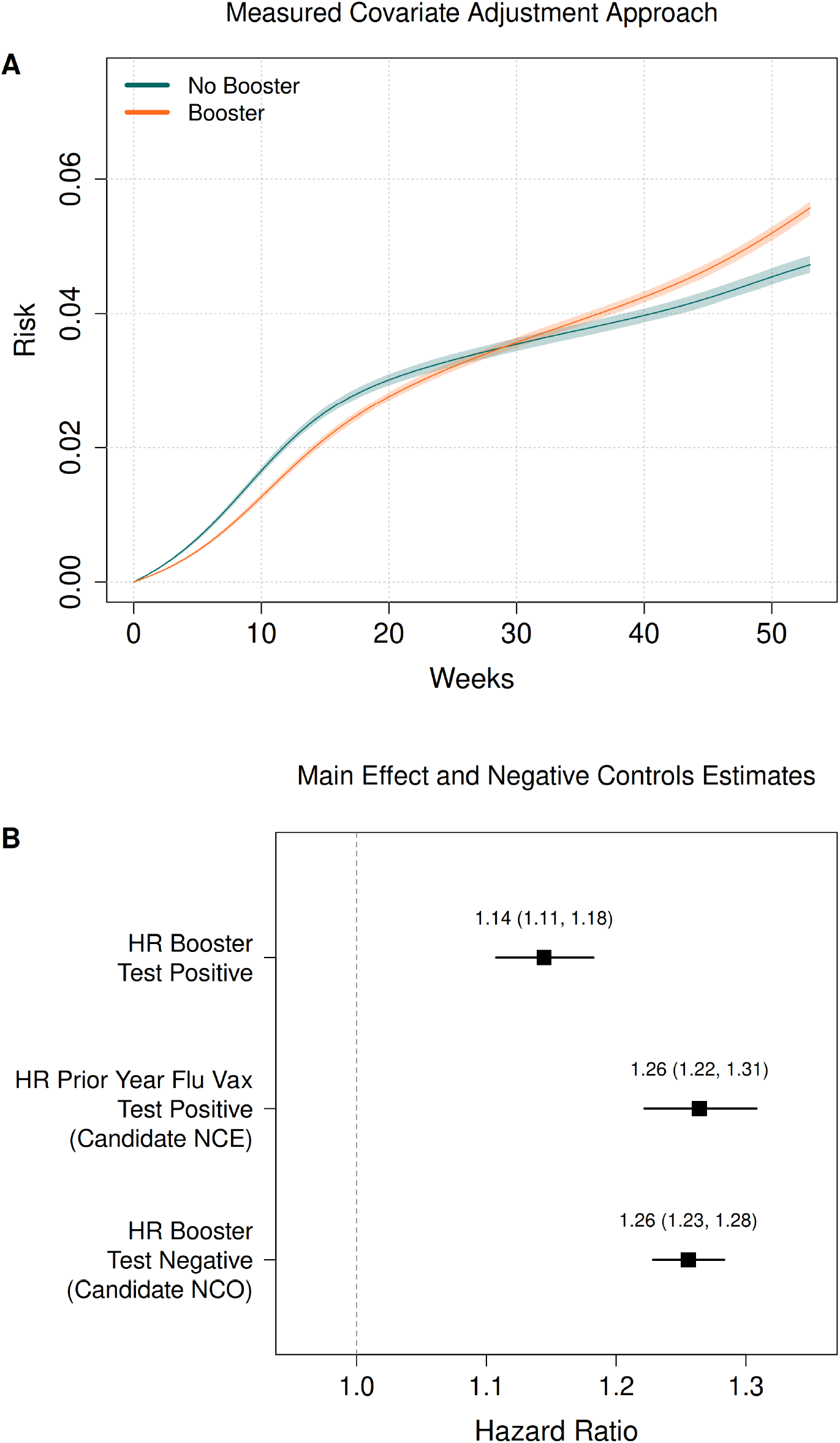
A) Cumulative incidence of testing positive from pooled logistic model under measured covariate adjustment approach. B) Forest plot – hazard ratio of testing positive comparing boosted to unboosted; hazard ratio of testing positive comparing prior year flu vaccine recipients to non recipients; hazard ratio of testing negative comparing boosted to unboosted.

## Alternative Approaches to De-biasing Effect Estimates

To aid in description of alternative approaches, we now introduce some additional notation. Briefly, let *A* be binary vaccination status (1: boosted, 0: unboosted), *X* be measured baseline covariates, *Z* be the negative control exposure, and *U* be unmeasured confounders. We denote by *T*_1_ the (uncensored) time to negative test and *T*_2_ the (uncensored) time to a positive test with *T* = min(*T*_1_, *T*_2_). Let *δ* be the cause of testing event such that *δ* = *j* if *T* = *T*_*j*_, for *j* = 1, 2 (non-SARS-CoV-2 infection, SARS-CoV-2 infection, respectively). In Appendix S1, we provide formal assumptions and identification results for each approach (Section S1.1–S1.7) alongside a simulation study demonstrating performance under a known data generation process (Section S1.9) as well as directed acyclic graphs (DAGs) (Section S1.10).

### Equi-Confounding

The EQC approach relaxes the strong no unmeasured confounding assumption of the measured covariate adjustment approach, assuming instead that (i) testing negative is a valid NCO and (ii) residual confounding is equivalent for test-positive and test-negative illness on the multiplicative scale (assumptions A5 and A6 in Appendix S1.3) (Figure S9c). The first (A5) requires that vaccination have no effect on testing negative. It is most plausible when all test-negatives have an illness, here presumably caused by non-SARS-CoV-2 respiratory pathogens. While there has been considerable debate about this assumption in the TND literature, recent evidence from randomized trials of the immunologically specific COVID-19 vaccine points to its validity [13, 14]. We lean on this empirical evidence to argue that testing negative is a valid NCO. The second assumption (A6), also referred to as equi-confounding, puts a parametric restriction on the form that residual unmeasured confounding must take, namely that it is equivalent for test-positive and test-negative events on the multiplicative scale at all time points.

As we show in Appendix S1.3, when these conditions hold the causal cause-specific hazard ratio, as well as causal survival and cumulative incidence functions are identifiable *among the vaccinated* — equivalent to the average treatment effect among the treated (ATT) on the multiplicative hazard scale. A practical estimator of the causal hazard ratio, is obtained via the difference in cause-specific log hazard ratios described in Algorithm 1. Notably, the coefficient for treatment in the cause-specific model for testing negative yields an estimate of the influence of residual unmeasured confounding that is then used to de-bias the coefficient from the cause-specific model for testing positive. This approach bears similarities to a difference-in-differences analysis on the hazard ratio scale. Estimates of the causal survival and cumulative incidence functions can then be obtained using the procedure described in the same appendix. Valid 95% confidence intervals can be constructed using the bootstrap.

#### Algorithm 1

Equi-confounding approach: Summary hazard ratio

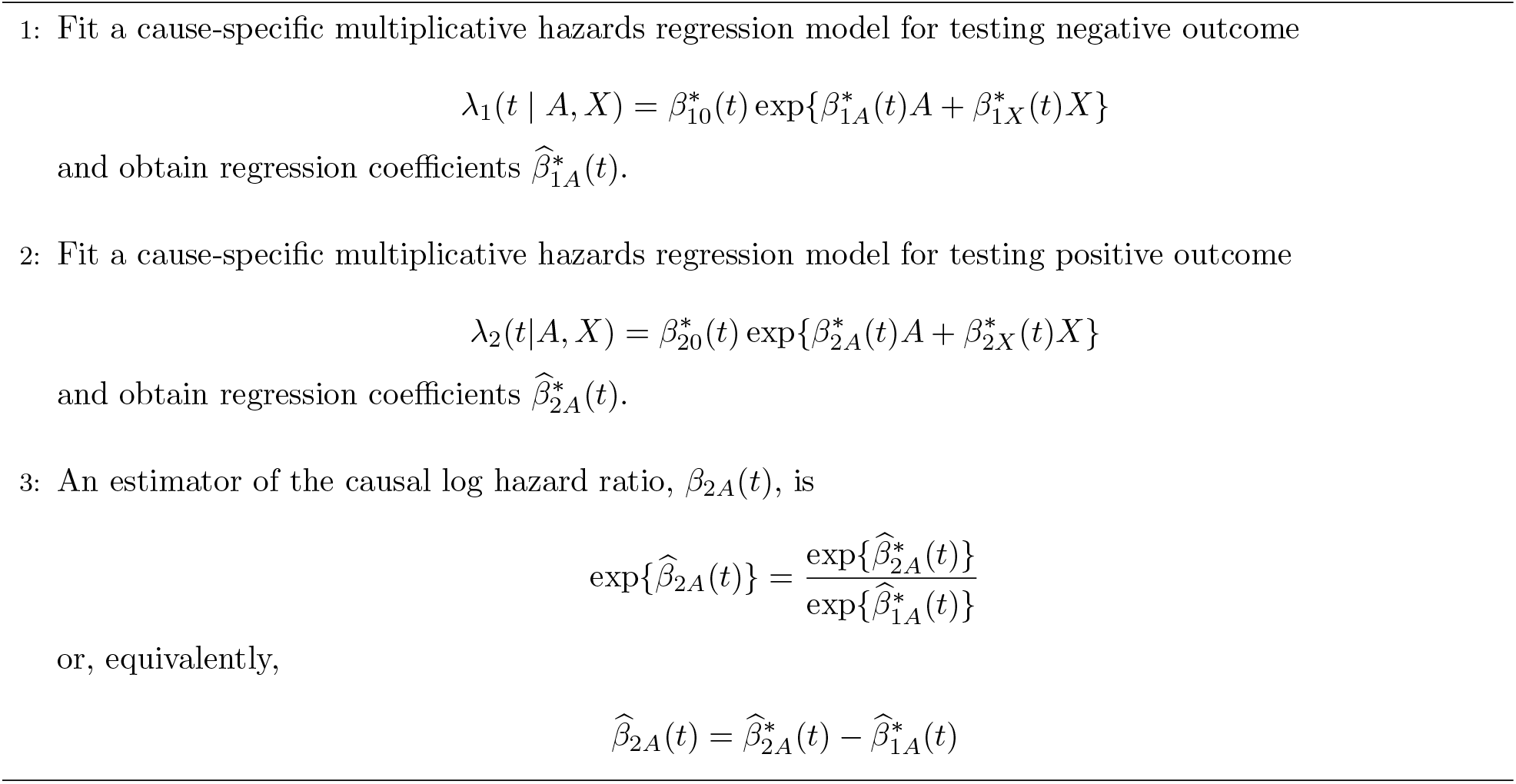

In our applied TTE, we adapted this approach to estimate discrete-time risks for treatment and control groups as in the measured covariate adjustment approach. Weekly estimates of risk were estimated with parametric g-computation using a pooled logistic regression model and Aalen-Johansen-type algorithm to standardize over the observed covariate distribution. We included a product term between treatment and time and modeled time flexibly using natural splines with knot points at 10, 20, 30, 40, and 50 weeks. Nonparametric bootstrapping with 100 replicates was used to calculate weekly percentile-based 95% confidence intervals for risk estimates in each group.

The EQC approach yielded a summary HR of 0.89 (0.86, 0.92) for treatment and 1.04 (1.01, 1.09) for prior year flu vaccination (Table 1). Pooled logistic results show a lower absolute risk of medically attended COVID-19 for the bivalent booster group compared to the no booster group over the entire follow-up period with no overlap of risk curves at any time. The absolute risk difference increases until a maximum at 15 weeks after which we note some waning of VE towards the null but never a complete loss (Figure 3, Table S12).

**Figure 3:**
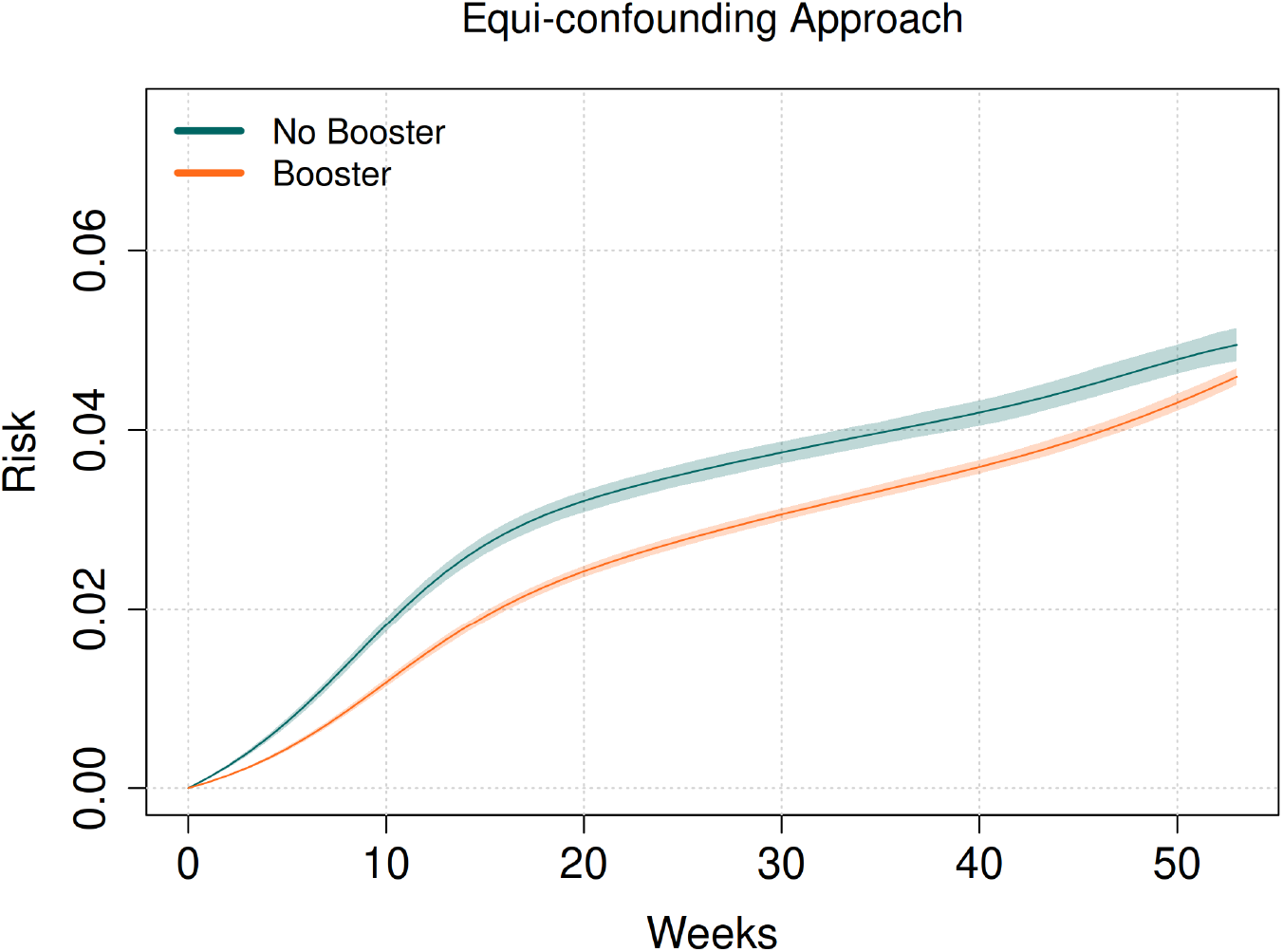
Cumulative incidence curves from pooled logistic models under EQC approach

### Test Negative Design

To illustrate the connection between the EQC approach and the TND, we can obtain a similar estimate of the hazard ratio in the previous section by restricting the analysis to person days with a test (Figure S9b). Among individuals with a PCR test result, we model the odds ratio of testing positive versus testing negative for SARS-CoV-2 using a generalized estimating equation with binomial likelihood and logit link, accounting for clustering within matched sets. This essentially describes the TND and its canonical logistic regression estimator (Appendix S1.4). Indeed, the TND has recently been shown to be valid under the NCO and equi-confounding assumptions described previously [6]. The resulting odds ratio can be used to construct a summary estimate of VE under standard test negative assumptions of a common symptom screen and valid NCO as well as equi-confounding.

The canonical test negative approach yielded a summary HR of 0.90 (0.87, 0.94) for treatment and 1.03 (0.99, 1.08) for prior year flu vaccination (Table 1). Notably, the effect of the NCE has nearly been attenuated to null which we interpret as evidence that much of the unmeasured confounding present in the measured confounding adjustment approach has been removed by the design here. Although the TND may be easier to understand conceptually and implement, we are neither able to unbiasedly assess the effect of treatment on the outcome of interest, nor empirically assess the effect of treatment on testing negative (the NCO) to use as an indicator for residual confounding as we can with the EQC approach.

### Proximal Causal Inference

The PCI approach replaces the (parametric) equi-confounding assumption A6 with a structural assumption about the existence of an additional negative control variable, assumption A6^*\**^. This variable must be unrelated to the outcome and the NCO except through the unmeasured confounder. We assume that an NCE and NCO proxy any unmeasured confounding without affecting or being affected by the primary outcome or treatment, respectively (Figure S9d). We designated prior year flu vaccination as our formal NCE and testing negative for COVID-19 as our NCO. As with our NCO, we again lean on theoretical immunological specificity and empirical evidence to assert that prior year flu vaccination has no effect on COVID-19 risk, justifying our choice of NCE. At most, influenza vaccination modulates innate immune pathways that could influence severity of off-target infections but would likely have minimal impact on test positivity, especially one year removed [15]. We choose prior year rather than current year influenza vaccination to minimize the effect of our NCE on our NCO, as the two variables should only be related through *U*. Antibody titers are measurable beyond a year and cross-reactivity is possible, however, influenza VE estimates beyond a single season are subject to many of the same biases we seek to mitigate here [16]. With valid negative control variables and a completeness assumption - that the NCE and NCO contain a sufficient amount of information about any unmeasured confounding - the causal effect of vaccination is identified via a bridge function that absorbs bias from unmeasured confounders [10] [17].

As we show in Appendix S1.5, when these conditions hold, the causal cause-specific hazard ratio, as well as causal survival and cumulative incidence functions are identifiable now *in the full study population*. A practical estimator of the causal hazard ratio is obtained via the two-stage regression procedure described in Algorithm 2 that imposes minimal restrictions on *U*. This two-stage estimation procedure is (somewhat) similar to instrumental variables estimation. Estimates of the causal survival and cumulative incidence functions can then be obtained using the procedure described in the same appendix where we also discuss some subtleties in the PCI approach related to modeling hazards on the additive or multiplicative scale.

#### Algorithm 2

Proximal inference approach: Summary hazard ratio

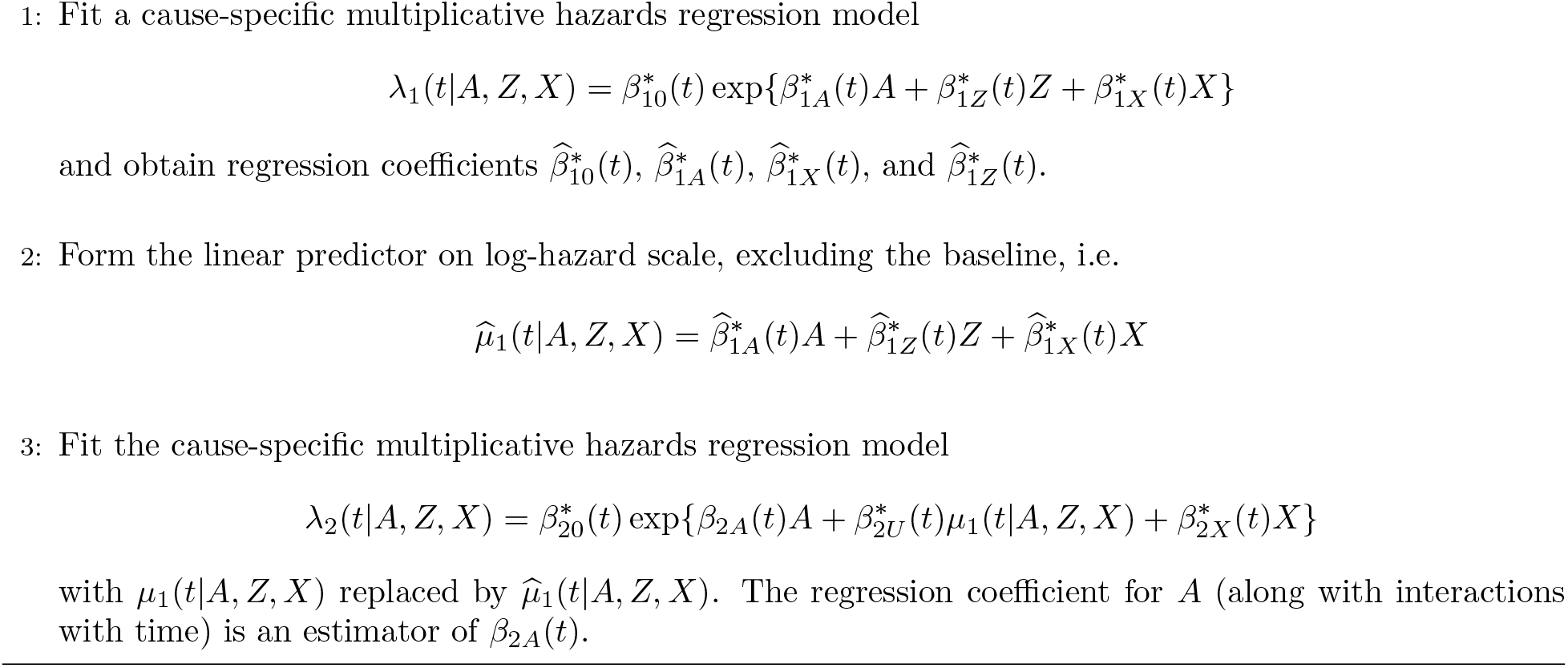

We similarly adapted the PCI approach to estimate discrete-time risks for treatment and control groups. Weekly estimates of risk were estimated with parametric g-computation using a pooled logistic regression model in two stages. In the first stage we modeled the discrete time hazard of the NCO. The log odds of the NCO was included as a covariate in the second stage pooled logistic model for the discrete time hazard of the primary outcome. A time-varying log odds ratio for treatment was extracted from the second stage model at reference covariate values with the bridge function fixed at zero. Counterfactual discrete-time hazards under treated and untreated scenarios were obtained by scaling each individual’s observed hazard predictions by the exponentiated treatment log odds ratio, adding the treatment effect for untreated individuals under the *A* = 1 intervention and removing it for treated individuals under the *A* = 0 intervention. Cause-specific cumulative incidence under each intervention was computed for each individual again using a Aalen–Johansen-type algorithm and then standardized over the observed covariate distribution. Nonparametric bootstrapping with 200 replicates was used to calculate weekly percentile-based 95% confidence intervals for risk estimates in each group.

The proximal causal inference approach yielded a summary HR of 0.84 (0.76, 0.90) for treatment (Table 1). No estimate for the effect of the NCE is obtained from the second stage model. Pooled logistic results show a lesser absolute and relative magnitude of booster protection compared to the EQC approach. We observe confidence intervals inclusive of null values by 36 weeks and RD/RR point estimates waning to null by 51 weeks (Figure 4, Table S13).

**Figure 4:**
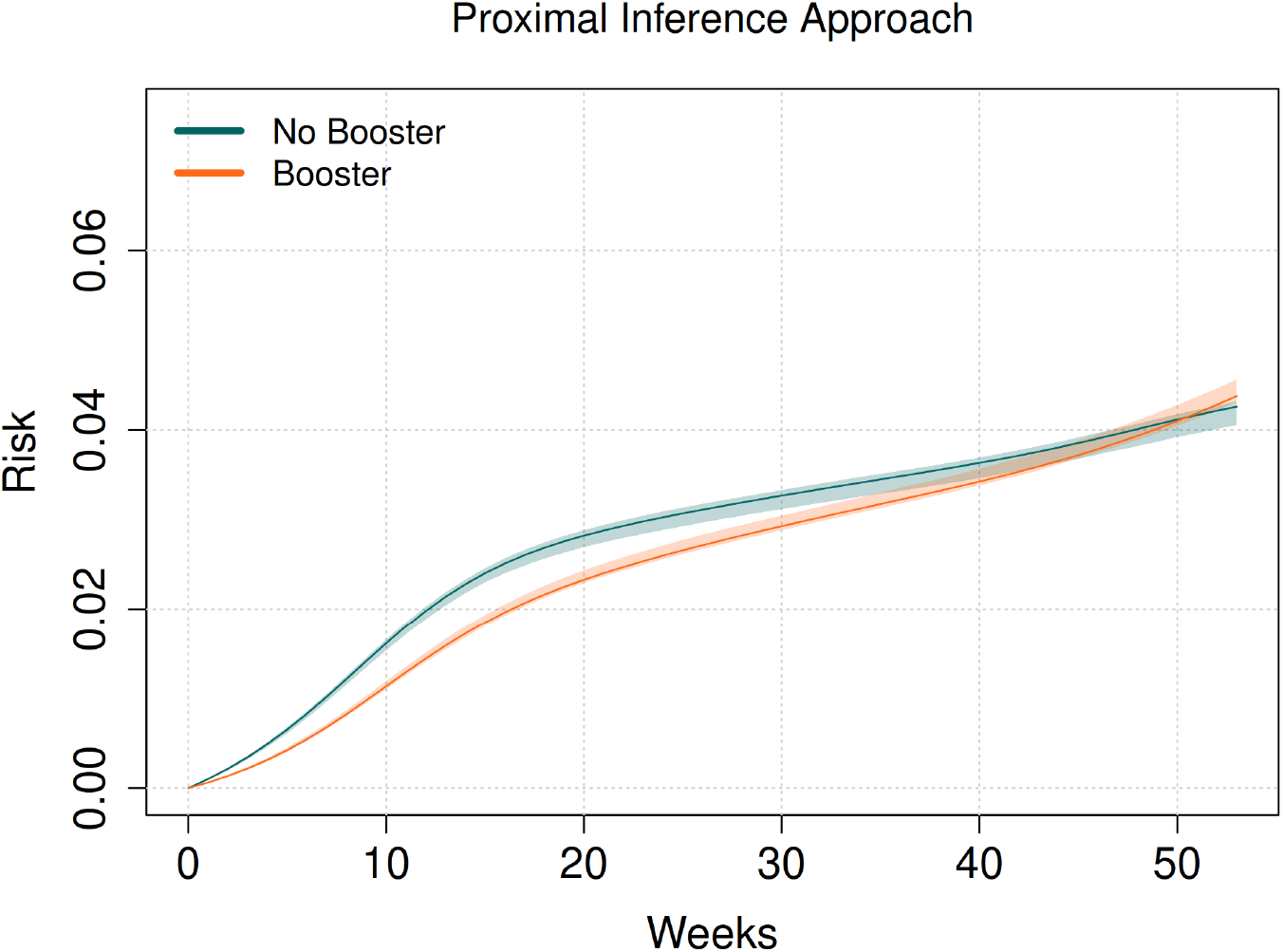
Cumulative incidence curves from pooled logistic models under PCI approach

## Discussion

We present two approaches for debiasing VE estimates obtained in a cohort setting by extending the target trial emulation framework beyond methods that rely on confounding adjustment alone. We implemented these approaches using a large electronic healthcare database and compared the results to an emulation using a standard covariate adjustment approach with no efforts to debias effects, as well as a canonical test negative study design.

Unmeasured confounding due to differences in health seeking behavior makes it challenging to design studies that aim to estimate VE against mild or moderate outcomes such as infection or medically attended illness. The literature describing many seasonal pathogens is skewed towards VE estimates against severe outcomes such as hospitalization and death, leaving substantial uncertainty about protection against non-severe outcomes relevant to the vast majority of the at-risk population. We know from the study of influenza and other pathogens that prevention of mild infections in lower risk populations can reduce the burden of severe infection and mortality in higher risk populations through indirect effects [18].

The body of literature describing the VE of this specific booster formulation is relatively small, but the estimates that do exist are relevant for comparison and exemplify the difficulties in studying protection against mild or moderate disease. VE against mild infection ranges from 28% to 56% (HR range: 0.72-0.44) for the few studies that considered this outcome [19, 20, 21, 22]. Tartof et al. [20] estimate a VE of 28% (22% - 33%) against outpatient encounters using a canonical test negative design in a cohort of Kaiser Permanente Southern California members. The majority of studies we found only considered severe outcomes such as hospitalization or death where VE ranged from 32% to 72% (HR range: 0.28-0.68) [23, 24, 25, 26, 27]. Additionally, study time horizons in the existing literature do not extend beyond 6 months, presumably due to these same uncertainties that compound with longer follow up periods. For comparison, we estimated a 6-month RR of 0.84 (0.81, 0.88) by EQC and 0.93 (0.91, 1.00) by PCI.

We note the differences between the results of the EQC and PCI approaches, where the former suggests that complete waning of bivalent booster protection does not occur over the course of a full year while the latter suggests that it may occur as soon as 36 weeks (at which point the RR confidence interval includes the null value). This disagreement is expected, as these bias adjustment approaches operate under different assumptions about unmeasured confounding. The EQC approach corrects for unmeasured confounding of exposure risk and differences in testing behavior between the treated and untreated groups under the assumption that these operate equivalently across illness types on the multiplicative scale. The assumption of equi-confounding is plausible over the course of a year provided the source of illness remains unknown to individuals prior to testing, however, we remain cautious of scenarios that would invalidate this assumption. Should unmeasured confounders such as social gathering behaviors begin to create a differential risk of COVID-19 and influenza on the multiplicative scale, this critical assumption would no longer hold. Equi-confounding would also be violated if COVID-19 or influenza disease presentation changes such that the relationship between vaccination status and testing propensity differs across illness. The PCI approach relaxes equi-confounding and instead assumes a valid NCE and NCO. Under valid negative control variables, the PCI approach accounts for bias from any unmeasured confounding *U*, including health-seeking behavior and differential illness risk, without requiring that these effects operate equivalently across illness types. The validity of either of these two approaches relies on the validity of the alternative assumptions that are needed for these methods in the specific scenario where they are applied. These alternatives to a confounding adjustment approach come at the cost of requiring the existence of valid NCO and NCE as well as the possibility of measuring them.

We made the decision to truncate follow-up at 1 year due to concerns of major deviations from assigned treatments and the validity of our assumptions over longer time horizons. A large proportion of the population who received a bivalent booster during this study enrollment period in late 2022 subsequently received another COVID-19 booster at the end of 2023; the same was true for a smaller proportion of the untreated population. The accumulation of undetected infections becomes more likely over time and presents a confounding challenge beyond the capabilities of the approaches presented here. At-home testing availability during our study period could have also contributed to some infections going undocumented; prior infection, regardless of symptom status, confers similar protection against future infection to that of the booster vaccination under study and could lead to bias in an unpredictable direction if accumulated differentially across treatment groups. We were less concerned with the dilution of the ITT effect due to untreated individuals receiving a booster and deviating from their assigned treatment strategy, as vaccinations only occurred at minimal levels throughout the summer months following our enrollment period.

The equi-confounding and proximal causal inference approaches for emulating a target trial represent some of the first applications of recent theoretical work improving the estimation of VE in settings with complex confounding. These methods show promise for helping diagnose and resolve the potentially thorny problem of residual unmeasured confounding for symptom-driven outcomes as large cohort data becomes more accessible and as the testing landscape changes with the advent of multiplex and at-home testing. More generally than VE estimation, these methods illustrate TTE as a principled approach for appropriately designing observational comparative effectiveness studies and analyzing the data under alternative assumptions.

## Data Availability

The individual-level electronic health record data analyzed in this study contain protected health information and cannot be shared publicly, in accordance with institutional policies and data use agreements with Kaiser Permanente Northern California. Aggregate results supporting the findings are contained in the manuscript. Analysis code is available at

https://github.com/BeauSchaeffer/tte-tnd

## Supplementary Material: Debiasing estimates of vaccine effectiveness under the target trial framework

### S1 Notation, assumptions, and proofs

#### S1.1 Preliminaries

Let *A* be binary vaccination status, *X* be measured baseline covariates, *Z* be the negative control exposure, and *U* be unmeasured confounders. We denote by *T*_1_ the (uncensored) time to negative test and *T*_2_ the (uncensored) time to a positive test with *T* = min(*T*_1_, *T*_2_). Let *δ* be the cause of testing event such that *δ* = *j* if *T* = *T*_*j*_, for *j* = 1, 2. For convenience, we also define an tri-level testing indicator of testing status at a landmark follow up time 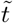 (e.g., 1-year) as 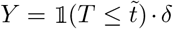*· δ*. Let *C* be the right censoring time due to other reasons (e.g. loss to follow up, administrative censoring). For each subject let *T* ^*\**^ = min(*T, C*) denote the observed censored event time and Δ = 1(*T ≤ C*) *· δ*, such that the observed data constitutes independent and identically distributed (iid) samples of (*X, Z, A, T* ^*\**^, *Y*, Δ, *δ*).

We define the cause-specific hazards function *λ*_*j*_(*t*|*·*) as

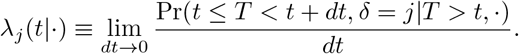

Likewise, we define the survival function as *S*(*t*|*·*)≡ Pr(*T > t*|*·*) and the cumulative incidence function as *F*_*j*_(*t*|*·*)≡ Pr(*T ≤ t, δ* = *j*|*·*). We denote the potential event time under an intervention that sets *A* = *a* by *T*^*a*^ and therefore define counterfactual corollaries of the above functions 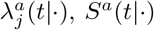, and 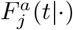 by replacing *T* with *T*^*a*^.

#### S1.2 Assumptions

We assume the following conditions hold for *a* ∈ {0, 1}:

(A1) *Consistency of potential outcomes*. For all individuals *i*, we have 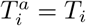 when *A*_*i*_ = *a*.

(A2) *Exchangeability conditional on* (*U, X*). That is, *T*^*a*^ *╨ A* | (*X, U*).

(A3) *Positivity*. For all covariate patterns in support of *X* and *U*, we have Pr(*A* = *a*|*X, U*) *>* 0.

(A4) *Independent censoring conditional on* (*A, X, Z*). That is, *C ╨ T* | (*A, Z, X*).

Note that these are the same as the identification conditions for the measured covariate adjustment approach when *U ╨ A* | *X* or *U* is empty (i.e., a degenerate constant) as represented in the directed acyclic graph in Figure S9a. However, in what follows we allow for the possibility of residual unmeasured confounding through *U*.

To facilitate identification arguments below, we define the structural (semi-parametric) multiplicative cause-specific hazard models

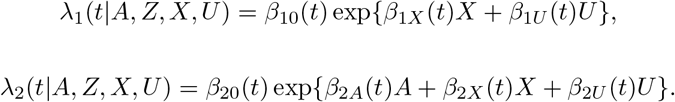

Note that we allow the influence of *A, X*, and *U* to vary over time; we could allow for arbitrary interactions between *A* and *X* or *X* and *U* (although not between *A* and *U*), but suppress these for now to increase readability.

#### S1.3 Equi-confounding approach

Here we describe the identification of VE under the EQC approach, extending the arguments in Boyer, Li, Shi, and Tchetgen Tchetgen [1] to a setting with right censoring. Unlike the measured covariate adjustment approach the EQC approach allows for unmeasured confounding (via non-degenerate *U*), but requires additional assumptions for identification of the causal parameters of interest, namely:

(A5) *Testing negative is a valid NCO*. That is, 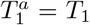 almost surely, 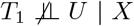, and *A ╨ T*_1_ | (*X, U*).

Equivalently, treatment has no causal effect on the test-negative event time, implying for all *t* that

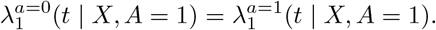

(A6) *Equi-confounding on multiplicative scale*. Residual confounding after conditioning on *X* is equivalent for test-negative and test-positive outcomes on the relative scale

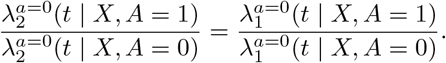

In section S1.6 we consider an alternative to A6 on the hazard scale rather than the additive scale.

**Causal hazard ratio** Starting with the completely nonparametric case, consider the causal time-varying cause-specific hazard ratio *among the vaccinated*

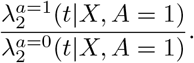

Multiplying by unity in the form of 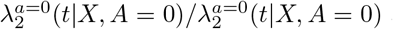 and applying consistency A1 yields the following decomposition

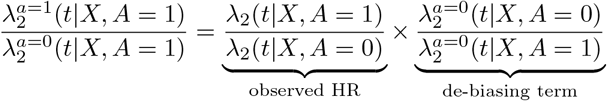

Under the NCO assumption A5 and equi-confounding A6, the de-biasing term is identified by the ratio of cause-specific hazards of testing negative, therefore

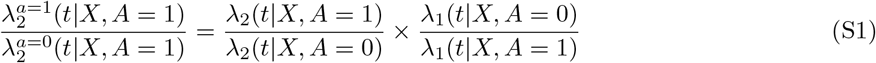

Further insight can be obtained from the structural model proposed above. Consider the identification of exp*{β*_2*A*_(*t*)} which is the causal time-varying hazard ratio for testing positive and which can be used to derive other quantities such as cumulative incidence or survival.

Under consistency A1 we have that

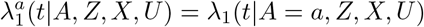

and

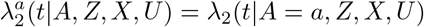

The structural model implies *T*_1_ is a valid negative control outcome A5 as

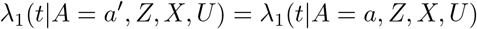

for all combinations of *a* and *a*^*′*^. It also implies that *Z* is a valid negative control exposure as *Z ╨* (*T*_1_, *T*_2_) | (*A, X, U*) and can be removed and ignored for now as it is not required for identification under equi-confounding.

The equi-confounding assumption implies *β*_1*U*_ (*t*) = *β*_2*U*_ (*t*) for all *t*, i.e. the net influence of unmeasured confounding is the same on (multiplicative) hazard ratio scale for test-positive and test-negative outcomes at all times over follow up. Note that we still allow for the effect of *U* to be time-varying as long as it applies equally to both outcomes. Denote *β*_*U*_ (*t*) := *β*_1*U*_ (*t*) = *β*_2*U*_ (*t*), taking expectation of *λ*_1_(*t*|*A, Z, X, U*) with respect to (*A, X*) we have

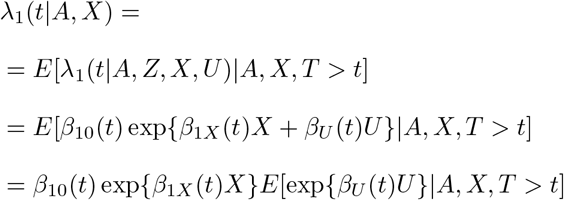

such that

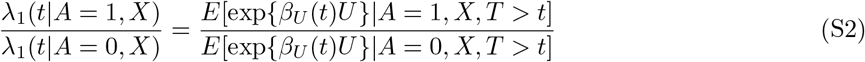

Likewise, taking expectation of *λ*_2_(*t*|*A, Z, X, U*) with respect to (*A, X*) we have

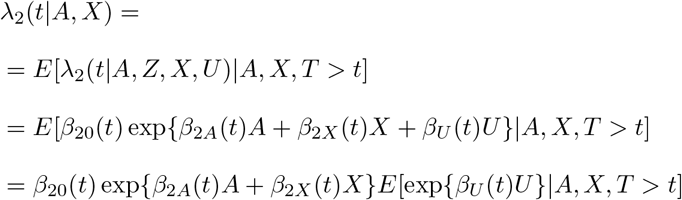

such that

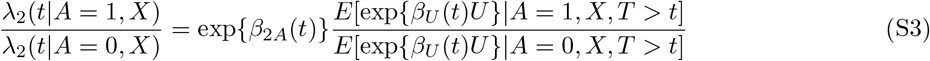

Dividing Equation S3 by Equation S2, we find that

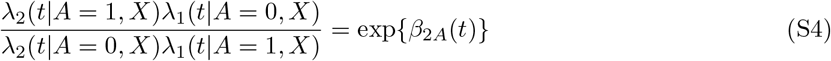

motivating the estimation steps described in Algorithm 1.

##### Marginal survival and cumulative incidence

Beyond the summary hazard ratio, Assumptions A5 and A6 also identify the counterfactual survival and cumulative incidence functions among the treated (the ATT). Returning to the nonparametric identification of Equation S1 the NCO assumption A5, and equi-confounding A6 give

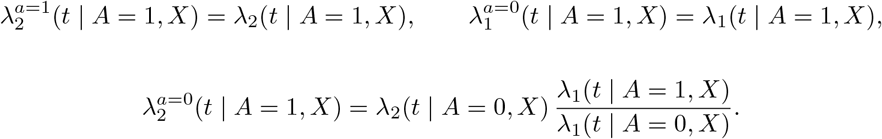

Since the survival of a counterfactual process equals the exponential of its negative integrated cause-specific hazards, and the identified quantities above are precisely those hazards (conditioning on the counterfactual risk set *{T*^*a*^ *> t}*),

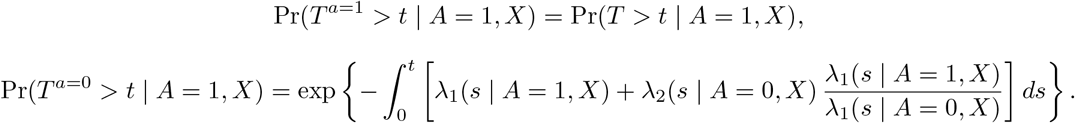

Standardizing over the covariate distribution among the treated yields the marginal counterfactual survival, cause-specific hazard, and cumulative incidence

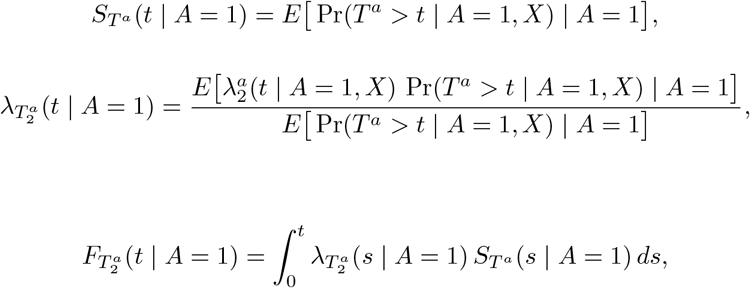

with 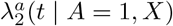 as above. Estimation is summarized in Algorithm 3.

###### Algorithm 3

Marginal survival and cumulative incidence, multiplicative equi-confounding

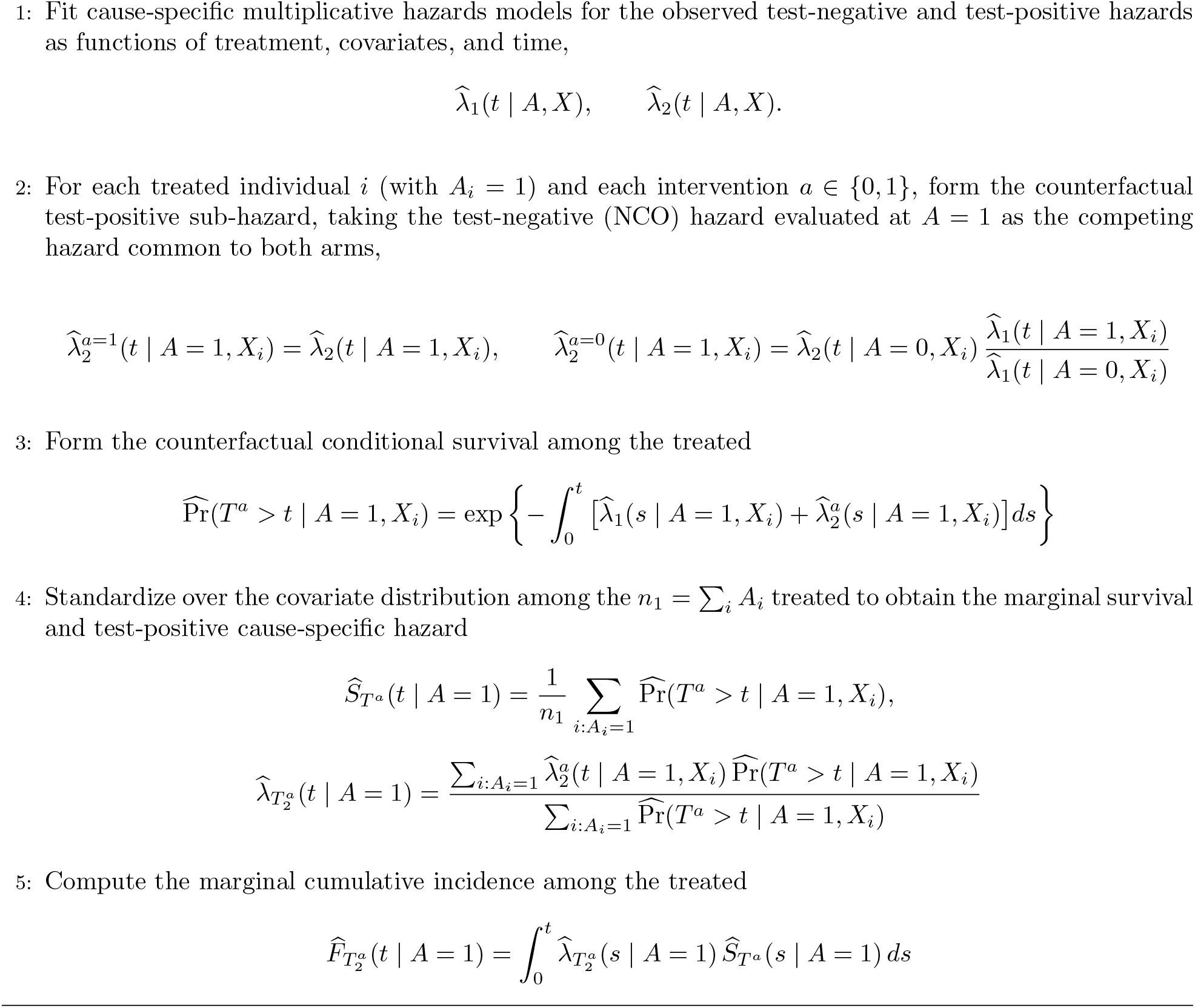

#### S1.4 Test negative design approach

The TND yields a similar summary estimate as the EQC approach under assumptions of a valid NCO and equi-confounding. To elaborate further, an interesting aspect of competing risks is that, among individuals at risk at time *t*, the conditional odds of cause 2 vs cause 1 given an event at *t* equals the ratio of cause-specific hazards, i.e.

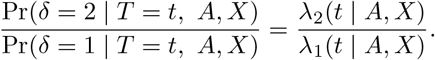

This suggests a similar result as Algorithm 1 for the EQC approach may be obtained by restricting to person-days with a test and estimating the logistic regression

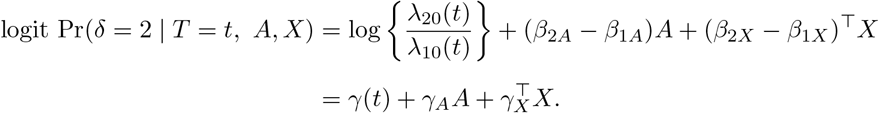

where *γ*_*A*_ estimates the log difference in cause-specific hazards and under assumptions A5 and A6, recovers the causal effect of VE.

#### S1.5 Proximal inference approach

Here we describe assumptions necessary for the identification of VE under the PCI approach as well as causal survival and cumulative incidence functions. Like the EQC approach, PCI allows for the unmeasured confounding via *U*. However, whereas EQC places strong parametric restrictions on the relationship between *U* and the NCO and focal outcome, PCI weakens this through the identification of another counfounding proxy—the NCE. We therefore replace assumption A6 with A6^*\**^ below:

(A6^*\**^) *Z is a valid NCE*. That is, 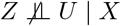, and *Z ╨* (*T*_1_, *T*_2_) | (*X, U*).

While nonparametric approaches via integration methods involving the so-called “confounding bridge” functions are possible [2], here we introduce easier to implement methods the build on traditional regression strategies. To do so, we have to impose a condition on *U* that is weaker than equi-confounding A6 but not fully nonparametric.

(A7^*\**^) *U is a location-shift conditional on T > t, i*.*e*.. That is,

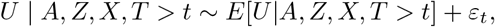

with *E*[*ε*_*t*_] = 0 and *ε*_*t*_ *╨* (*A, Z, X*) | *T > t*

For ease of exposition, consider the case that the the mean-model is linear in (*A, Z, X*)

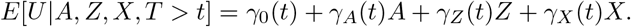

We note, however, that results hold for more general nonlinear *E*[*U* |*A, Z, X, T > t*].

Taking the expectation of *λ*_1_(*t*|*A, Z, X, U*) with respect to (*A, Z, X, T > t*) we have

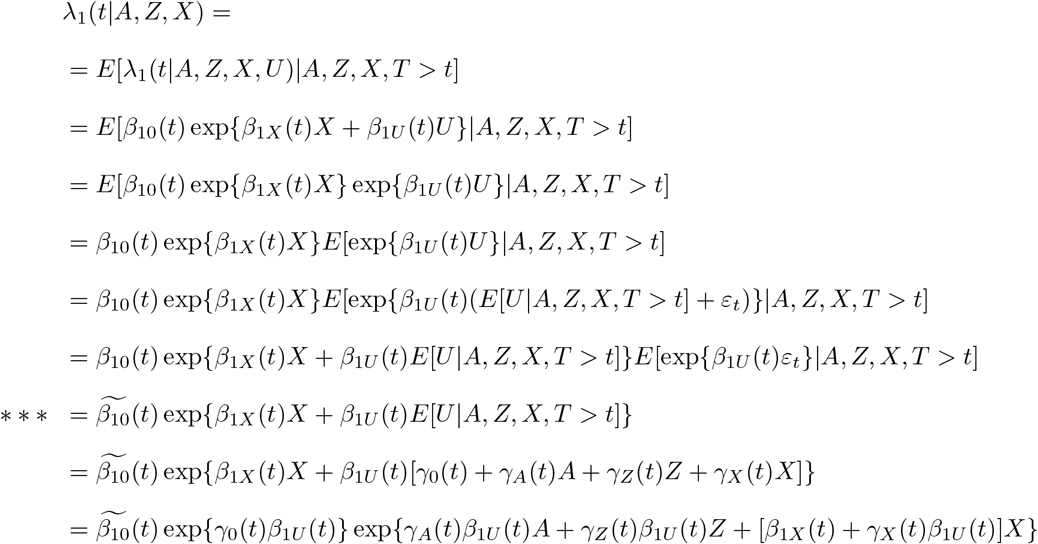

where

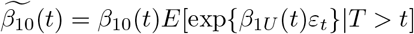

which does not depend on (*A, Z, X*) given *T > t* since *ε*_*t*_ *╨* (*A, Z, X*) | *T > t*, and we assume this moment generating function exists. Therefore, we can define

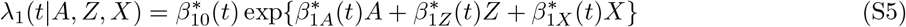

with

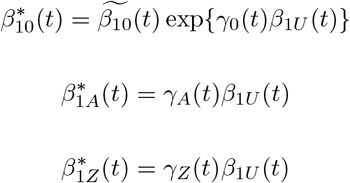

and

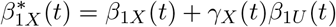

Notice from stared line (*\* * \**) above that

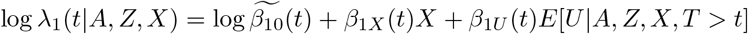

Therefore, solving for *E*[*U* |*A, Z, X, T > t*], we have

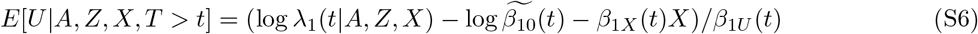

This step requires *β*_1*U*_ (*t*)*≠* 0, i.e. the unmeasured confounder is relevant to the test-negative outcome. Separating *β*_2*A*_(*t*) from the bridge coefficient in the second stage additionally requires *γ*_*Z*_(*t*)*≠* 0 (a relevant NCE, Assumption A6^*\**^), so that log *λ*_1_(*t*|*A, Z, X*) varies with *Z* given (*A, X*).

Now, taking expectation of *λ*_2_(*t*|*A, Z, X, U*) with respect to (*A, Z, X, T > t*) we have

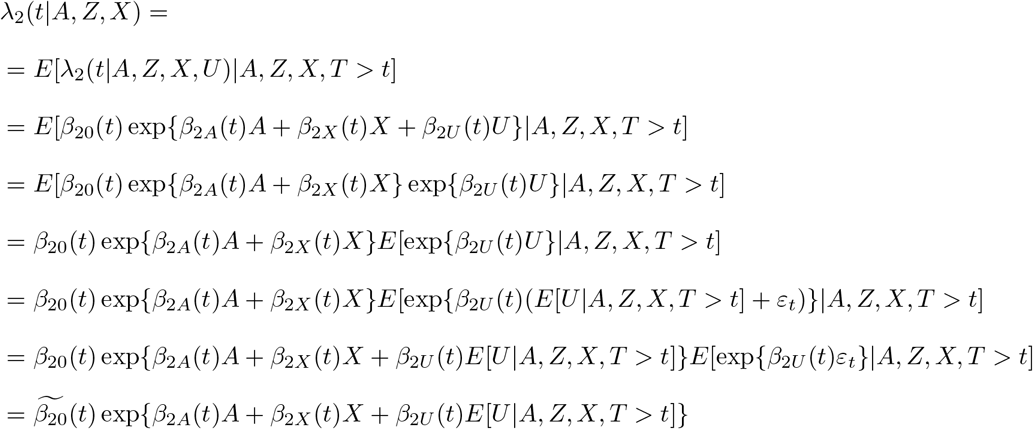

Plugging in the result in Equation S6 for *E*[*U* |*A, Z, X, T > t*], we have

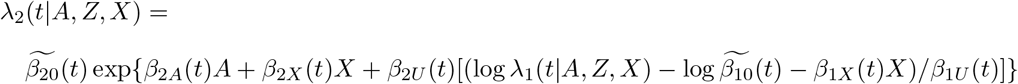

Combining and redefining terms, this suggests

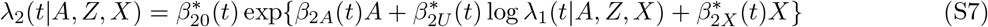

with

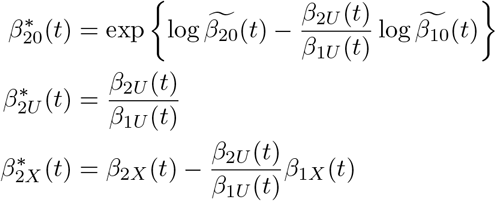

The bridge log *λ*_1_(*t* | *A, Z, X*) carries the stage-one time-varying intercept log 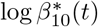, a function of time alone. It is therefore collinear with the second-stage baseline log 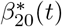 and may be removed from the bridge without changing *β*_2*A*_(*t*), since the second-stage time baseline absorbs it. In estimation described in Algorithm 2 we subtract this baseline (the stage-one fit at reference covariates, on the link scale).

##### Marginal survival and cumulative incidence

For marginal survival and cumulative incidence estimates, note that we have

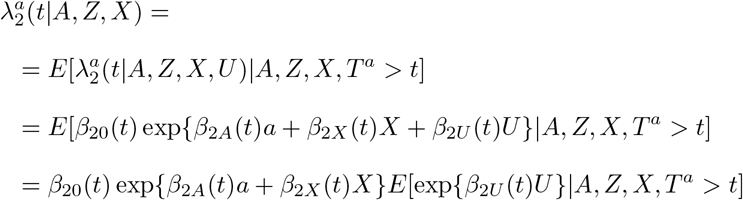

and

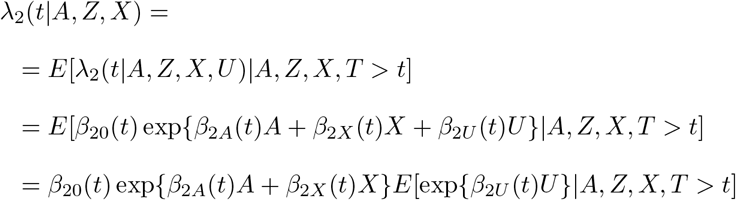

So the relationship between counterfactual and observed cause-specific hazards is

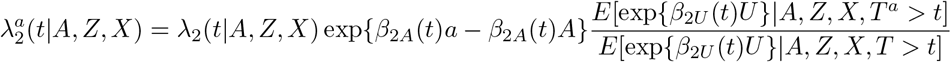

which is not identifiable from the observed data, due to the unavailability of *U*, unless

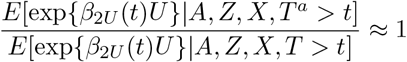

which occurs when events are rare (and therefore survivor distributions are approximately baseline distribution of *U*, i.e. negligible depletion)^1^. Thus, marginal counterfactual survival and cumulative incidence are not recoverable via proximal inference methods using simple regression without further assumptions. This is despite the fact that the conditional causal hazard ratio *β*_2*A*_(*t*) is identifiable. It also does not apply to fully nonparametric approaches, although these require solving a complex integral equation requiring different assumptions for the existence of valid solutions.

In the main text, we use the negligible depletion approximation and assume

(A8^*\**^) *negligible depletion on U over* [0, *t*]. That is,

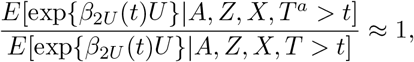

such that

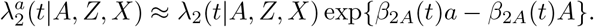

This allows for estimation of the marginal cumulative incidence and survival function using simple regression methods as defined below. In the observed data the risk of the test positive outcome is only about 4 to 5 percent, so negligible depletion is possible. Furthermore, as we show in Section S1.6, under an additive structural model, the counterfactual survival and cumulative incidence functions are identified exactly via regression methods without the need for an assumption about negligible depletion. Thus, we can compare our approximation with the exact results under the additive model. Figure S2 shows the difference between the two is minimal.

Applying iterated expectations, the counterfactual survival is

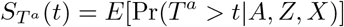

Because it is unaffected by treatment, we also have for the competing risk

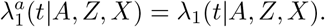

Combining with the assumed switching function under negligible depletion we have:

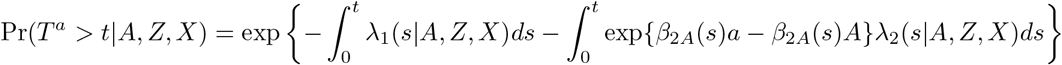

The marginal counterfactual hazards are then

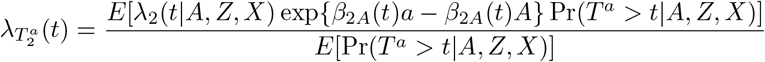

and we have

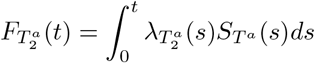

motivating the estimation approach described in Algorithm 4 below.

###### Algorithm 4

Marginal survival and cumulative incidence, multiplicative hazards (approximation)

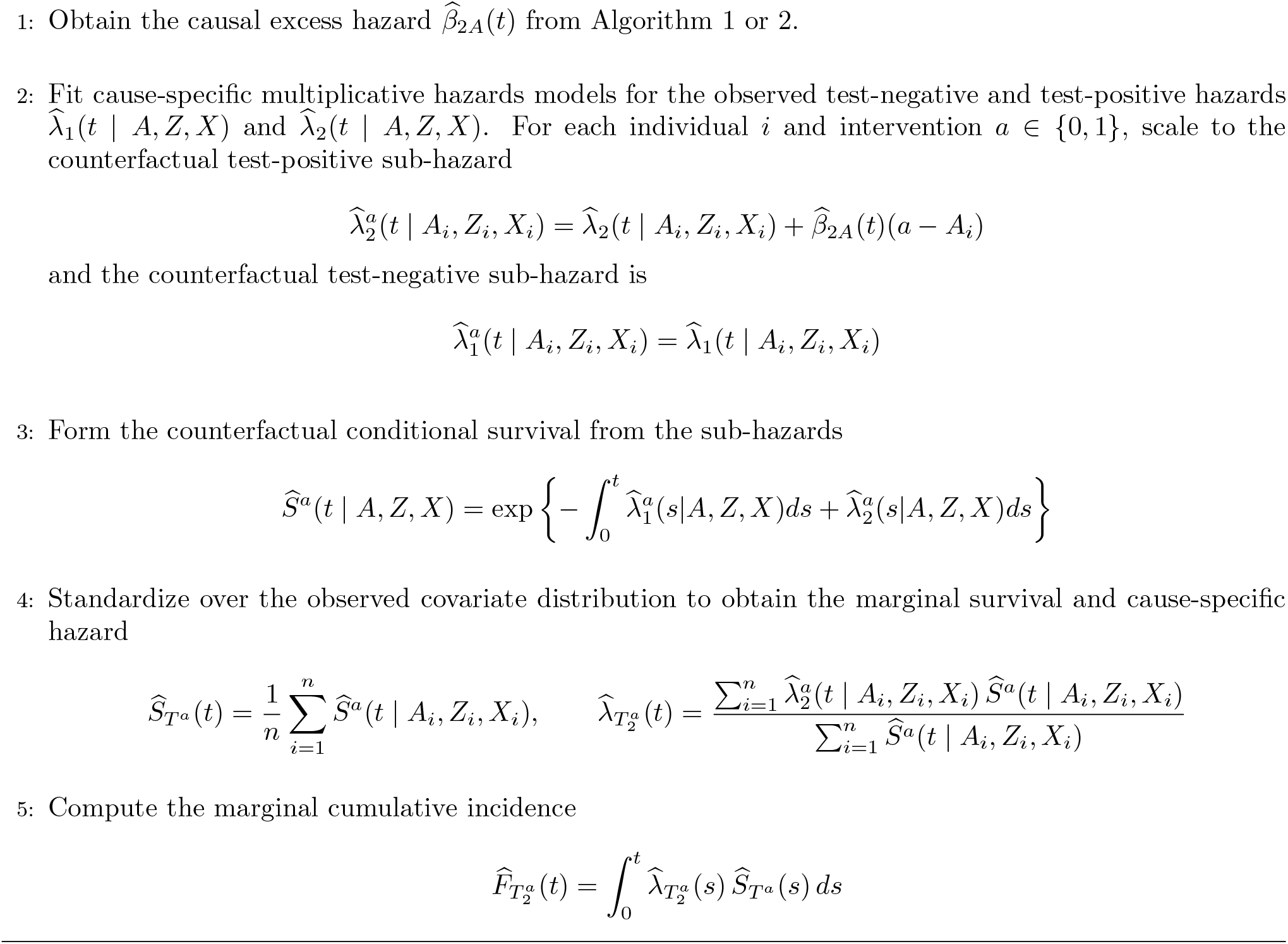

#### S1.6 Additive hazards scale

The identification results above have direct analogs under an *additive* rather than multiplicative structural definition of the cause-specific hazards, following the proximal survival analysis results of Li, Linderman, Shi, and Tchetgen [3]. An advantage of the additive definitions is that, because the intervention enters the hazard additively and independently of *U*, the counterfactual survival and cumulative incidence functions are identified *exactly*, without the negligible depletion approximation noted for the multiplicative case.

We posit the structural additive cause-specific hazard models

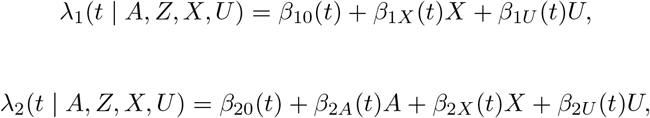

so that *β*_2*A*_(*t*) is now the causal time-varying *excess* cause-specific hazard of testing positive (a hazard difference rather than a ratio). As before *λ*_1_ does not depend on (*A, Z*), so testing negative is a valid NCO (Assumption A5) and *Z* is a valid NCE (Assumption A6^*\**^). Under consistency, exchangeability, positivity and the structural model above the counterfactual cause-specific hazards are

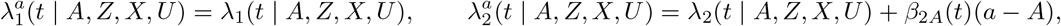

where, crucially, the intervention contrast *β*_2*A*_(*t*)(*a − A*) does not involve *U*.

##### Equi-confounding

Under an additive hazards model, we can consider an analog equi-confounding condition to Assumption A6 on the additive scale^2^, namely

(A6^*†*^) *Equi-confounding on additive scale*. Residual confounding after conditioning on *X* is equivalent for test-negative and test-positive outcomes on the additive scale

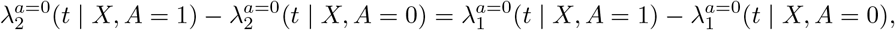

which (when residual confounding is present) is equivalent to the requirement that *β*_1*U*_ (*t*) = *β*_2*U*_ (*t*) =: *β*_*U*_ (*t*) in the structural model. Marginalizing each structural hazard over *U* conditional on being at risk is exact for additive hazards, since the expectation of a sum is the sum of expectations and no frailty term arises:

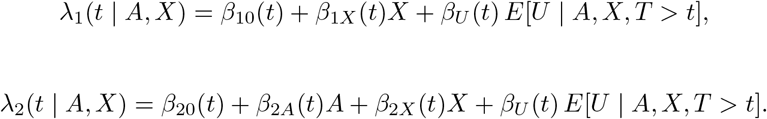

Because the residual confounding term *β*_*U*_ (*t*) *E*[*U* | *A, X, T > t*] is shared, it cancels in the difference-in-differences

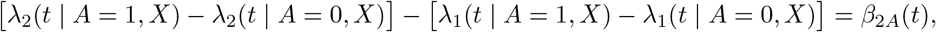

so *β*_2*A*_(*t*) is identified as the observed test-positive cause-specific hazard difference minus the observed test-negative (NCO) hazard difference (Algorithm 5). This is the additive analog of Algorithm 1, with the difference of log hazard ratios replaced by a difference of hazard differences.

###### Algorithm 5

Equi-confounding approach, additive hazards

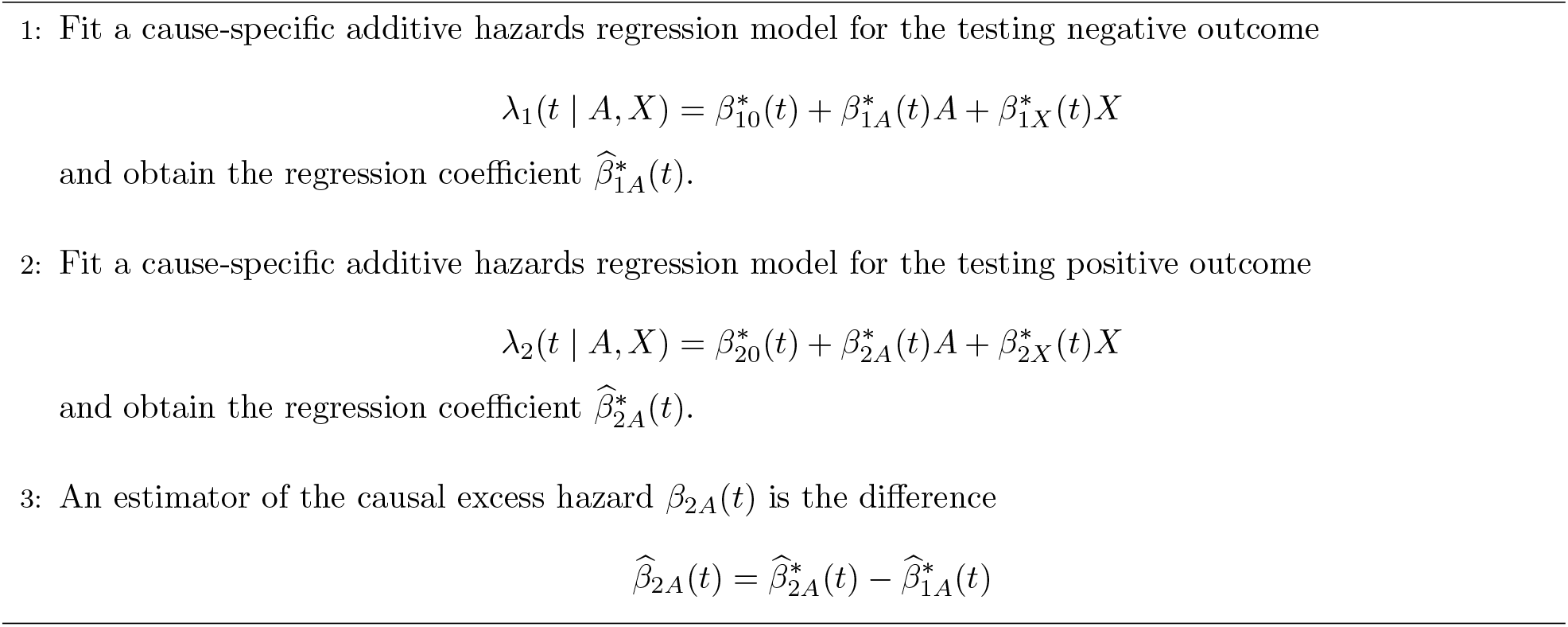

##### Proximal inference

Although proximal inference does not require additivity in general, as shown by Li, Linderman, Shi, and Tchetgen [3], several convenient properties arise when using additive hazard models under the regression-based (as opposed to the nonparameteric) approach to survival data. Retaining the location-shift model^3^ for *U* | *A, Z, X, T > t* from Section S1.5, marginalizing *λ*_1_ over *U* gives

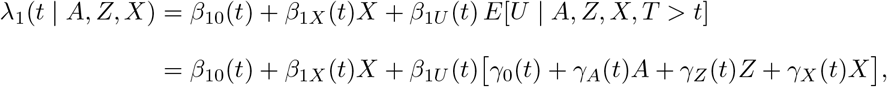

where the mean-zero residual *ε*_*t*_ drops out exactly. Collecting terms,

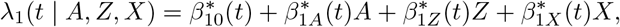

With 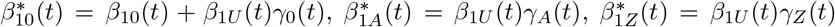, and 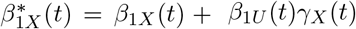. Solving for *β*_1*U*_ (*t*) *E*[*U* | *A, Z, X, T > t*] = *λ*_1_(*t* | *A, Z, X*) *− β*_10_(*t*) *− β*_1*X*_ (*t*)*X* and substituting into the marginalized second hazard

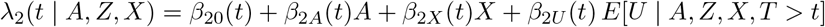

yields the two-stage additive reduced form

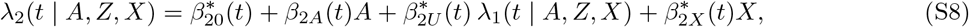

with

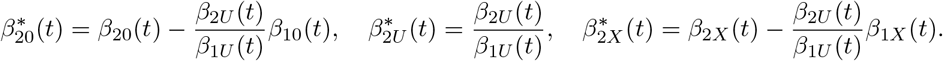

The coefficient on *A* is the causal excess hazard *β*_2*A*_(*t*). This is the additive analog of Algorithm 2: the fitted NCO hazard *λ*_1_(*t* | *A, Z, X*) itself (rather than its logarithm) serves as the bridge covariate in the second stage. As in the multiplicative case, identification requires *β*_1*U*_ (*t*)*≠* 0 and *γ*_*Z*_(*t*) 0. Because the time-varying baseline 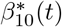 is collinear with the second-stage baseline, in estimation the covariate part 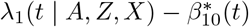 is used as the bridge (Algorithm 6).

##### Marginal survival and cumulative incidence

Let 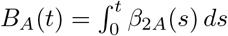 *ds* denote the cumulative causal excess hazard. Because the intervention contrast is additive and free of *U*, the *U*-conditional counterfactual survival factorizes,

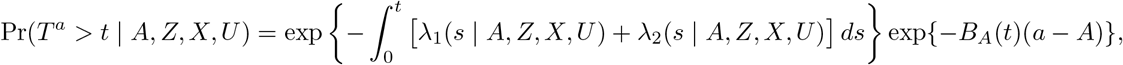

###### Algorithm 6

Proximal inference approach, additive hazards

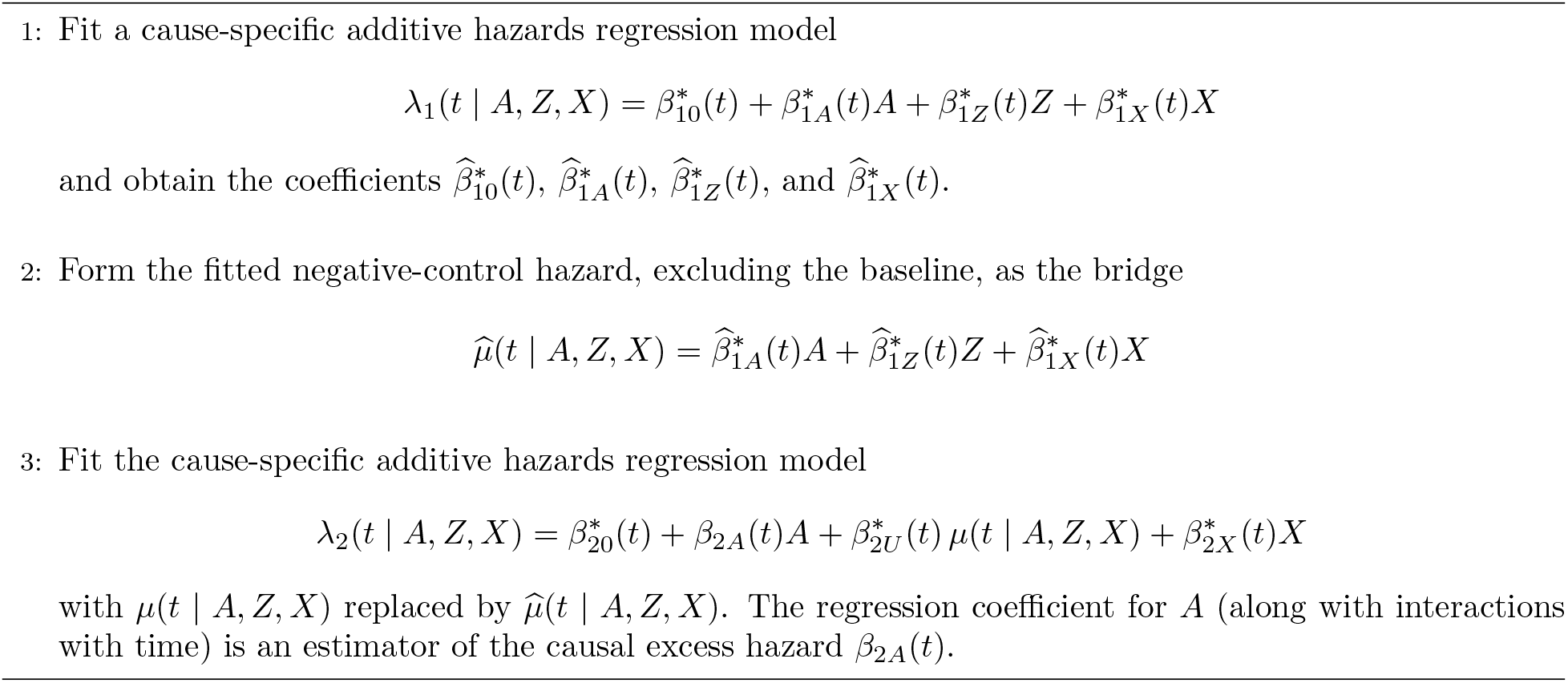

and the second factor, being independent of *U*, survives marginalization over *U:*

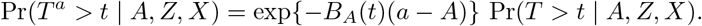

That is, the counterfactual conditional survival equals the observed conditional survival (which is identified nonparametrically) times a known scalar. The marginal counterfactual survival, cause-specific hazard, and cumulative incidence functions follow as

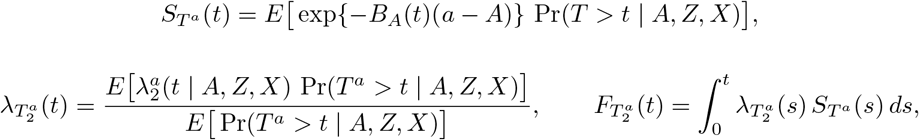

with 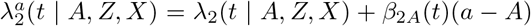 and *β*_2*A*_(*t*) identified by either the equi-confounding subtraction or the two-stage procedure above. The estimation steps are summarized in Algorithm 7. The cost is that additive hazard models are not constrained to [0, 1] and may require truncation in finite samples. In practice, Li, Linderman, Shi, and Tchetgen [3] suggest replacing *S*^*a*^(*t*) with

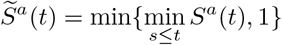

as a post hoc fix.

###### Algorithm 7

Marginal survival and cumulative incidence, additive hazards

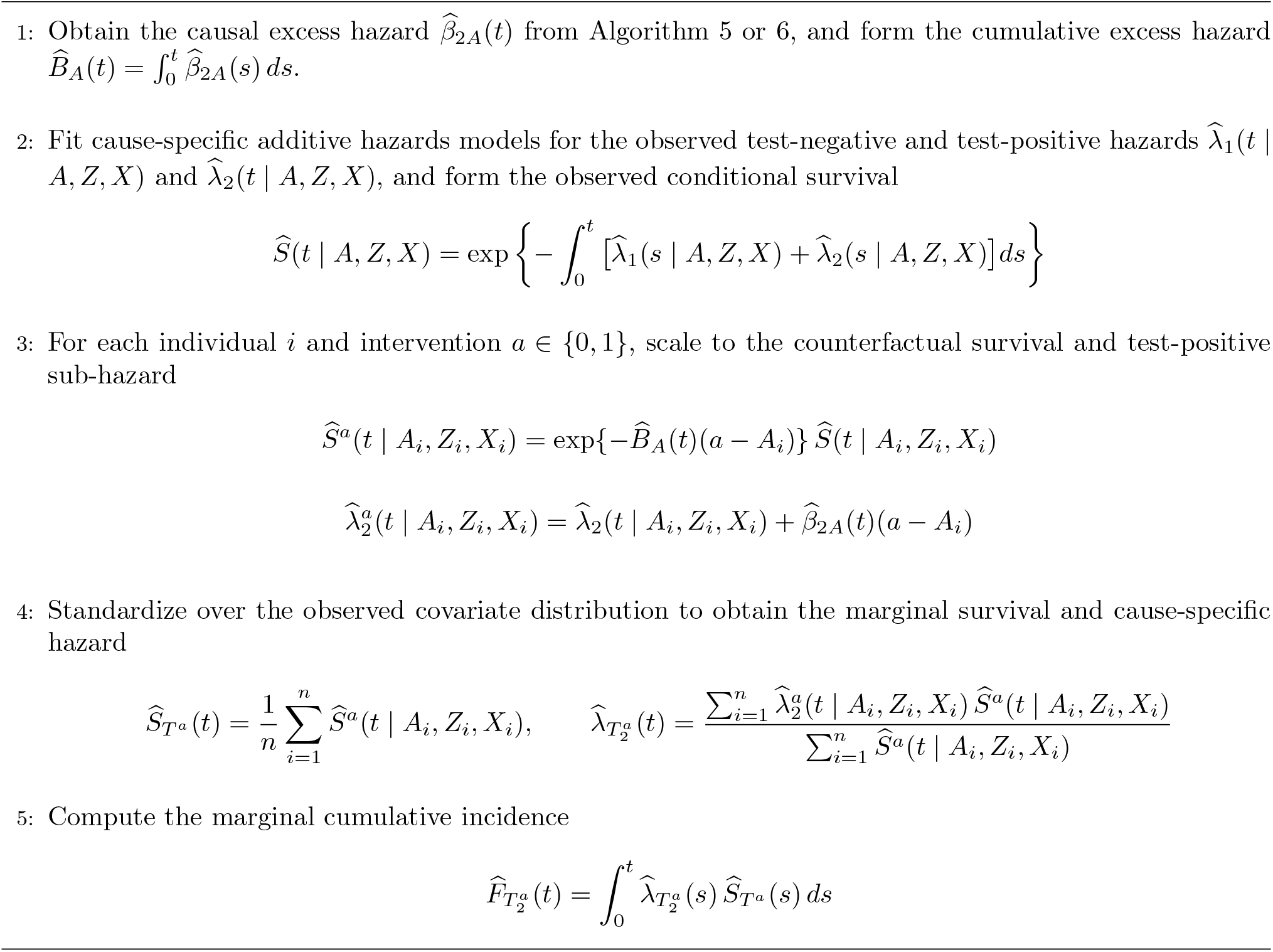

We compare results obtained from the multiplicative specifications for the EQC and PCI approaches presented in the main text to their additive analogs described here. Substituting assumption A6^*†*^ for A6 and re-specifying additive models results in protective VE of larger magnitude under the additive formulation of the EQC approach, owing to the high underlying risk of the NCO relative to the outcome of interest (NCO risks presented in Figure S10). Specification of additive models under the PCI approach yields similar results to the multiplicative formulation, which is expected given that the bridge coefficient in the second stage model is freely estimated, unlike the EQC correction which is subtracted directly.

**Figure S1:**
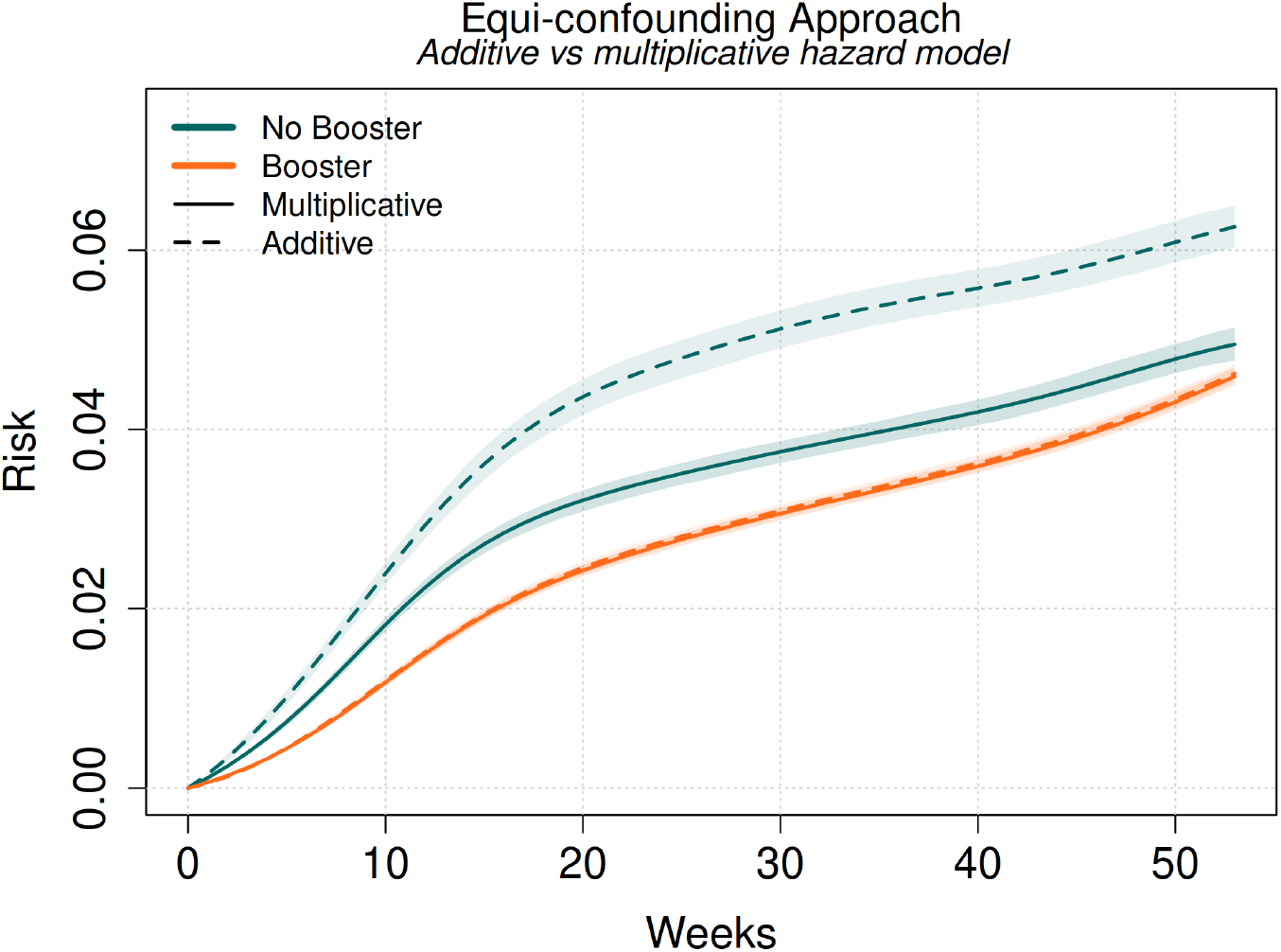
Cumulative incidence curves from pooled logistic models under EQC approach, comparing multiplicative to additive formulations.

**Figure S2:**
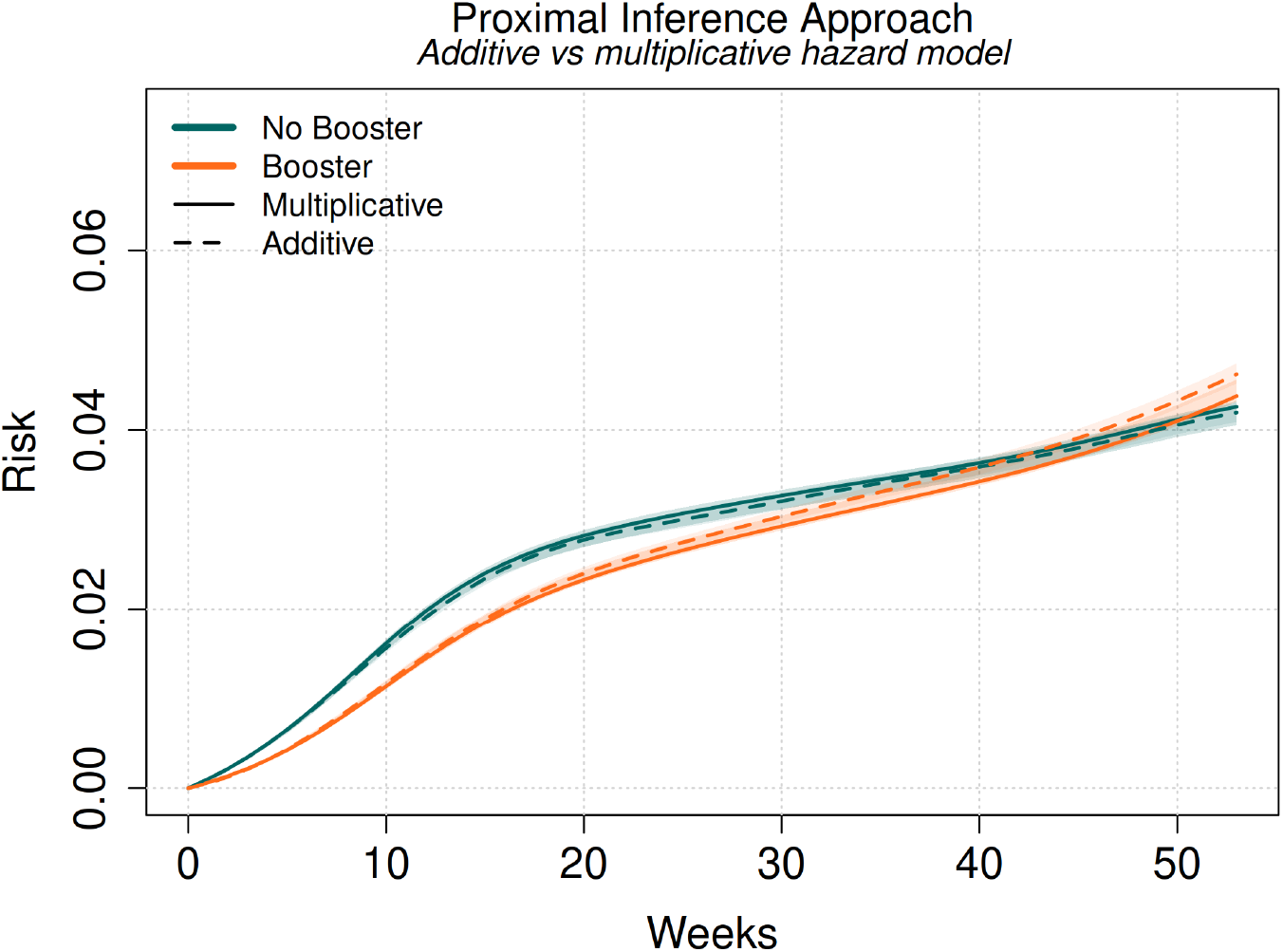
Cumulative incidence curves from pooled logistic models under PCI approach, comparing multiplicative to additive formulations.

#### S1.7 Net-risk of positive test (non-competing)

In the section above, testing negative and testing positive are treated as competing events, the estimand is the cumulative incidence of the *first* test being positive. However, often the scientific target is instead the effect of vaccination on test-positive illness itself (as a negative test does not preclude a later positive test). As long as data on repeat testing practices are available, we can modify the approaches above to target the *net-risk* of testing positive. Here we present an alternative set up in which testing negative no longer censors the test-positive process, and show that (i) the identification results carry through for the alternative estimand under weaker censoring assumptions, and (ii) the cumulative incidence is recovered by a Kaplan–Meier-type rather than an Aalen–Johansen estimator.

##### Setup and estimand

Let *T*_2_ denote the time to the first positive test and take as the target causal estimand the net cumulative incidence 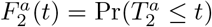 under an intervention setting *A* = *a*. Define the net test-positive hazard on the risk set *{T*_2_ *> t}* (“not yet positive”),

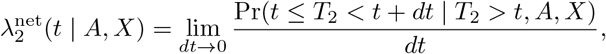

which differs from the cause-specific hazard of the competing-risks encoding in that an intervening negative test does not remove the individual from the risk set. Genuine terminal events (death, disenrollment, loss to follow up, administrative censoring) are included via right-censoring process represented by *C*.

##### Two new options for constructing negative-control

To be a valid NCO, in addition to being unaffected by focal vaccination the test-negative event process must reflect the structure of unmeasured residual confounding. Under equi-confounding, the mirroring should be exact on the specified scale (additive vs. multiplicative). Under the proximal inference set up, the test-negative event process need only be *U*-”relevant” —i.e. associated with source of unmeasured confounding. However, additional restrictions on the data generation process are required for the bridge to be strictly estimable using linear models.

Because the purpose of modeling the test negative event process is bias correction for the target outcome, rather than inference about testing negative specifically, multiple definitions are possible. The competing risk formulation in previous section represents one such option (retained as option C below). Here we discuss two more: the net time to first negative test (untruncated by a positive test) and the recurrent negative test intensity on the positive risk set.

- **Option A (first-negative time)**. Let *T*_1_ be the time to the first negative test (positives non-truncating) and

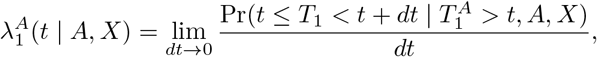

a single-event hazard on the risk set 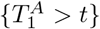, with survival interpretation 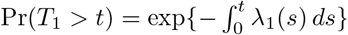
- **Option B (negative-test intensity on the positive risk set)**. Let *N*_1_(*t*) count negative tests and

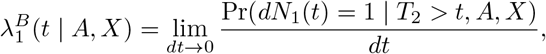

a recurrent-event intensity evaluated on the same risk set *{T*_2_ *> t}* as 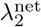; an individual contributes a negative event in every interval one occurs and remains at risk until the first positive.
- **Option C (competing risks)**. As before, let *T* = min(*T*_1_, *T*_2_) be the time to first test (positive or negative) and *δ* be the cause of the testing event such that *δ* = *j* if *T* = *T*_*j*_ for *j* = 1, 2

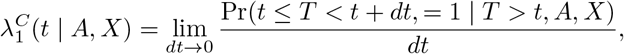

a competing event (with *T*_2_) evaluated on the risk set *{T > t}*; follow up is truncated after first test regardless of whether it is positive or negative.

As we show below, each option requires different conditions on the shared *U*, with option B being the easiest to derive from a single shared structural model. More practically, each negative control definition has different data requirements. Option A requires follow-up that continues past a positive test to capture subsequent negative tests. Option B requires the full negative-test history up to *T*_2_. Option B also implies that subjects can repeatedly test negative over the follow up period. Finally, Option C only requires the first test result (whether positive or negative) be recorded for each individual.

##### Assumptions

Consistency, exchangeability, and positivity (A1–A3) are now expressed for the target outcome *T*_2_:

(A1^*‡*^) *Consistency of potential outcomes*. For all individuals *i*, we have 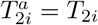 when *A*_*i*_ = *a*.

(A2^*‡*^) *Exchangeability conditional on* (*U, X*). That is, 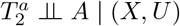.

(A3^*‡*^) *Positivity*. For all covariate patterns in support of *X* and *U*, we have Pr(*A* = *a*|*X, U*) *>* 0. The independent-censoring condition A4 is applied separately to each testing process,

(A4^*‡*^) *Independent censoring conditional on* (*A, X, Z*). That is, *C ╨ T*_2_ | (*A, Z, X*) and *C ╨ T*_1_ | (*A, Z, X*). The negative-control (A5) and equi-confounding (A6) conditions—or, for the proximal approach, the valid-NCE condition A6^*\**^—are re-stated on the net hazards, as detailed below.

(A5^*‡*^) *Testing negative is a valid NCO*. That is, 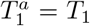 almost surely, 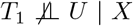, and *A ╨ T*_1_ | (*X, U*).

Equivalently, treatment has no causal effect on the test-negative event time, implying for all *t* that

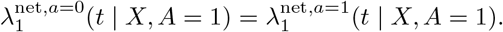

(A6^*‡*^) *Equi-confounding on multiplicative scale*. Residual confounding after conditioning on *X* is equivalent for test-negative and test-positive outcomes on the relative scale

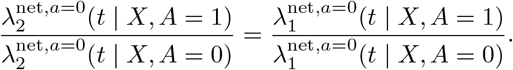

Alternatively for PCI approach we define

(A6^*‡\**^) *Z is a valid NCE*. That is, 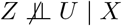, and *Z ╨* (*T*_1_, *T*_2_) | (*X, U*).

##### Equi-confounding

Write the net test-negative (NCO) hazard as 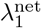 (two concrete options were given above). Equi-confounding on the net hazards scale,

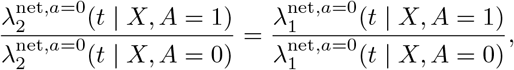

together with consistency and the NCO condition 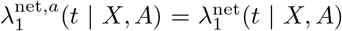, yields the corrected net hazard

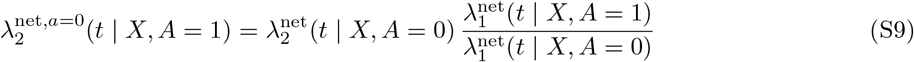

by the same algebra as in Section S1.3 (the additive analogs follow identically, replacing the ratio by the hazard difference). Equation S9 is exact whenever the NCO and equi-confounding conditions hold for the hazard function specified, i.e. Option B equates residual confounding between two hazards sharing the risk set *{T*_2_ *> t}*, whereas Option A equates it across the different risk sets *{T*_2_ *> t}* and 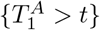, and Option C equates it. However, not all definitions of the hazard produce equally justifiable equi-confounding structures. Returning to our structural model in Section S1.2 with *β*_1*U*_ (*t*) = *β*_2*U*_ (*t*) = *β*_*U*_ (*t*), marginalizing over the survivor-conditional law of *U* gives

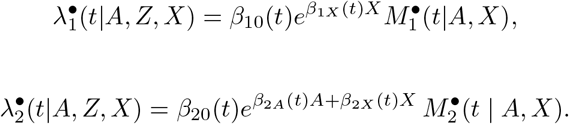

with 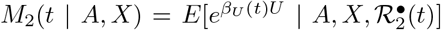 and 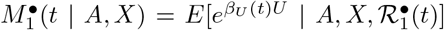 over the corresponding risk sets 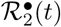 and 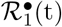, with *•∈ {A, B, C}*. The corrected hazard S9 under equi-confounding recovers the causal contrast of interest exp*{β*_2*A*_(*t*)} when

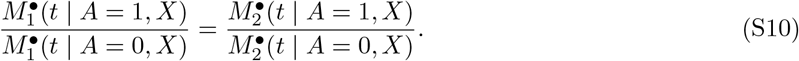

Under Option B, 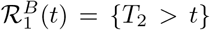 and 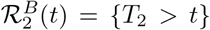 give 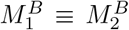 as functions, so S10 holds identically and the structural model with *β*_1*U*_ (*t*) = *β*_2*U*_ (*t*) = *β*_*U*_ (*t*) *implies* the required equi-confounding. Likewise, under Option C, 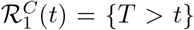 and 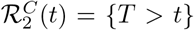 give 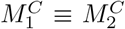 as functions and S10 holds identically. However, under Option A, 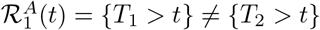, so 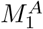 and *M*_2_ are expectations over different survivor sets, and the residual bias in the corrected hazard under equi-confounding is

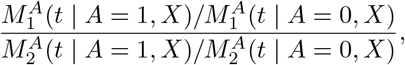

which equals unity in the limits (i) *t →* 0 (both risk sets equal the full cohort, so the bias is zero at the origin and can only accumulate through differential survivor selection); (ii) both test events rare over [0, *t*] (negligible depletion of either risk set); (iii) *U ╨ A* | *X* within each risk set, or *β*_*U*_*≡* 0 (no residual confounding to correct); and (iv) 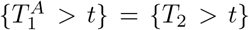 almost surely. Outside these limits, Option A’s cross-risk-set equi-confounding cannot by defined via the structural model, and instead must be assumed as a primitive.

Estimation under the net-risk outcome is the same as Algorithm 1 for multiplicative models with the net-risk hazard definitions 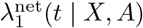 and 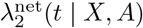 over risk sets A or B replacing the cause-specific hazards. Under option A, both hazards can be estimated using (non-truncated) Cox models of time to first negative and time to first positive test. Under option B, the recurrent test-negative intensity can be modeled using an Andersen-Gill model [4] which generalizes Cox model to recurrent events under counting process framework.

##### Marginal survival and cumulative incidence

Because a negative test is no longer a competing event, the counterfactual survival and cumulative incidence can be estimated via standard Kaplan-Meier methods based on the net hazard. For instance, the conditional survival among the treated is

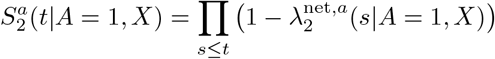

where for *a* = 0 we have

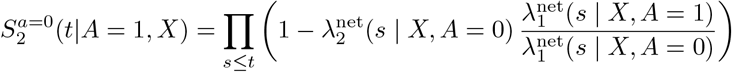

under equi-confounding, and *a* = 1 is simply the observed conditional survival (i.e., replace 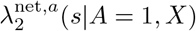 with ovserved 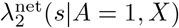).

Likewise, the conditional cumulative incidence is simply

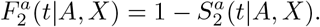

Relative to the competing-risks (Aalen–Johansen) cumulative incidence of a first positive test, 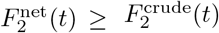, since removing the competing decrement can only increase cumulative risk, with equality only when negative tests are absent. Standardization over the covariate distribution is trivial once the conditional counterfactual elements are identified. For completeness, in Algorithm 8 we outline the full steps to recover the marginal counterfactual cumulative incidence and survival.

##### Proximal inference

The proximal approach of Section S1.5 can also be extended to target the net risk of testing positive. Re-stating the location-shift model of Section S1.5 on the net risk set,

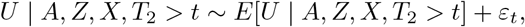

and the valid-NCE condition A6^*\**^, the two-stage construction is largely unchanged: (1) fit the net test-negative hazard 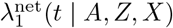 and form the bridge (its logarithm in the multiplicative case, or 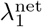 itself in the additive case of Section S1.6); (2) regress the net test-positive hazard on the bridge together with (*A, X*). The coefficient on *A* identifies *β*_2*A*_(*t*).

The counterfactual net hazard follows by the same switching relation,

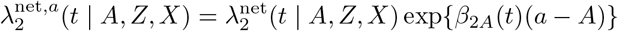

###### Algorithm 8

Net-risk marginal survival and cumulative incidence, multiplicative equi-confounding

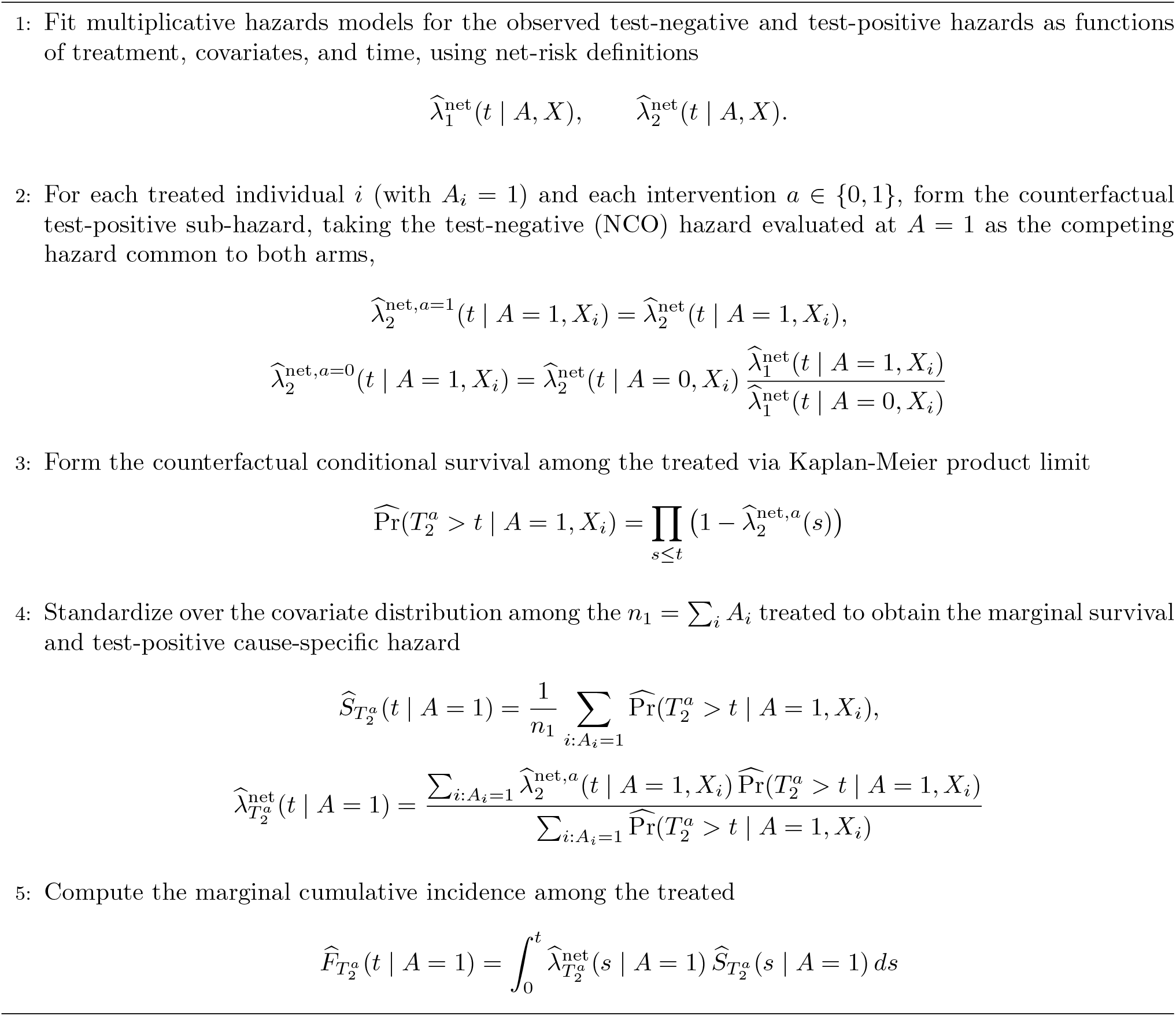

for multiplicative structural model or

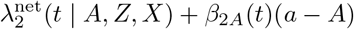

for the additive structural model.

Unlike equi-confounding, the proximal approach depends crucially on the location-shift structural model (at least when using simple regression) so the survivor-conditioning consistency between the two stages is essential. The bridge inversion solves for 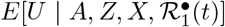 from the stage-one test-negative hazard and substitutes it into the stage-two positive hazard, which conditions on *{T*_2_ *> t}*; this substitution is valid only when the two stages condition on the same survivor set. Option B, whose test negative intensity is evaluated on 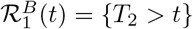, satisfies this automatically and yields the exact results of Section S1.5 with *T > t* replaced by *T*_2_ *> t*. Option A, whose NCO hazard conditions on 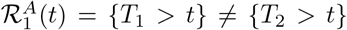, incurs the same survivor-set mismatch described above for equi-confounding, now in the bridge inversion; it is therefore the less natural choice for the proximal approach, and we recommend Option B whenever the recurrent negative-test history is available.

#### S1.8 Net risk results

Results from the analytic cohort in this subsection are from the net-risk formulation (S1.7) under Option B in which the negative test process is treated as a recurrent event. Both equi-confounding and proximal inference approaches are reported.

##### Cox model

The equi-confounding approach to net-risk estimation yields a HR of 0.87 (0.83, 0.90) and the proximal inference approach yields an HR of 0.80 (0.72, 0.87) (Table S1). Both estimates indicate slightly higher protection conferred by bivalent booster against test positive (net-risk) compared to first test positive (competing risks, Table 1).

**Table S1:** Effectiveness of the bivalent COVID-19 booster against net risk of positive SARS-CoV-2 test over 52 weeks, with negative-control exposure diagnostics (per-protocol).

| Approach | Net Risk: Bivalent<br>Booster |  | Net Risk: Prior<br>Year Flu Vax<br>(NCE) |  | VE (%) | 95% CI |
| --- | --- | --- | --- | --- | --- | --- |
|  | HR | 95% CI | HR | 95% CI |  |  |
| EQC | 0.87 | (0.83, 0.90) | 1.06 | (1.02, 1.10) | 13.3 | (9.9, 16.8) |
| PCI | 0.80 | (0.72, 0.87) | — | — | 20.2 | (13.5, 27.7) |

*Note.* Entries are hazard ratios (HR) with 95% confidence intervals from Cox proportional hazards models for net risk of positive test, i.e. the risk when individuals are not censored at negative test times, in contrast to the first-test-positive estimand under competing risks in Table 1. Vaccine effectiveness is $VE = 100 \times (1 - HR)$ computed from the net-risk HR and represents average relative vaccine effectiveness over 52 weeks. Analyses are per-protocol and adjusted for the baseline covariates in Table S8. Analytic cohort and matched pairs are identical to the main text analyses. *Net Risk: Bivalent Booster* is the target association from which VE is derived. *Net Risk: Prior Year Flu Vax (NCE)* is the association between the negative-control exposure (prior-season influenza vaccination) and net risk, expected to be null ( $HR = 1$ ) absent residual confounding; departures from 1 signal uncontrolled confounding. “—” denotes the NCE association under PCI, which is not separately estimated because the NCE and NCO together form the proximal bridge.
*Abbreviations:* EQC, equi-confounding; PCI, proximal causal inference; CI, confidence interval; HR, hazard ratio; VE, vaccine effectiveness; NCE, negative control exposure; NCO, negative control outcome.

##### Pooled logistic model

Net-risk curves lie above competing-events curves for both arms in both approaches, as expected in the absence of censoring at negative test times. The EQC and PCI approaches yield more protective estimates of VE against net-risk of medically attended COVID-19 (Table S2, Table S3) on both the multiplicative and additive risk scales compared to the competing risks formulation (Table S12, Table S13).

**Figure S3:**
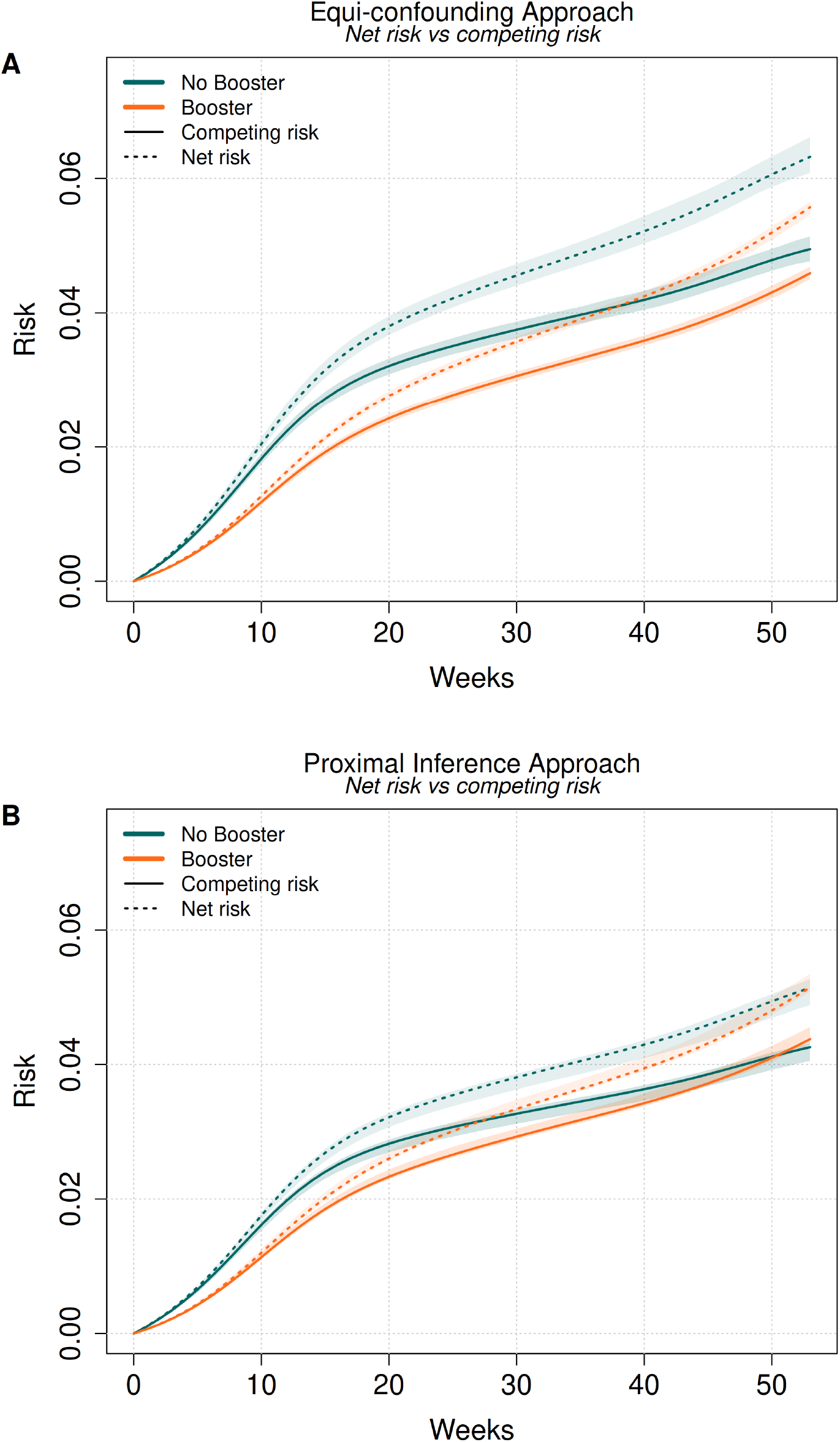
Net-risk and competing-events cumulative incidence of testing positive by arm, per-protocol analysis. A) Equi-confounding approach. B) Proximal inference approach. Solid lines are competing-events estimates and dotted lines are net-risk estimates.

**Table S2:** Pooled logistic results by week, net risk (EQC)

| Time (weeks) | Risk Ratio (95% CI) | Risk Difference (95% CI) |
| --- | --- | --- |
| 10 | 0.62 (0.59, 0.65) | -0.0077 (-0.0087, -0.0069) |
| 20 | 0.73 (0.70, 0.76) | -0.0104 (-0.0120, -0.0090) |
| 30 | 0.78 (0.75, 0.81) | -0.0099 (-0.0117, -0.0083) |
| 40 | 0.81 (0.78, 0.84) | -0.0097 (-0.0118, -0.0079) |
| 50 | 0.86 (0.82, 0.89) | -0.0086 (-0.0111, -0.0065) |

**Table S3:** Pooled logistic results by week, net risk (PCI)

| Time (weeks) | Risk Ratio (95% CI) | Risk Difference (95% CI) |
| --- | --- | --- |
| 10 | 0.68 (0.66, 0.74) | -0.0055 (-0.0061, -0.0044) |
| 20 | 0.81 (0.79, 0.87) | -0.0062 (-0.0068, -0.0040) |
| 30 | 0.88 (0.86, 0.95) | -0.0047 (-0.0053, -0.0018) |
| 40 | 0.92 (0.90, 1.00) | -0.0035 (-0.0044, -0.0002) |
| 50 | 0.97 (0.95, 1.05) | -0.0014 (-0.0024, 0.0026) |

##### Testing behavior

We note a greater deviation in the negative testing behavior between arms under the net-risk formulation relative to the competing risks formulation, as expected when individuals are not censored after their first negative test (Figure S4).

**Figure S4:**
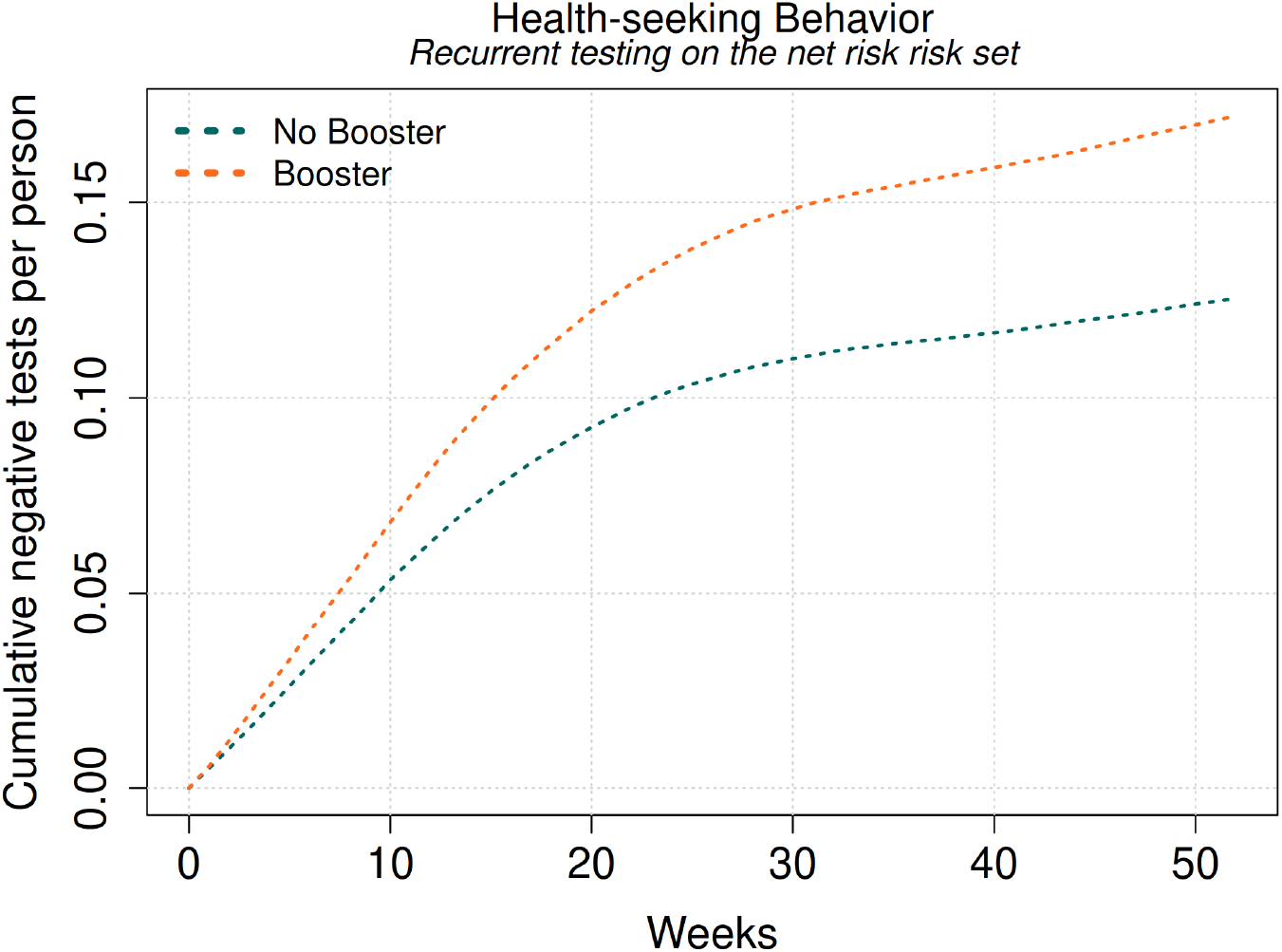
Cumulative negative tests per person by arm on the net-risk risk set, per-protocol analysis. Because negative tests are recurrent under Option B and do not remove an individual from the risk set, this quantity is a cumulative mean count rather than a cumulative incidence, and is not directly comparable to Figure S10.

#### S1.9 Simulation study

We conducted a simulation study to (i) confirm that the de-biasing approaches of Sections S1.3 and S1.5 recover the causal effect in the presence of known unmeasured confounding, (ii) demonstrate that the proximal inference (PCI) approach remains valid when the equi-confounding (EQC) assumption (Assumption A6) is violated, (iii) compare the competing-risks encoding with the two net-risk negative-control constructions of Section S1.7, and (iv) compare the pooled discrete-time estimator used in the main analysis against Cox proportional-hazards estimators, for both the causal log hazard-ratio and the cumulative incidence.

##### Data-generating process

We simulate in continuous time on a weekly grid over *τ* = 53 weeks for *N* = 10,000 individuals. Data are generated via the following process:

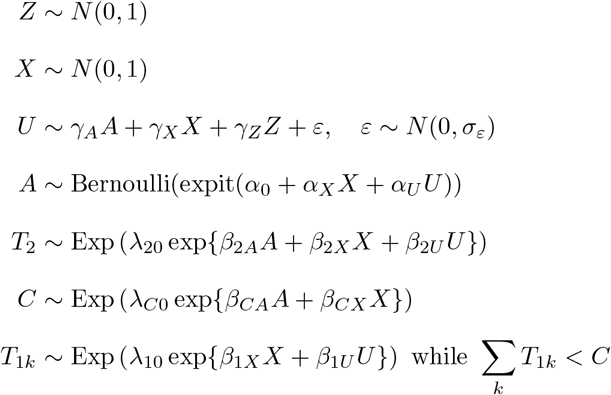

where *α*_*U*_ *≠* 0, so that *U* confounds treatment, *T*_2_ follows the proportional hazard, *T*_1*k*_ arise as a recurrent Poisson process that does not depend on *A* but is related to *U* (so testing negative is a valid NCO), and *Z* is related to *A* through *U* but not to *T*_1_ (so is a valid NCE). When *β*_1*U*_ = *β*_2*U*_ multiplicative equi-confounding holds exactly (e.g. via by the Option-B argument of Section S1.7). Right-censoring time *C* depends on (*A, X*) only. We set *β*_2*A*_ = −0.7 (the true causal log hazard-ratio), (*β*_2*X*_, *β*_1*X*_) = (0.3, 0.4), confounding parameters (*γ*_*A*_, *γ*_*Z*_, *γ*_*X*_, *σ*_*ε*_) = (0.5, 0.6, 0.3, 0.3), and baseline rates (*λ*_20_, *λ*_10_) = (0.0019, 0.0032) calibrated so that roughly 8% of individuals test positive and 18% test negative at least once over follow-up.

Two scenarios are generated:

- *Equi-confounding holds*: *β*_2*U*_ = *β*_1*U*_ = 0.8, so the equi-confounding assumption is satisfied exactly.
- *Equi-confounding violated* : *β*_2*U*_ = 1.0 *> β*_1*U*_ = 0.6, so the positive outcome is more heavily confounded, violating the equi-confounding assumption.

From each dataset we construct the three risk-set encodings: the *competing-risks* encoding (option C, the first test of either type is terminal, i.e. *T* = min(*T*_1_, *T*_2_)); *net-risk Option A* (time to first positive, with the first-negative time as a single-event NCO on its own risk set, i.e. *T* = *T*_2_ and *T*_1_ defined over 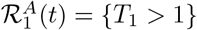 and *net-risk Option B* (time to first positive, with the recurrent negative-test intensity on the positive risk set as the NCO, i.e. *T* = *T*_2_ and *T*_1_ defined over 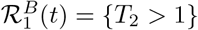.

##### Estimators

Within each risk-set encoding we estimate *β*_2*A*_ comparing three strategies:

- *Naive*: regression of the test-positive hazard on (*A, X*) with no negative-control de-biasing.
- *Equi-confounding* : the difference between the test-positive and test-negative treatment coefficient on the log-hazard scale.
- *Proximal inference*: a two-stage estimator in which in stage one the test-negative hazard is regressed on (*A, Z, X*) and the “bridge” covariate is formed from the fitted log-odds (Section S1.5), and in stage two the test-positive hazard is regressed on (*A, X*, bridge) with the *A* coefficient identifying *β*_2*A*_.

For each strategy, we also compare three estimators of the cause-specific hazard:

- *Pooled* : a pooled discrete-time logistic model with natural-spline time–treatment interactions (as in the main analysis)
- *Cox, scalar* : a proportional-hazards Cox model with baseline unspecified
- *Cox, time-varying* : a Cox model with piecewise time-varying treatment coefficients

Cumulative incidence among the treated (the ATT) is obtained by the Kaplan–Meier estimator for the net risk encodings and by the Aalen–Johansen estimator for the competing-risk encoding, contrasting the observed treated risk 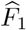 with the de-biased untreated risk 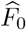.

##### Main results: pooled logistic model, competing-risks encoding

Table S4 and Figure S5 summarize recovery of *β*_2*A*_ over 200 replications for the pooled discrete-time logistic model on the competing-risks encoding. The naive estimator is severely confounded in both scenarios (bias +0.69 when equi-confounding holds, +0.89 when violated). When equi-confounding holds, both de-biasing strategies recover *β*_2*A*_ with negligible bias (EQC −0.01, PCI −0.02). When equi-confounding is violated, the EQC estimator is biased (+0.35), while PCI remains unbiased (−0.03), because it does not rely on this assumption. Both results are consistent with theory of Sections S1.3 and S1.5 above.

**Table S4:** Bias, standard error, and RMSE of the causal log hazard-ratio estimate (*β*_2*A*_ = −0.7) under three de-biasing strategies, by equi-confounding assumption status. All estimates use the pooled discrete-time logistic model on the competing-risks encoding. Results based on 200 replications with *N* = 4,000 per replication.

| Scenario | Strategy | Bias | SE | RMSE |
| --- | --- | --- | --- | --- |
| Equi-confounding holds | naive | 0.6894 | 0.1515 | 0.7058 |
|  | eqc | -0.0134 | 0.1736 | 0.1736 |
|  | pci | -0.0182 | 0.2130 | 0.2132 |
| Equi-confounding violated | naive | 0.8899 | 0.1322 | 0.8997 |
|  | eqc | 0.3522 | 0.1627 | 0.3878 |
|  | pci | -0.0262 | 0.2586 | 0.2593 |

**Figure S5:**
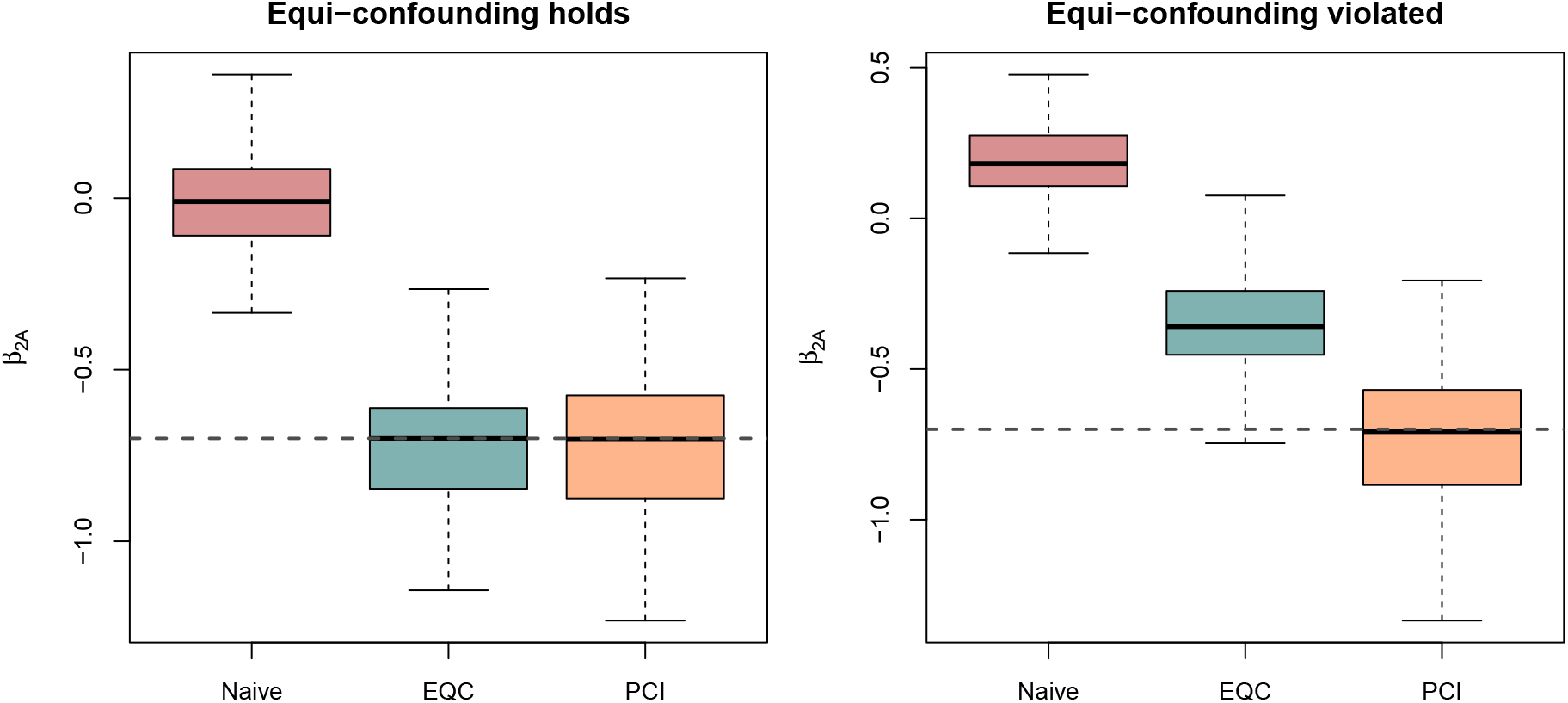
Sampling distributions of the estimated causal log hazard-ratio 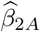 over 200 replications, by de-biasing strategy, when equi-confounding holds (left) and when it is violated (right); the dashed line marks the true value −0.70. Results use the pooled discrete-time logistic model on the competing-risks encoding. The naive estimator is severely confounded in both scenarios; the equi-confounding estimator is unbiased only when its assumption holds; the proximal-inference estimator is unbiased in both scenarios.

##### Robustness to the choice of cause-specific hazard estimator

Table S5 and Figure S6 compare the pooled logistic model against the Cox time-varying and Cox scalar estimators using the competing-risks encoding under equi-confounding violation. All three cause-specific hazard estimators produce essentially the same bias pattern for each de-biasing strategy. So the pooled

##### Robustness to the choice of risk-set encoding

Table S6 and Figure S7 compare the three risk-set encodings of Section S1.7—competing-risks, net-risk Option A, and net-risk Option B—using the pooled logistic model, for each de-biasing strategy and equi-confounding scenario. The bias pattern for each strategy is nearly identical across all three encodings: naive is severely confounded under all three, EQC is unbiased only when its assumption holds (with a small additional residual under Option A, consistent with the survivor-set mismatch identified in Section S1.7), and PCI remains near-unbiased regardless of encoding. Thus the choice between the competing-risks formulation used in the main analysis and either net-risk alternative does not affect the qualitative comparison between EQC and PCI.

**Table S5:** Cross-estimator robustness: bias, standard error, and RMSE for three cause-specific hazard estimators (pooled logistic, Cox proportional hazards, Cox time-varying), under three de-biasing strategies, in the scenario where equi-confounding assumption is violated. Results based on competing-risks encoding with 200 replications (*N* = 4,000 per rep). The pooled logistic model is used in the main analysis.

| Strategy | Estimator | Bias | SE | RMSE |
| --- | --- | --- | --- | --- |
| naive | pooled | 0.8899 | 0.1322 | 0.8997 |
| naive | cox_scalar | 0.8906 | 0.1286 | 0.8998 |
| naive | cox_tv | 0.8918 | 0.1312 | 0.9013 |
| eqc | pooled | 0.3522 | 0.1627 | 0.3878 |
| eqc | cox_scalar | 0.3546 | 0.1596 | 0.3887 |
| eqc | cox_tv | 0.3558 | 0.1617 | 0.3907 |
| pci | pooled | -0.0262 | 0.2586 | 0.2593 |
| pci | cox_scalar | -0.0228 | 0.2533 | 0.2537 |
| pci | cox_tv | -0.0183 | 0.2531 | 0.2531 |

##### Cumulative incidence curves

Figure S8 plots the sampling distribution of the de-biased untreated cumulative incidence curve 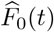 over the full follow-up period, for each strategy and scenario, alongside the true untreated curve *F*_0_(*t*) obtained analytically from the known data-generating process. The PCI-corrected curve tracks the true counterfactual risk trajectory closely in both scenarios, while the EQC-corrected curve tracks the truth only when equi-confounding holds and is displaced downward once it is violated, mirroring the bias pattern seen in the log hazard-ratio.

**Figure S6:**
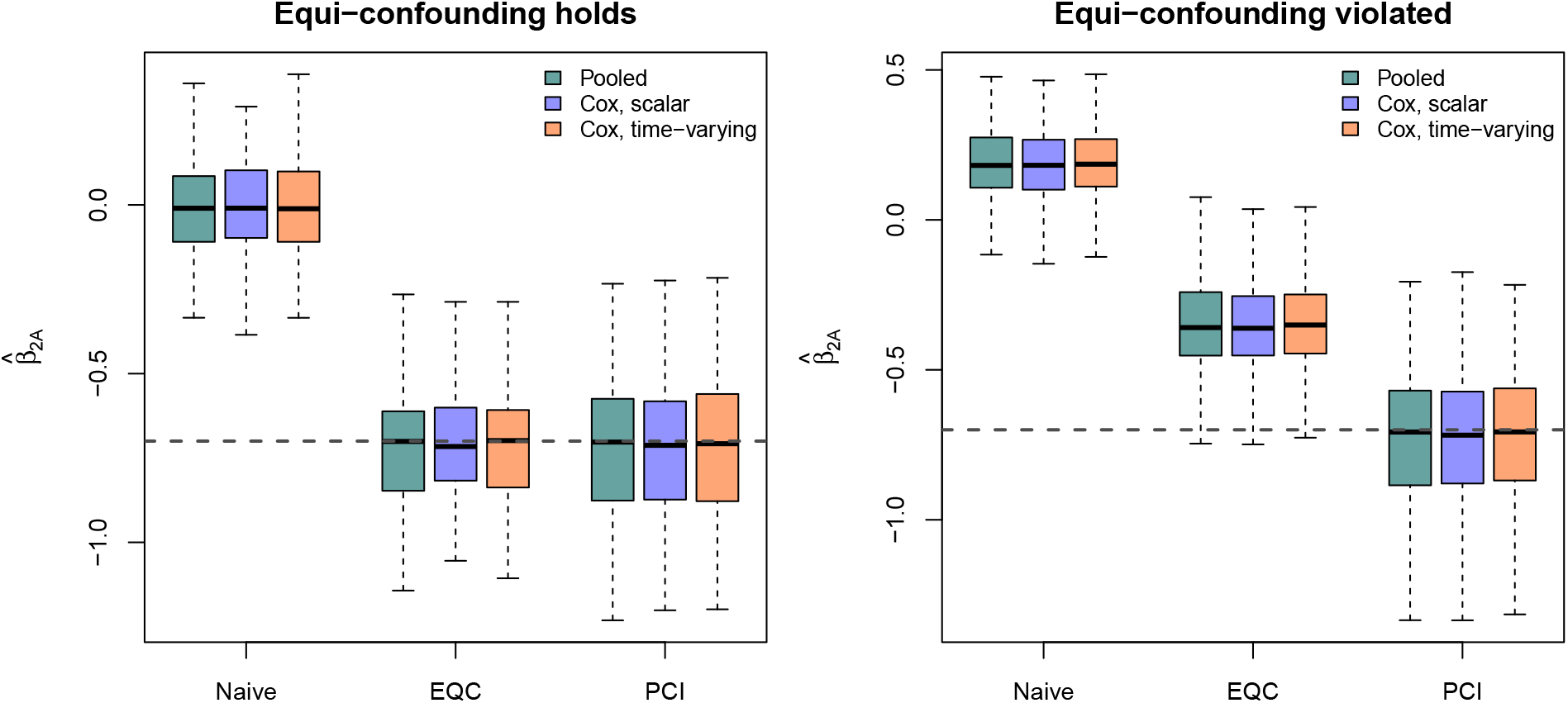
Sampling distributions of the estimated causal log hazard-ratio 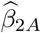 by de-biasing strategy and cause-specific hazard estimator (pooled logistic, Cox time-varying, Cox scalar), within the competing-risks encoding, under each equi-confounding scenario; the dashed line marks the true value −0.70. The three estimators agree closely within each strategy, indicating that the robustness of proximal inference to equi-confounding violation does not depend on the choice of hazard model.

**Table S6:** Robustness to the choice of risk-set encoding: bias, standard error, and RMSE of the causal log hazard-ratio estimate under three de-biasing strategies and three risk-set encodings (competing-risks, net-risk Option A, net-risk Option B), by equi-confounding assumption status. All estimates use the pooled discrete-time logistic model. Results based on 200 replications with *N* = 4,000 per replication.

| Scenario | Strategy | Encoding | Bias | SE | RMSE |
| --- | --- | --- | --- | --- | --- |
| Equi-confounding holds | Naive | Competing risk | 0.6894 | 0.1515 | 0.7058 |
|  |  | Net-risk, Option A | 0.7322 | 0.1302 | 0.7436 |
|  |  | Net-risk, Option B | 0.7322 | 0.1302 | 0.7436 |
|  | Equi-confounding | Competing risk | -0.0134 | 0.1736 | 0.1736 |
|  |  | Net-risk, Option A | 0.0313 | 0.1563 | 0.1590 |
|  |  | Net-risk, Option B | -0.0018 | 0.1546 | 0.1542 |
|  | Proximal (PCI) | Competing risk | -0.0182 | 0.2130 | 0.2132 |
|  |  | Net-risk, Option A | 0.0014 | 0.1929 | 0.1925 |
|  |  | Net-risk, Option B | -0.0057 | 0.1876 | 0.1872 |
| Equi-confounding violated | Naive | Competing risk | 0.8899 | 0.1322 | 0.8997 |
|  |  | Net-risk, Option A | 0.9103 | 0.1178 | 0.9178 |
|  |  | Net-risk, Option B | 0.9103 | 0.1178 | 0.9178 |
|  | Equi-confounding | Competing risk | 0.3522 | 0.1627 | 0.3878 |
|  |  | Net-risk, Option A | 0.3700 | 0.1489 | 0.3987 |
|  |  | Net-risk, Option B | 0.3625 | 0.1460 | 0.3907 |
|  | Proximal (PCI) | Competing risk | -0.0262 | 0.2586 | 0.2593 |
|  |  | Net-risk, Option A | -0.0086 | 0.2338 | 0.2334 |
|  |  | Net-risk, Option B | -0.0193 | 0.2342 | 0.2344 |

**Figure S7:**
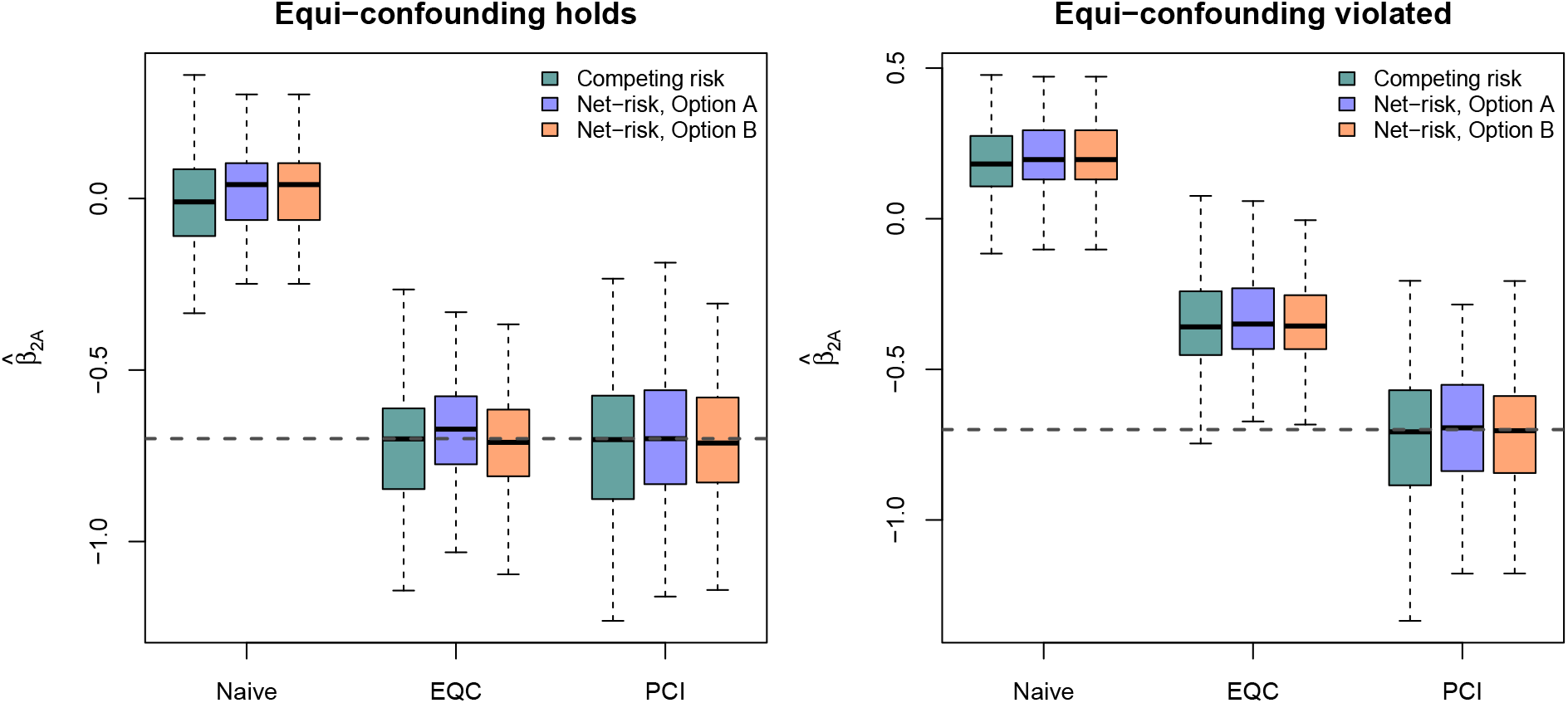
Sampling distributions of the estimated causal log hazard-ratio 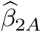 by de-biasing strategy and risk-set encoding (competing-risks, net-risk Option A, net-risk Option B), using the pooled discrete-time logistic model, under each equi-confounding scenario; the dashed line marks the true value −0.70. The three encodings agree closely within each strategy.

**Figure S8:**
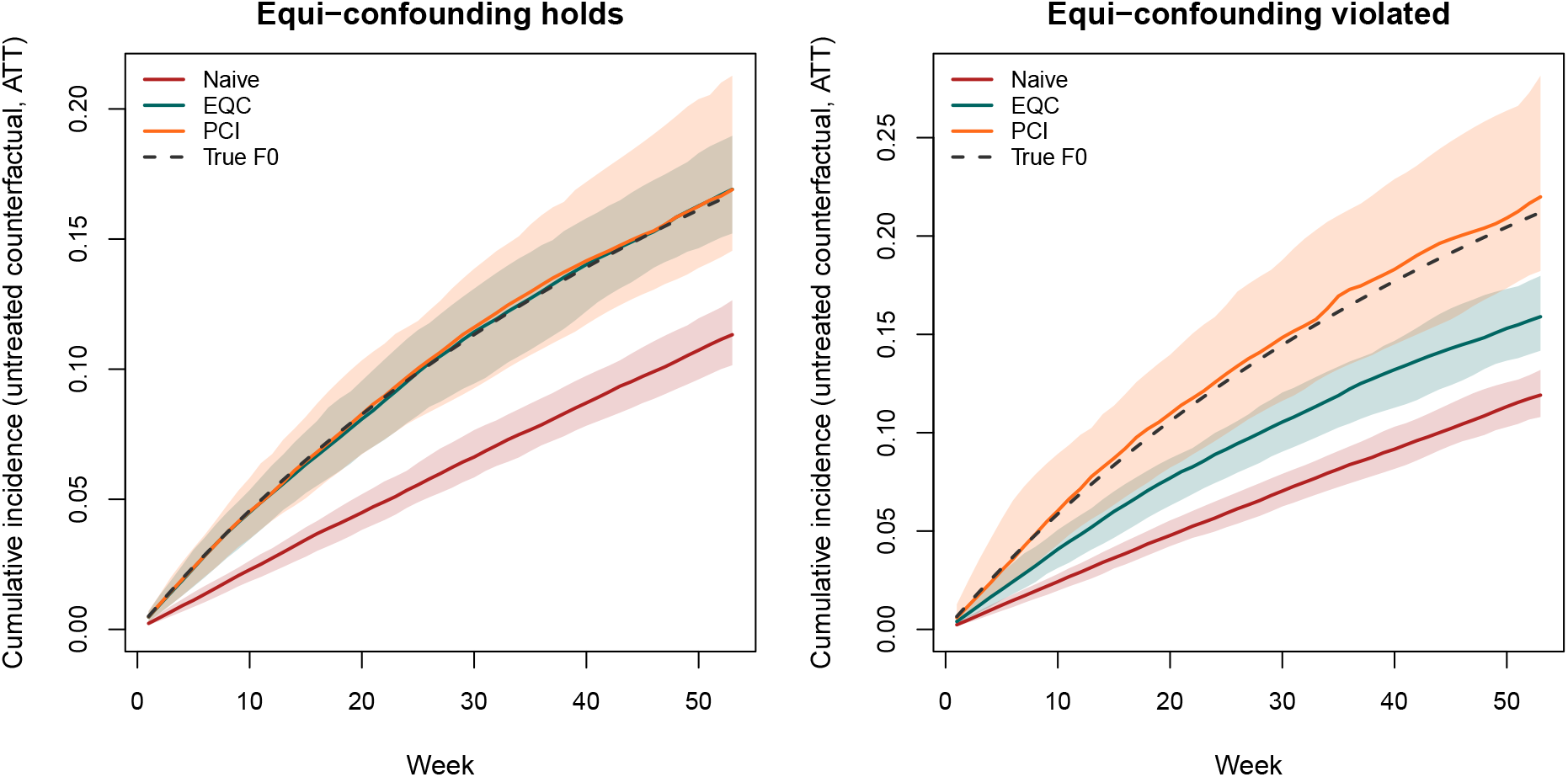
Sampling distributions of the de-biased untreated cumulative incidence curve 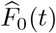 among the treated (ATT), by de-biasing strategy, over the full 53-week follow-up (competing-risks encoding, pooled logistic model). Solid lines show the median across replications and shaded ribbons the 10th–90th percentile band; the dashed grey line is the true untreated cumulative incidence *F*_0_(*t*) computed analytically from the data-generating process.

#### S1.10 Directed Acyclic Graphs

**Figure S9:**
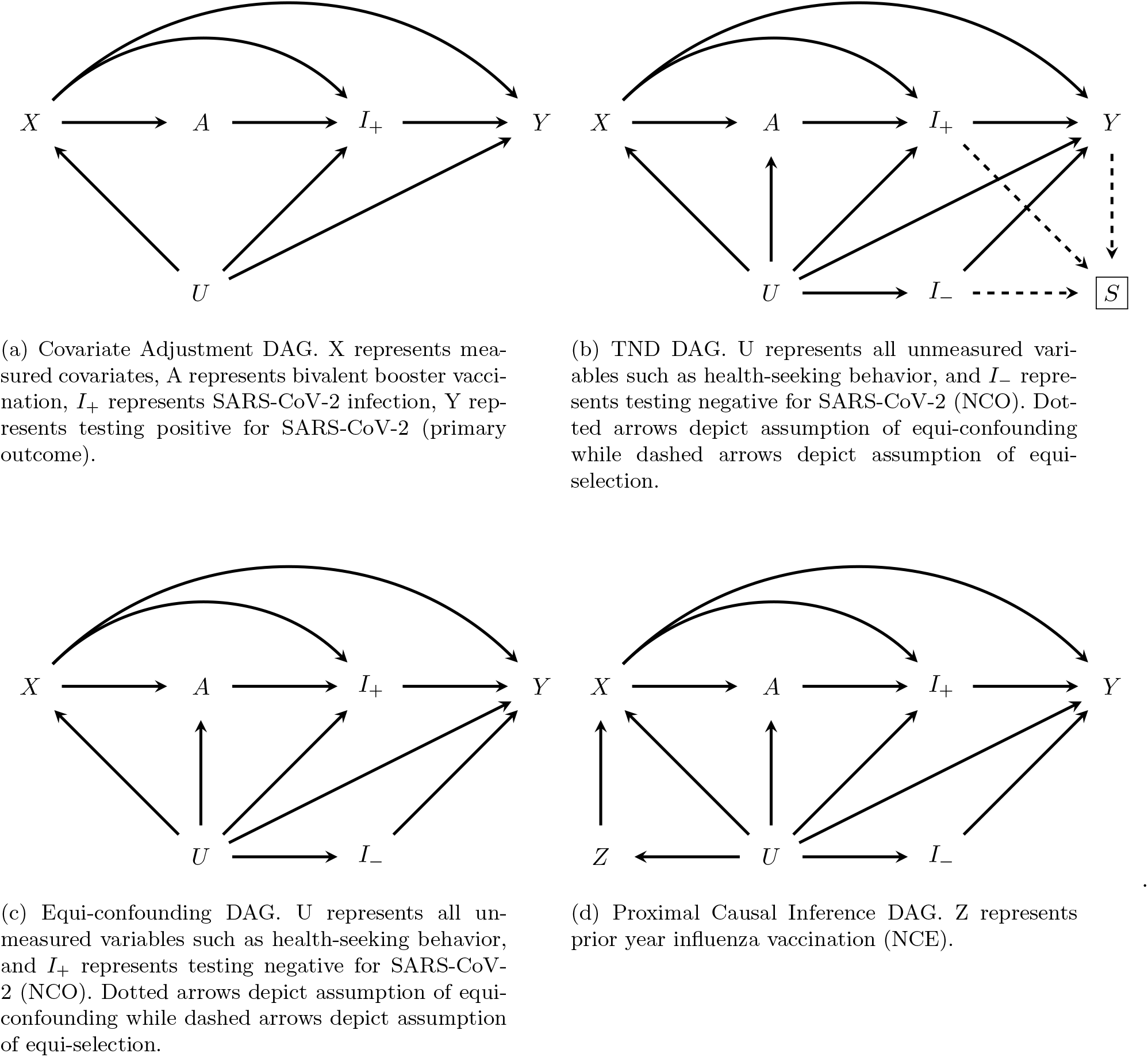
Directed acyclic graphs for estimation approaches

### S2 Additional Results

#### S2.1 Covariate Balance

**Table S7:** Covariate balance before matching.

|  | level | Booster | No Booster | SMD |
| --- | --- | --- | --- | --- |
| n |  | 189121 | 674832 |  |
| Age (mean (SD)) |  | 48.21 (16.50) | 46.45 (16.94) | 0.105 |
| BMI (mean (SD)) |  | 28.06 (6.52) | 28.52 (6.66) | 0.069 |
| NDI (mean (SD)) |  | -0.41 (0.79) | -0.28 (0.82) | 0.156 |
| Test count (mean (SD)) |  | 1.45 (2.03) | 1.39 (2.05) | 0.032 |
| Weeks last inf/vax (mean (SD)) |  | -37.91 (7.46) | -35.12 (7.29) | 0.378 |
| Sex (%) | F | 108275 (57.3) | 384033 (56.9) | 0.007 |
|  | M | 80846 (42.7) | 290799 (43.1) |  |
| Race (%) | White | 83222 (44.0) | 255639 (37.9) | 0.217 |
|  | Asian | 57417 (30.4) | 181197 (26.9) |  |
|  | Hispanic | 29248 (15.5) | 153276 (22.7) |  |
|  | Black | 9198 (4.9) | 43649 (6.5) |  |
|  | Unknown/ot | 10036 (5.3) | 41071 (6.1) |  |
| Service region (%) | Central valley | 13775 (7.3) | 74799 (11.1) | 0.165 |
|  | East bay | 56140 (29.7) | 186081 (27.6) |  |
|  | North bay | 10310 (5.5) | 39849 (5.9) |  |
|  | North valley | 46505 (24.6) | 171726 (25.4) |  |
|  | Peninsula | 31174 (16.5) | 86695 (12.8) |  |
|  | South bay | 31217 (16.5) | 115682 (17.1) |  |
| Prior year flu (%) | 0 | 60520 (32.0) | 362026 (53.6) | 0.448 |
|  | 1 | 128601 (68.0) | 312806 (46.4) |  |
| Prior COVID-19 infection (%) | 0 | 158990 (84.1) | 551698 (81.8) | 0.062 |
|  | 1 | 30131 (15.9) | 123134 (18.2) |  |
| Charlson (%) | 0 | 135236 (71.5) | 485002 (71.9) | 0.017 |
|  | 1 | 27048 (14.3) | 97740 (14.5) |  |
|  | 2 | 11624 (6.1) | 40430 (6.0) |  |
|  | 3+ | 15213 (8.0) | 51660 (7.7) |  |

**Table S8:**
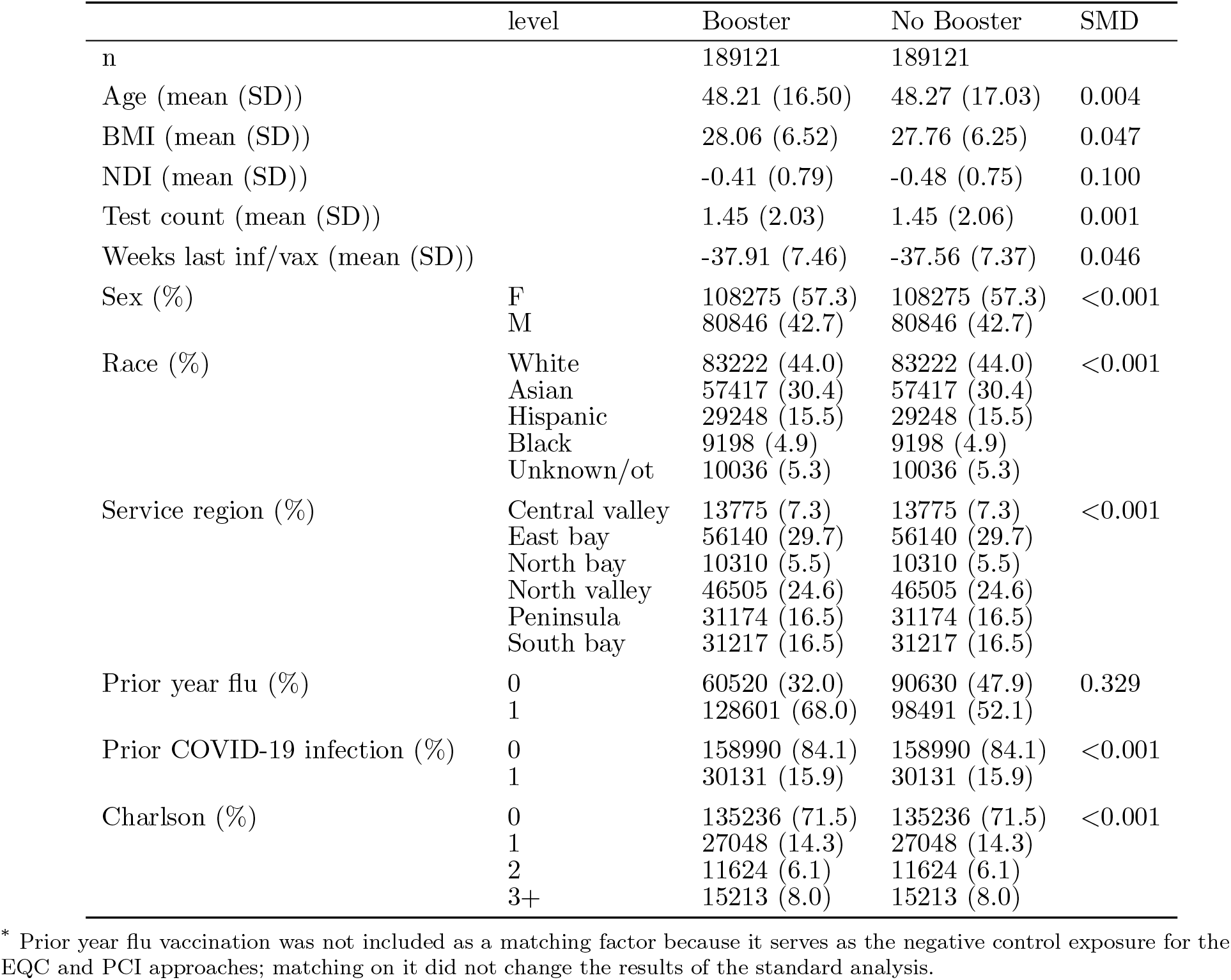
Covariate balance after matching.

|  | level | Booster | No Booster | SMD |
| --- | --- | --- | --- | --- |
| n |  | 189121 | 189121 |  |
| Age (mean (SD)) |  | 48.21 (16.50) | 48.27 (17.03) | 0.004 |
| BMI (mean (SD)) |  | 28.06 (6.52) | 27.76 (6.25) | 0.047 |
| NDI (mean (SD)) |  | -0.41 (0.79) | -0.48 (0.75) | 0.100 |
| Test count (mean (SD)) |  | 1.45 (2.03) | 1.45 (2.06) | 0.001 |
| Weeks last inf/vax (mean (SD)) |  | -37.91 (7.46) | -37.56 (7.37) | 0.046 |
| Sex (%) | F | 108275 (57.3) | 108275 (57.3) | <0.001 |
|  | M | 80846 (42.7) | 80846 (42.7) |  |
| Race (%) | White | 83222 (44.0) | 83222 (44.0) | <0.001 |
|  | Asian | 57417 (30.4) | 57417 (30.4) |  |
|  | Hispanic | 29248 (15.5) | 29248 (15.5) |  |
|  | Black | 9198 (4.9) | 9198 (4.9) |  |
|  | Unknown/ot | 10036 (5.3) | 10036 (5.3) |  |
| Service region (%) | Central valley | 13775 (7.3) | 13775 (7.3) | <0.001 |
|  | East bay | 56140 (29.7) | 56140 (29.7) |  |
|  | North bay | 10310 (5.5) | 10310 (5.5) |  |
|  | North valley | 46505 (24.6) | 46505 (24.6) |  |
|  | Peninsula | 31174 (16.5) | 31174 (16.5) |  |
|  | South bay | 31217 (16.5) | 31217 (16.5) |  |
| Prior year flu (%) | 0 | 60520 (32.0) | 90630 (47.9) | 0.329 |
|  | 1 | 128601 (68.0) | 98491 (52.1) |  |
| Prior COVID-19 infection (%) | 0 | 158990 (84.1) | 158990 (84.1) | <0.001 |
|  | 1 | 30131 (15.9) | 30131 (15.9) |  |
| Charlson (%) | 0 | 135236 (71.5) | 135236 (71.5) | <0.001 |
|  | 1 | 27048 (14.3) | 27048 (14.3) |  |
|  | 2 | 11624 (6.1) | 11624 (6.1) |  |
|  | 3+ | 15213 (8.0) | 15213 (8.0) |  |

\* Prior year flu vaccination was not included as a matching factor because it serves as the negative control exposure for the EQC and PCI approaches; matching on it did not change the results of the standard analysis.

**Table S9:** Covariate balance among individuals event-free and uncensored through week 52 (ITT)

|  | level | Booster | No Booster | SMD |
| --- | --- | --- | --- | --- |
| n |  | 145649 | 147913 |  |
| Age (mean (SD)) |  | 48.76 (16.64) | 48.57 (17.03) | 0.011 |
| BMI (mean (SD)) |  | 27.98 (6.46) | 27.68 (6.17) | 0.047 |
| NDI (mean (SD)) |  | -0.40 (0.79) | -0.48 (0.75) | 0.104 |
| Test count (mean (SD)) |  | 1.29 (1.84) | 1.31 (1.90) | 0.012 |
| Weeks last inf/vax (mean (SD)) |  | -37.90 (7.39) | -37.56 (7.30) | 0.046 |
| Sex (%) | F | 82075 (56.4) | 83527 (56.5) | 0.002 |
|  | M | 63574 (43.6) | 64386 (43.5) |  |
| Race (%) | White | 64129 (44.0) | 64995 (43.9) | 0.002 |
|  | Asian | 44535 (30.6) | 45349 (30.7) |  |
|  | Hispanic | 22296 (15.3) | 22675 (15.3) |  |
|  | Black | 6978 (4.8) | 7053 (4.8) |  |
|  | Unknown/ot | 7711 (5.3) | 7841 (5.3) |  |
| Service region (%) | Central valley | 11068 (7.6) | 11054 (7.5) | 0.012 |
|  | East bay | 42746 (29.3) | 43780 (29.6) |  |
|  | North bay | 8024 (5.5) | 8210 (5.6) |  |
|  | North valley | 36695 (25.2) | 36752 (24.8) |  |
|  | Peninsula | 23415 (16.1) | 24143 (16.3) |  |
|  | South bay | 23701 (16.3) | 23974 (16.2) |  |
| Prior year flu (%) | 0 | 47157 (32.4) | 71765 (48.5) | 0.333 |
|  | 1 | 98492 (67.6) | 76148 (51.5) |  |
| Prior COVID-19 infection (%) | 0 | 123268 (84.6) | 125162 (84.6) | <0.001 |
|  | 1 | 22381 (15.4) | 22751 (15.4) |  |
| Charlson (%) | 0 | 104419 (71.7) | 106711 (72.1) | 0.013 |
|  | 1 | 20571 (14.1) | 20862 (14.1) |  |
|  | 2 | 9108 (6.3) | 9018 (6.1) |  |
|  | 3+ | 11551 (7.9) | 11322 (7.7) |  |

**Table S10:** Covariate balance among individuals event-free and uncensored through week 52 (PP)

|  | level | Booster | No Booster | SMD |
| --- | --- | --- | --- | --- |
| n |  | 118409 | 93401 |  |
| Age (mean (SD)) |  | 47.70 (16.18) | 48.08 (17.03) | 0.023 |
| BMI (mean (SD)) |  | 27.98 (6.46) | 27.84 (6.18) | 0.022 |
| NDI (mean (SD)) |  | -0.41 (0.79) | -0.44 (0.76) | 0.049 |
| Test count (mean (SD)) |  | 1.30 (1.85) | 1.28 (1.89) | 0.014 |
| Weeks last inf/vax (mean (SD)) |  | -37.84 (7.34) | -36.73 (7.30) | 0.151 |
| Sex (%) | F | 67030 (56.6) | 52617 (56.3) | 0.006 |
|  | M | 51379 (43.4) | 40784 (43.7) |  |
| Race (%) | White | 51333 (43.4) | 40481 (43.3) | 0.056 |
|  | Asian | 36707 (31.0) | 27036 (28.9) |  |
|  | Hispanic | 18311 (15.5) | 15874 (17.0) |  |
|  | Black | 5623 (4.7) | 4750 (5.1) |  |
|  | Unknown/ot | 6435 (5.4) | 5260 (5.6) |  |
| Service region (%) | Central valley | 8649 (7.3) | 7807 (8.4) | 0.067 |
|  | East bay | 35095 (29.6) | 27085 (29.0) |  |
|  | North bay | 6182 (5.2) | 5285 (5.7) |  |
|  | North valley | 29735 (25.1) | 24496 (26.2) |  |
|  | Peninsula | 19465 (16.4) | 13669 (14.6) |  |
|  | South bay | 19283 (16.3) | 15059 (16.1) |  |
| Prior year flu (%) | 0 | 40829 (34.5) | 52505 (56.2) | 0.447 |
|  | 1 | 77580 (65.5) | 40896 (43.8) |  |
| Prior COVID-19 infection (%) | 0 | 99905 (84.4) | 77674 (83.2) | 0.033 |
|  | 1 | 18504 (15.6) | 15727 (16.8) |  |
| Charlson (%) | 0 | 86940 (73.4) | 67571 (72.3) | 0.025 |
|  | 1 | 16306 (13.8) | 13289 (14.2) |  |
|  | 2 | 6790 (5.7) | 5609 (6.0) |  |
|  | 3+ | 8373 (7.1) | 6932 (7.4) |  |

#### S2.2 Per protocol Results

**Table S11:** Pooled logistic results by week (STD)

| Time (weeks) | Risk Ratio (95% CI) | Risk Difference (95% CI) |
| --- | --- | --- |
| 1 | 0.69 (0.61, 0.75) | -0.0003 (-0.0004, -0.0002) |
| 2 | 0.69 (0.62, 0.75) | -0.0006 (-0.0008, -0.0005) |
| 3 | 0.70 (0.63, 0.75) | -0.0010 (-0.0013, -0.0008) |
| 4 | 0.70 (0.65, 0.75) | -0.0014 (-0.0018, -0.0012) |
| 5 | 0.71 (0.66, 0.76) | -0.0019 (-0.0023, -0.0015) |
| 6 | 0.72 (0.67, 0.76) | -0.0023 (-0.0028, -0.0019) |
| 7 | 0.73 (0.68, 0.77) | -0.0028 (-0.0033, -0.0023) |
| 8 | 0.74 (0.70, 0.78) | -0.0032 (-0.0038, -0.0027) |
| 9 | 0.75 (0.71, 0.79) | -0.0036 (-0.0043, -0.0030) |
| 10 | 0.77 (0.73, 0.80) | -0.0039 (-0.0046, -0.0032) |
| 11 | 0.78 (0.74, 0.82) | -0.0041 (-0.0050, -0.0033) |
| 12 | 0.80 (0.76, 0.83) | -0.0042 (-0.0051, -0.0035) |
| 13 | 0.81 (0.78, 0.84) | -0.0042 (-0.0052, -0.0034) |
| 14 | 0.83 (0.79, 0.86) | -0.0041 (-0.0051, -0.0032) |
| 15 | 0.84 (0.81, 0.88) | -0.0039 (-0.0050, -0.0030) |
| 16 | 0.86 (0.82, 0.89) | -0.0037 (-0.0048, -0.0028) |
| 17 | 0.88 (0.84, 0.91) | -0.0034 (-0.0046, -0.0025) |
| 18 | 0.89 (0.86, 0.92) | -0.0031 (-0.0042, -0.0022) |
| 19 | 0.90 (0.87, 0.94) | -0.0028 (-0.0039, -0.0019) |
| 20 | 0.92 (0.88, 0.95) | -0.0025 (-0.0036, -0.0015) |
| 21 | 0.93 (0.90, 0.96) | -0.0022 (-0.0033, -0.0011) |
| 22 | 0.94 (0.91, 0.97) | -0.0019 (-0.0030, -0.0008) |
| 23 | 0.95 (0.92, 0.99) | -0.0016 (-0.0027, -0.0005) |
| 24 | 0.96 (0.93, 1.00) | -0.0013 (-0.0024, -0.0001) |
| 25 | 0.97 (0.94, 1.01) | -0.0010 (-0.0021, 0.0002) |
| 26 | 0.98 (0.95, 1.01) | -0.0007 (-0.0019, 0.0005) |
| 27 | 0.99 (0.95, 1.02) | -0.0005 (-0.0016, 0.0008) |
| 28 | 0.99 (0.96, 1.03) | -0.0002 (-0.0014, 0.0011) |
| 29 | 1.00 (0.97, 1.04) | 0.0000 (-0.0012, 0.0013) |
| 30 | 1.01 (0.97, 1.05) | 0.0003 (-0.0010, 0.0016) |
| 31 | 1.01 (0.98, 1.05) | 0.0005 (-0.0008, 0.0018) |
| 32 | 1.02 (0.98, 1.06) | 0.0007 (-0.0006, 0.0021) |
| 33 | 1.03 (0.99, 1.07) | 0.0010 (-0.0004, 0.0023) |
| 34 | 1.03 (0.99, 1.07) | 0.0012 (-0.0002, 0.0026) |
| 35 | 1.04 (1.00, 1.08) | 0.0014 (0.0000, 0.0028) |
| 36 | 1.04 (1.01, 1.08) | 0.0017 (0.0003, 0.0031) |
| 37 | 1.05 (1.01, 1.09) | 0.0019 (0.0005, 0.0033) |
| 38 | 1.06 (1.02, 1.09) | 0.0022 (0.0008, 0.0036) |
| 39 | 1.06 (1.03, 1.10) | 0.0025 (0.0010, 0.0039) |
| 40 | 1.07 (1.03, 1.11) | 0.0027 (0.0013, 0.0042) |
| 41 | 1.08 (1.04, 1.11) | 0.0030 (0.0016, 0.0045) |
| 42 | 1.08 (1.04, 1.12) | 0.0033 (0.0019, 0.0049) |
| 43 | 1.09 (1.05, 1.13) | 0.0036 (0.0022, 0.0052) |
| 44 | 1.09 (1.06, 1.13) | 0.0040 (0.0026, 0.0055) |
| 45 | 1.10 (1.07, 1.14) | 0.0043 (0.0028, 0.0058) |
| 46 | 1.11 (1.07, 1.15) | 0.0047 (0.0031, 0.0061) |
| 47 | 1.12 (1.08, 1.15) | 0.0051 (0.0035, 0.0065) |
| 48 | 1.12 (1.09, 1.16) | 0.0055 (0.0039, 0.0070) |
| 49 | 1.13 (1.10, 1.17) | 0.0060 (0.0044, 0.0074) |
| 50 | 1.14 (1.11, 1.18) | 0.0065 (0.0050, 0.0080) |
| 51 | 1.15 (1.12, 1.19) | 0.0071 (0.0056, 0.0087) |
| 52 | 1.17 (1.13, 1.21) | 0.0077 (0.0062, 0.0093) |

**Table S12:** Pooled logistic results by week (EQC)

| Time (weeks) | Risk Ratio (95% CI) | Risk Difference (95% CI) |
| --- | --- | --- |
| 1 | 0.57 (0.50, 0.65) | -0.0005 (-0.0006, -0.0004) |
| 2 | 0.57 (0.51, 0.65) | -0.0010 (-0.0013, -0.0008) |
| 3 | 0.58 (0.53, 0.65) | -0.0016 (-0.0020, -0.0013) |
| 4 | 0.59 (0.54, 0.65) | -0.0023 (-0.0027, -0.0019) |
| 5 | 0.60 (0.55, 0.66) | -0.0030 (-0.0035, -0.0025) |
| 6 | 0.60 (0.56, 0.66) | -0.0037 (-0.0043, -0.0032) |
| 7 | 0.61 (0.57, 0.66) | -0.0044 (-0.0051, -0.0039) |
| 8 | 0.62 (0.59, 0.66) | -0.0052 (-0.0059, -0.0045) |
| 9 | 0.63 (0.60, 0.67) | -0.0058 (-0.0066, -0.0051) |
| 10 | 0.65 (0.61, 0.68) | -0.0064 (-0.0073, -0.0056) |
| 11 | 0.66 (0.62, 0.69) | -0.0070 (-0.0079, -0.0061) |
| 12 | 0.67 (0.64, 0.70) | -0.0074 (-0.0084, -0.0064) |
| 13 | 0.68 (0.65, 0.72) | -0.0077 (-0.0087, -0.0067) |
| 14 | 0.69 (0.66, 0.73) | -0.0079 (-0.0089, -0.0068) |
| 15 | 0.71 (0.68, 0.74) | -0.0080 (-0.0091, -0.0069) |
| 16 | 0.72 (0.69, 0.75) | -0.0080 (-0.0092, -0.0068) |
| 17 | 0.73 (0.70, 0.76) | -0.0080 (-0.0092, -0.0068) |
| 18 | 0.74 (0.71, 0.77) | -0.0080 (-0.0092, -0.0067) |
| 19 | 0.75 (0.72, 0.78) | -0.0079 (-0.0091, -0.0066) |
| 20 | 0.76 (0.73, 0.79) | -0.0078 (-0.0090, -0.0065) |
| 21 | 0.76 (0.73, 0.80) | -0.0077 (-0.0090, -0.0063) |
| 22 | 0.77 (0.74, 0.81) | -0.0076 (-0.0089, -0.0062) |
| 23 | 0.78 (0.75, 0.81) | -0.0075 (-0.0089, -0.0061) |
| 24 | 0.78 (0.75, 0.82) | -0.0074 (-0.0088, -0.0061) |
| 25 | 0.79 (0.76, 0.82) | -0.0073 (-0.0088, -0.0060) |
| 26 | 0.80 (0.76, 0.83) | -0.0073 (-0.0087, -0.0059) |
| 27 | 0.80 (0.77, 0.84) | -0.0072 (-0.0086, -0.0057) |
| 28 | 0.81 (0.77, 0.84) | -0.0071 (-0.0085, -0.0057) |
| 29 | 0.81 (0.78, 0.84) | -0.0070 (-0.0084, -0.0056) |
| 30 | 0.82 (0.78, 0.85) | -0.0069 (-0.0083, -0.0055) |
| 31 | 0.82 (0.79, 0.85) | -0.0068 (-0.0082, -0.0054) |
| 32 | 0.82 (0.79, 0.86) | -0.0067 (-0.0082, -0.0052) |
| 33 | 0.83 (0.80, 0.86) | -0.0067 (-0.0081, -0.0051) |
| 34 | 0.83 (0.80, 0.87) | -0.0066 (-0.0080, -0.0050) |
| 35 | 0.84 (0.80, 0.87) | -0.0065 (-0.0080, -0.0050) |
| 36 | 0.84 (0.81, 0.88) | -0.0064 (-0.0080, -0.0049) |
| 37 | 0.84 (0.81, 0.88) | -0.0063 (-0.0080, -0.0047) |
| 38 | 0.85 (0.81, 0.89) | -0.0062 (-0.0079, -0.0046) |
| 39 | 0.85 (0.82, 0.89) | -0.0061 (-0.0078, -0.0045) |
| 40 | 0.86 (0.82, 0.89) | -0.0061 (-0.0077, -0.0044) |
| 41 | 0.86 (0.82, 0.90) | -0.0060 (-0.0077, -0.0042) |
| 42 | 0.86 (0.83, 0.90) | -0.0059 (-0.0076, -0.0042) |
| 43 | 0.87 (0.83, 0.90) | -0.0058 (-0.0076, -0.0041) |
| 44 | 0.87 (0.83, 0.91) | -0.0057 (-0.0075, -0.0040) |
| 45 | 0.87 (0.84, 0.91) | -0.0057 (-0.0074, -0.0039) |
| 46 | 0.88 (0.84, 0.91) | -0.0056 (-0.0073, -0.0037) |
| 47 | 0.88 (0.85, 0.92) | -0.0054 (-0.0072, -0.0036) |
| 48 | 0.89 (0.85, 0.92) | -0.0053 (-0.0070, -0.0035) |
| 49 | 0.89 (0.86, 0.93) | -0.0051 (-0.0068, -0.0033) |
| 50 | 0.90 (0.87, 0.94) | -0.0048 (-0.0066, -0.0030) |
| 51 | 0.91 (0.87, 0.94) | -0.0045 (-0.0063, -0.0027) |
| 52 | 0.92 (0.88, 0.95) | -0.0041 (-0.0060, -0.0023) |

**Table S13:** Pooled logistic results by week (PCI)

| Time (weeks) | Risk Ratio (95% CI) | Risk Difference (95% CI) |
| --- | --- | --- |
| 1 | 0.62 (0.56, 0.71) | -0.0004 (-0.0005, -0.0003) |
| 2 | 0.62 (0.57, 0.71) | -0.0008 (-0.0009, -0.0006) |
| 3 | 0.63 (0.58, 0.72) | -0.0013 (-0.0015, -0.0009) |
| 4 | 0.64 (0.60, 0.72) | -0.0018 (-0.0020, -0.0013) |
| 5 | 0.65 (0.61, 0.72) | -0.0023 (-0.0026, -0.0018) |
| 6 | 0.66 (0.63, 0.72) | -0.0028 (-0.0031, -0.0022) |
| 7 | 0.67 (0.64, 0.73) | -0.0034 (-0.0037, -0.0026) |
| 8 | 0.68 (0.66, 0.74) | -0.0039 (-0.0043, -0.0030) |
| 9 | 0.69 (0.67, 0.75) | -0.0044 (-0.0048, -0.0034) |
| 10 | 0.70 (0.68, 0.76) | -0.0048 (-0.0052, -0.0037) |
| 11 | 0.72 (0.70, 0.77) | -0.0051 (-0.0056, -0.0039) |
| 12 | 0.73 (0.71, 0.79) | -0.0054 (-0.0058, -0.0040) |
| 13 | 0.74 (0.72, 0.80) | -0.0055 (-0.0060, -0.0041) |
| 14 | 0.76 (0.74, 0.81) | -0.0056 (-0.0061, -0.0041) |
| 15 | 0.77 (0.75, 0.83) | -0.0055 (-0.0061, -0.0039) |
| 16 | 0.78 (0.76, 0.84) | -0.0055 (-0.0061, -0.0038) |
| 17 | 0.79 (0.78, 0.86) | -0.0054 (-0.0059, -0.0037) |
| 18 | 0.81 (0.79, 0.87) | -0.0052 (-0.0057, -0.0034) |
| 19 | 0.82 (0.80, 0.88) | -0.0051 (-0.0056, -0.0032) |
| 20 | 0.83 (0.81, 0.89) | -0.0049 (-0.0054, -0.0030) |
| 21 | 0.84 (0.82, 0.90) | -0.0047 (-0.0053, -0.0028) |
| 22 | 0.84 (0.83, 0.91) | -0.0046 (-0.0052, -0.0026) |
| 23 | 0.85 (0.83, 0.92) | -0.0044 (-0.0050, -0.0024) |
| 24 | 0.86 (0.84, 0.92) | -0.0043 (-0.0049, -0.0022) |
| 25 | 0.87 (0.85, 0.93) | -0.0041 (-0.0047, -0.0021) |
| 26 | 0.87 (0.86, 0.94) | -0.0040 (-0.0046, -0.0018) |
| 27 | 0.88 (0.86, 0.95) | -0.0038 (-0.0044, -0.0016) |
| 28 | 0.88 (0.87, 0.95) | -0.0037 (-0.0043, -0.0015) |
| 29 | 0.89 (0.87, 0.96) | -0.0035 (-0.0041, -0.0012) |
| 30 | 0.90 (0.88, 0.97) | -0.0034 (-0.0040, -0.0010) |
| 31 | 0.90 (0.89, 0.97) | -0.0033 (-0.0038, -0.0009) |
| 32 | 0.91 (0.89, 0.98) | -0.0031 (-0.0037, -0.0007) |
| 33 | 0.91 (0.90, 0.98) | -0.0030 (-0.0035, -0.0005) |
| 34 | 0.92 (0.90, 0.99) | -0.0029 (-0.0033, -0.0003) |
| 35 | 0.92 (0.91, 1.00) | -0.0027 (-0.0032, -0.0002) |
| 36 | 0.93 (0.91, 1.00) | -0.0026 (-0.0030, 0.0000) |
| 37 | 0.93 (0.92, 1.01) | -0.0025 (-0.0029, 0.0002) |
| 38 | 0.93 (0.92, 1.01) | -0.0023 (-0.0028, 0.0004) |
| 39 | 0.94 (0.93, 1.02) | -0.0022 (-0.0027, 0.0005) |
| 40 | 0.94 (0.93, 1.02) | -0.0021 (-0.0025, 0.0007) |
| 41 | 0.95 (0.93, 1.02) | -0.0019 (-0.0024, 0.0009) |
| 42 | 0.95 (0.94, 1.03) | -0.0018 (-0.0023, 0.0011) |
| 43 | 0.96 (0.94, 1.03) | -0.0017 (-0.0022, 0.0013) |
| 44 | 0.96 (0.95, 1.04) | -0.0015 (-0.0021, 0.0015) |
| 45 | 0.97 (0.95, 1.04) | -0.0013 (-0.0019, 0.0016) |
| 46 | 0.97 (0.96, 1.05) | -0.0012 (-0.0017, 0.0018) |
| 47 | 0.98 (0.96, 1.05) | -0.0010 (-0.0016, 0.0021) |
| 48 | 0.98 (0.97, 1.06) | -0.0007 (-0.0013, 0.0024) |
| 49 | 0.99 (0.97, 1.07) | -0.0005 (-0.0010, 0.0028) |
| 50 | 1.00 (0.98, 1.08) | -0.0002 (-0.0007, 0.0031) |
| 51 | 1.01 (0.99, 1.09) | 0.0002 (-0.0004, 0.0035) |
| 52 | 1.02 (1.00, 1.10) | 0.0007 (-0.0000, 0.0040) |

**Figure S10:**
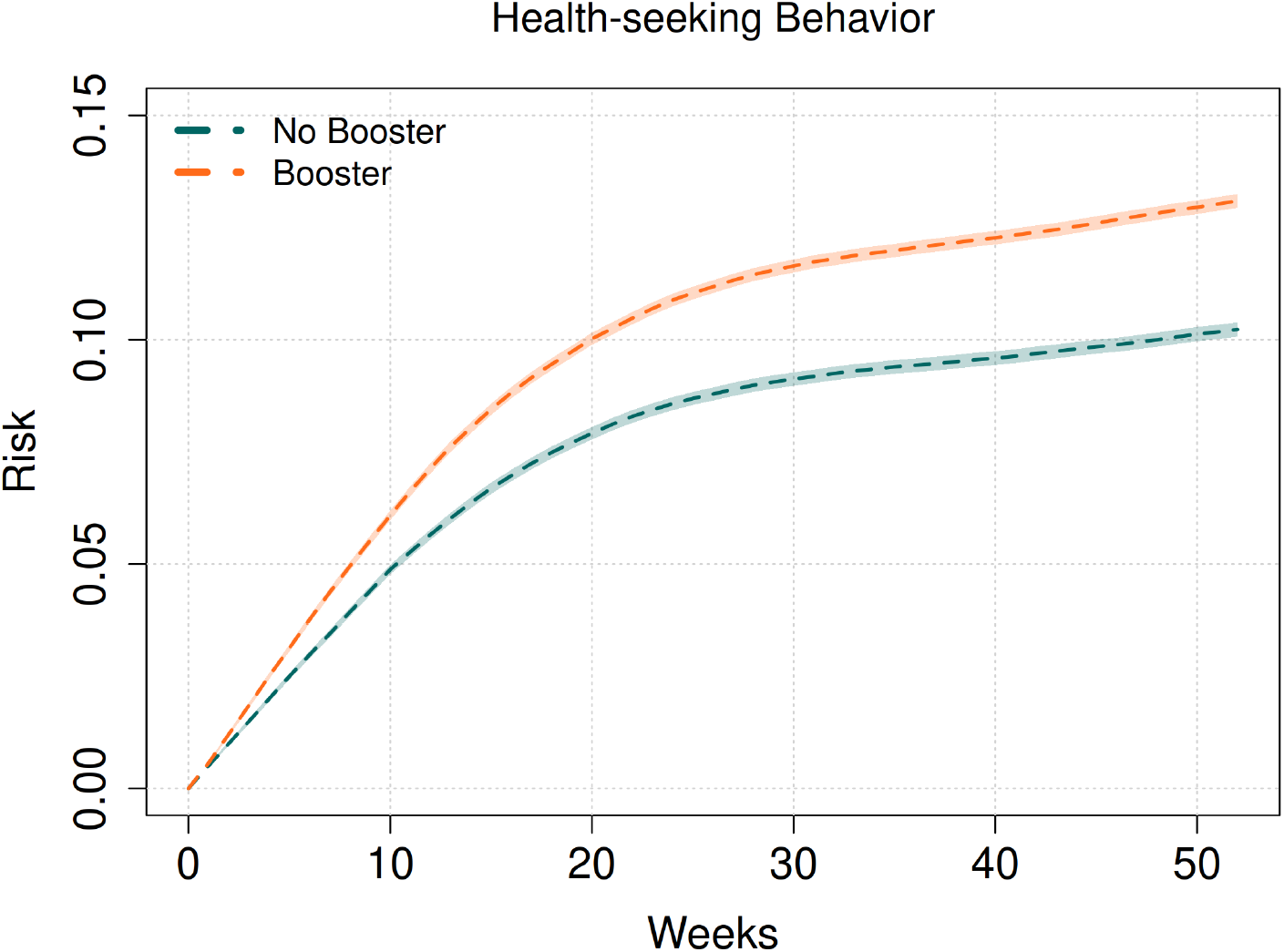
Cumulative incidence of testing negative by arm, per-protocol analysis. Testing negative serves as a negative control outcome in the EQC and PCI approaches. Cumulative incidence curves are expected to behave similarly in the absence of unmeasured confounding. Curves are Aalen–Johansen estimates and shaded bands are 95% confidence intervals

**Figure S11:**
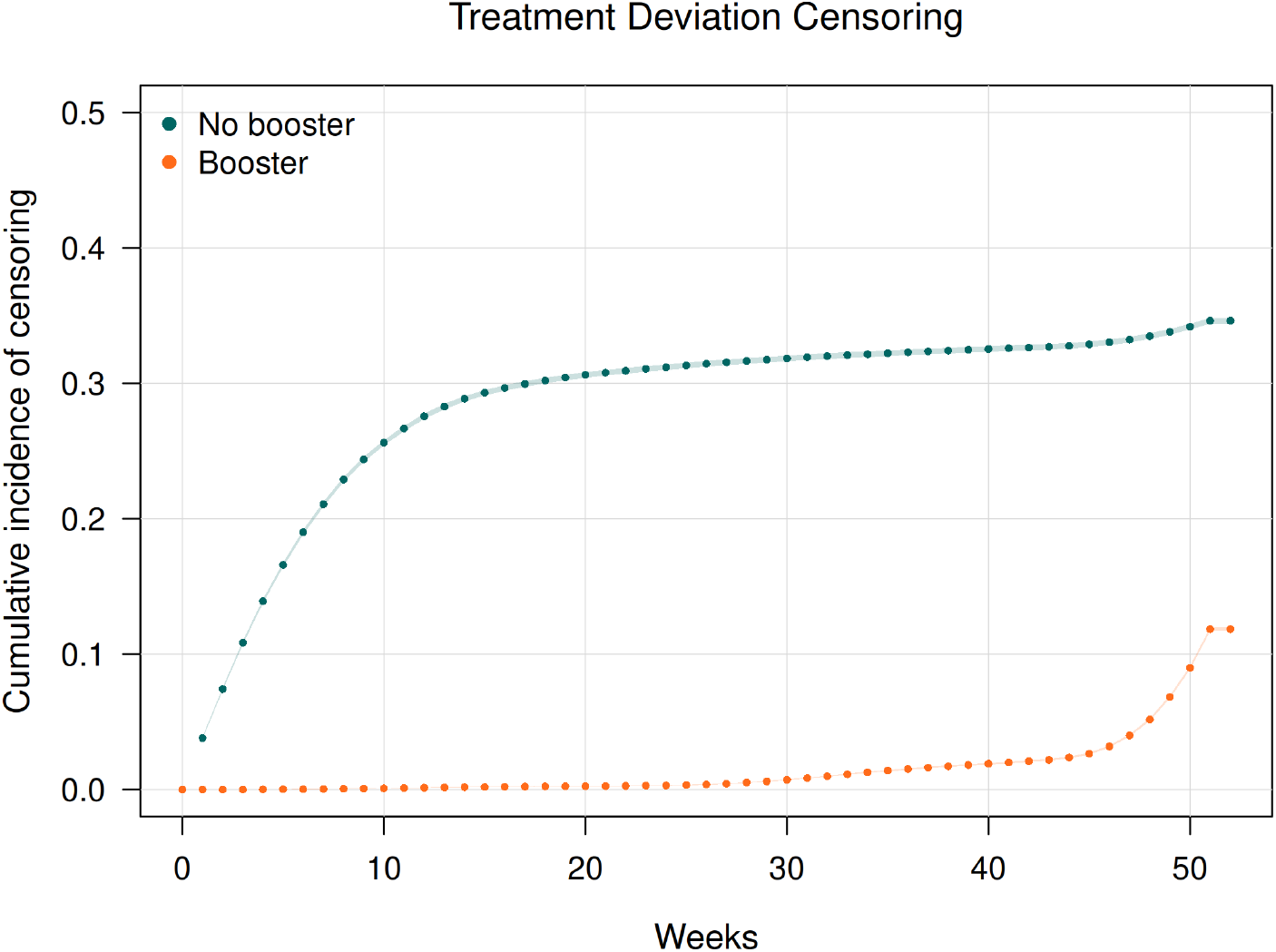
Cumulative incidence of treatment deviation censoring by arm, per-protocol analysis. Treatment deviation is defined as receipt of an additional booster vaccine dose after matching: a subsequent booster among the no-booster arm or a second booster among the booster arm. Points are Aalen–Johansen estimates and shaded bands are 95% confidence intervals

#### S2.3 Intention to Treat Results

**Table S14:** Cox results.

| Approach | Bivalent Booster |  | Prior year flu vaccine |  |
| --- | --- | --- | --- | --- |
|  | HR | VE (%) | HR | VE (%) |
| STD | 1.15 (1.11, 1.18) | -14.83 (-18.27, -11.50) | 1.25 (1.22, 1.29) | -25.44 (-29.47, -21.54) |
| TND | 0.93 (0.90, 0.97) | 6.54 (2.98, 9.96) | 1.03 (0.99, 1.07) | -2.53 (-6.70, 1.48) |
| EQC | 0.95 (0.92, 0.98) | 5.06 (1.72, 8.00) | 1.02 (0.99, 1.07) | -2.36 (-6.98, 1.26) |
| PCI | 0.93 (0.87, 0.98) | 6.97 (2.15, 12.75) | — | — |

**Table S15:** Pooled logistic results by week (STD)

| Time (weeks) | Risk Ratio (95% CI) | Risk Difference (95% CI) |
| --- | --- | --- |
| 1 | 0.69 (0.62, 0.76) | -0.0003 (-0.0004, -0.0002) |
| 2 | 0.70 (0.64, 0.76) | -0.0006 (-0.0008, -0.0005) |
| 3 | 0.71 (0.65, 0.77) | -0.0009 (-0.0012, -0.0007) |
| 4 | 0.73 (0.67, 0.78) | -0.0013 (-0.0016, -0.0010) |
| 5 | 0.74 (0.68, 0.79) | -0.0016 (-0.0020, -0.0013) |
| 6 | 0.76 (0.70, 0.80) | -0.0019 (-0.0024, -0.0016) |
| 7 | 0.77 (0.72, 0.81) | -0.0022 (-0.0028, -0.0018) |
| 8 | 0.79 (0.74, 0.82) | -0.0025 (-0.0031, -0.0020) |
| 9 | 0.80 (0.76, 0.84) | -0.0027 (-0.0033, -0.0022) |
| 10 | 0.82 (0.78, 0.85) | -0.0028 (-0.0035, -0.0023) |
| 11 | 0.83 (0.79, 0.87) | -0.0029 (-0.0037, -0.0023) |
| 12 | 0.85 (0.81, 0.88) | -0.0029 (-0.0037, -0.0022) |
| 13 | 0.87 (0.83, 0.90) | -0.0028 (-0.0037, -0.0021) |
| 14 | 0.88 (0.84, 0.91) | -0.0026 (-0.0036, -0.0019) |
| 15 | 0.90 (0.86, 0.93) | -0.0024 (-0.0034, -0.0016) |
| 16 | 0.91 (0.87, 0.95) | -0.0022 (-0.0032, -0.0013) |
| 17 | 0.92 (0.89, 0.96) | -0.0020 (-0.0030, -0.0010) |
| 18 | 0.94 (0.90, 0.97) | -0.0017 (-0.0027, -0.0007) |
| 19 | 0.95 (0.91, 0.99) | -0.0014 (-0.0024, -0.0004) |
| 20 | 0.96 (0.93, 1.00) | -0.0012 (-0.0022, -0.0001) |
| 21 | 0.97 (0.94, 1.01) | -0.0009 (-0.0019, 0.0001) |
| 22 | 0.98 (0.95, 1.01) | -0.0007 (-0.0016, 0.0004) |
| 23 | 0.99 (0.96, 1.02) | -0.0004 (-0.0014, 0.0007) |
| 24 | 0.99 (0.96, 1.03) | -0.0002 (-0.0012, 0.0010) |
| 25 | 1.00 (0.97, 1.04) | 0.0001 (-0.0009, 0.0012) |
| 26 | 1.01 (0.98, 1.05) | 0.0003 (-0.0007, 0.0014) |
| 27 | 1.01 (0.98, 1.05) | 0.0005 (-0.0005, 0.0017) |
| 28 | 1.02 (0.99, 1.06) | 0.0007 (-0.0004, 0.0019) |
| 29 | 1.03 (0.99, 1.06) | 0.0009 (-0.0002, 0.0021) |
| 30 | 1.03 (1.00, 1.07) | 0.0011 (-0.0000, 0.0024) |
| 31 | 1.04 (1.01, 1.08) | 0.0013 (0.0002, 0.0026) |
| 32 | 1.04 (1.01, 1.08) | 0.0015 (0.0004, 0.0028) |
| 33 | 1.05 (1.02, 1.09) | 0.0017 (0.0006, 0.0031) |
| 34 | 1.05 (1.02, 1.09) | 0.0020 (0.0008, 0.0033) |
| 35 | 1.06 (1.03, 1.10) | 0.0022 (0.0011, 0.0035) |
| 36 | 1.06 (1.03, 1.10) | 0.0024 (0.0013, 0.0037) |
| 37 | 1.07 (1.04, 1.11) | 0.0026 (0.0015, 0.0040) |
| 38 | 1.08 (1.05, 1.11) | 0.0029 (0.0017, 0.0042) |
| 39 | 1.08 (1.05, 1.12) | 0.0031 (0.0020, 0.0045) |
| 40 | 1.09 (1.06, 1.12) | 0.0034 (0.0022, 0.0048) |
| 41 | 1.09 (1.06, 1.13) | 0.0036 (0.0024, 0.0050) |
| 42 | 1.10 (1.07, 1.14) | 0.0039 (0.0027, 0.0053) |
| 43 | 1.10 (1.07, 1.14) | 0.0041 (0.0029, 0.0056) |
| 44 | 1.11 (1.08, 1.14) | 0.0044 (0.0032, 0.0059) |
| 45 | 1.11 (1.08, 1.15) | 0.0047 (0.0035, 0.0062) |
| 46 | 1.12 (1.09, 1.15) | 0.0050 (0.0038, 0.0065) |
| 47 | 1.12 (1.09, 1.16) | 0.0053 (0.0041, 0.0067) |
| 48 | 1.13 (1.10, 1.16) | 0.0056 (0.0044, 0.0070) |
| 49 | 1.13 (1.10, 1.17) | 0.0060 (0.0048, 0.0074) |
| 50 | 1.14 (1.11, 1.17) | 0.0063 (0.0051, 0.0078) |
| 51 | 1.15 (1.12, 1.18) | 0.0068 (0.0054, 0.0082) |
| 52 | 1.15 (1.12, 1.19) | 0.0072 (0.0059, 0.0086) |

**Table S16:**
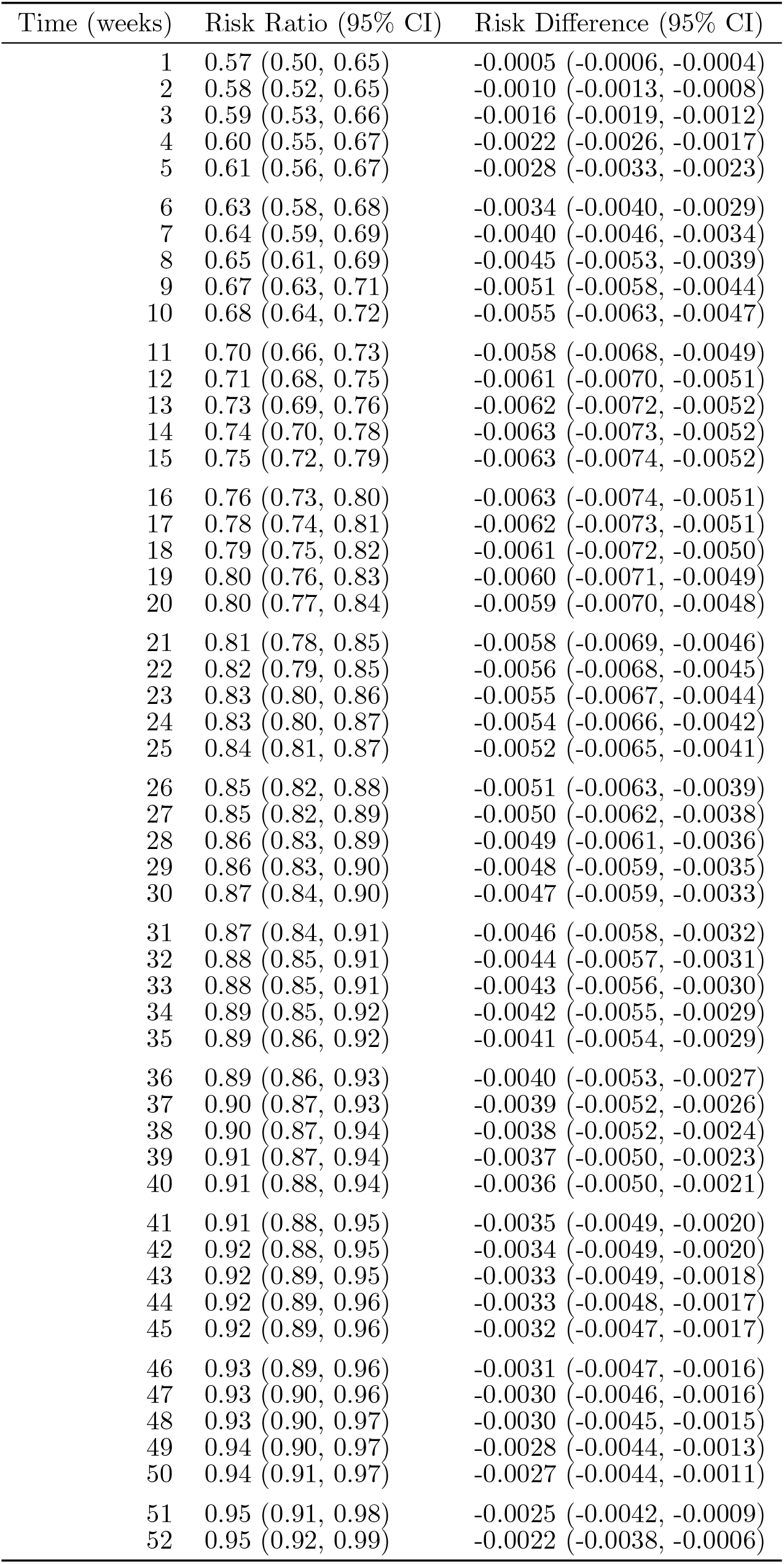
Pooled logistic results by week (EQC)

**Table S17:**
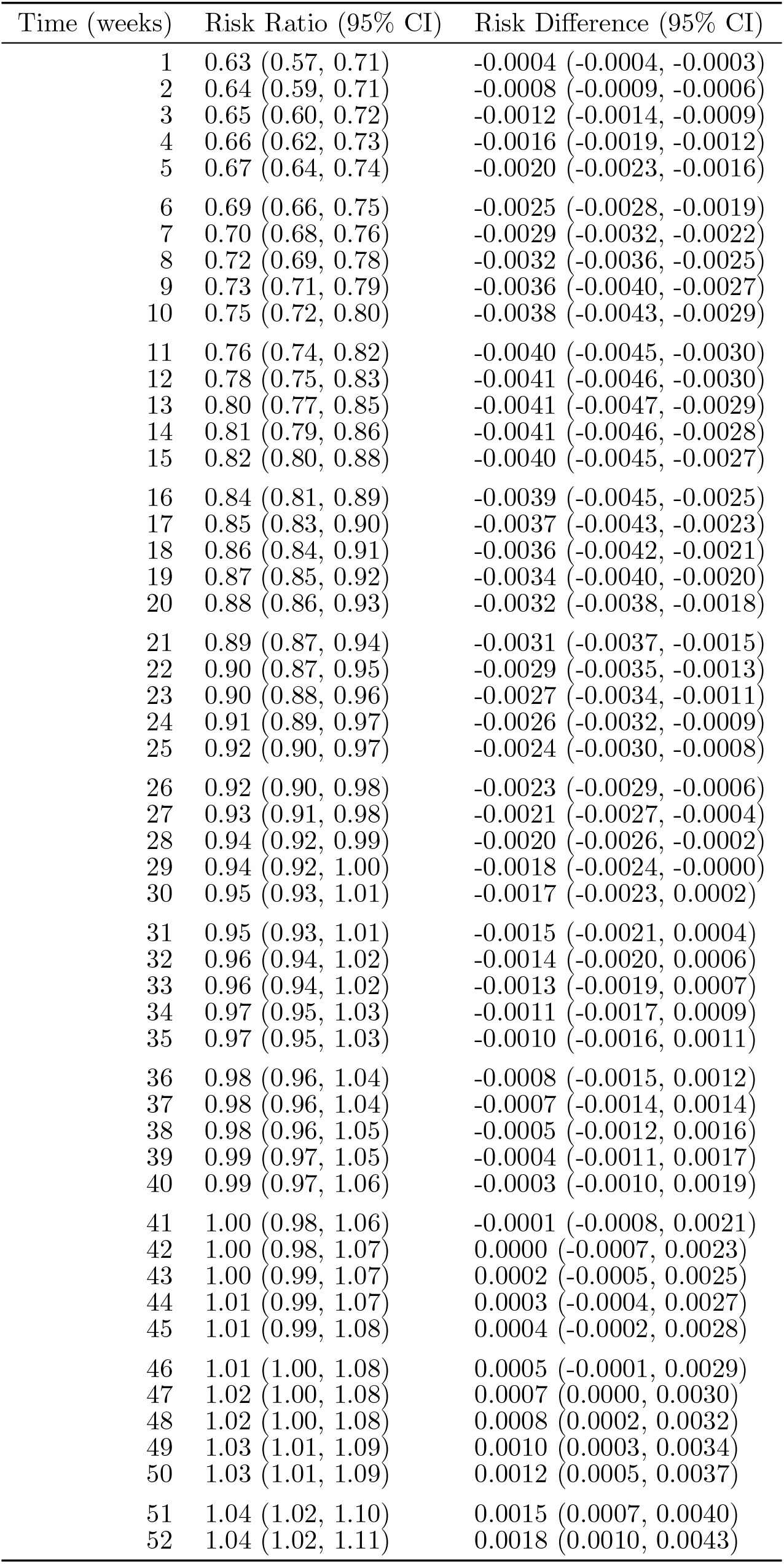
Pooled logistic results by week (PCI)

| Time (weeks) | Risk Ratio (95% CI) | Risk Difference (95% CI) |
| --- | --- | --- |
| 1 | 0.63 (0.57, 0.71) | -0.0004 (-0.0004, -0.0003) |
| 2 | 0.64 (0.59, 0.71) | -0.0008 (-0.0009, -0.0006) |
| 3 | 0.65 (0.60, 0.72) | -0.0012 (-0.0014, -0.0009) |
| 4 | 0.66 (0.62, 0.73) | -0.0016 (-0.0019, -0.0012) |
| 5 | 0.67 (0.64, 0.74) | -0.0020 (-0.0023, -0.0016) |
| 6 | 0.69 (0.66, 0.75) | -0.0025 (-0.0028, -0.0019) |
| 7 | 0.70 (0.68, 0.76) | -0.0029 (-0.0032, -0.0022) |
| 8 | 0.72 (0.69, 0.78) | -0.0032 (-0.0036, -0.0025) |
| 9 | 0.73 (0.71, 0.79) | -0.0036 (-0.0040, -0.0027) |
| 10 | 0.75 (0.72, 0.80) | -0.0038 (-0.0043, -0.0029) |
| 11 | 0.76 (0.74, 0.82) | -0.0040 (-0.0045, -0.0030) |
| 12 | 0.78 (0.75, 0.83) | -0.0041 (-0.0046, -0.0030) |
| 13 | 0.80 (0.77, 0.85) | -0.0041 (-0.0047, -0.0029) |
| 14 | 0.81 (0.79, 0.86) | -0.0041 (-0.0046, -0.0028) |
| 15 | 0.82 (0.80, 0.88) | -0.0040 (-0.0045, -0.0027) |
| 16 | 0.84 (0.81, 0.89) | -0.0039 (-0.0045, -0.0025) |
| 17 | 0.85 (0.83, 0.90) | -0.0037 (-0.0043, -0.0023) |
| 18 | 0.86 (0.84, 0.91) | -0.0036 (-0.0042, -0.0021) |
| 19 | 0.87 (0.85, 0.92) | -0.0034 (-0.0040, -0.0020) |
| 20 | 0.88 (0.86, 0.93) | -0.0032 (-0.0038, -0.0018) |
| 21 | 0.89 (0.87, 0.94) | -0.0031 (-0.0037, -0.0015) |
| 22 | 0.90 (0.87, 0.95) | -0.0029 (-0.0035, -0.0013) |
| 23 | 0.90 (0.88, 0.96) | -0.0027 (-0.0034, -0.0011) |
| 24 | 0.91 (0.89, 0.97) | -0.0026 (-0.0032, -0.0009) |
| 25 | 0.92 (0.90, 0.97) | -0.0024 (-0.0030, -0.0008) |
| 26 | 0.92 (0.90, 0.98) | -0.0023 (-0.0029, -0.0006) |
| 27 | 0.93 (0.91, 0.98) | -0.0021 (-0.0027, -0.0004) |
| 28 | 0.94 (0.92, 0.99) | -0.0020 (-0.0026, -0.0002) |
| 29 | 0.94 (0.92, 1.00) | -0.0018 (-0.0024, -0.0000) |
| 30 | 0.95 (0.93, 1.01) | -0.0017 (-0.0023, 0.0002) |
| 31 | 0.95 (0.93, 1.01) | -0.0015 (-0.0021, 0.0004) |
| 32 | 0.96 (0.94, 1.02) | -0.0014 (-0.0020, 0.0006) |
| 33 | 0.96 (0.94, 1.02) | -0.0013 (-0.0019, 0.0007) |
| 34 | 0.97 (0.95, 1.03) | -0.0011 (-0.0017, 0.0009) |
| 35 | 0.97 (0.95, 1.03) | -0.0010 (-0.0016, 0.0011) |
| 36 | 0.98 (0.96, 1.04) | -0.0008 (-0.0015, 0.0012) |
| 37 | 0.98 (0.96, 1.04) | -0.0007 (-0.0014, 0.0014) |
| 38 | 0.98 (0.96, 1.05) | -0.0005 (-0.0012, 0.0016) |
| 39 | 0.99 (0.97, 1.05) | -0.0004 (-0.0011, 0.0017) |
| 40 | 0.99 (0.97, 1.06) | -0.0003 (-0.0010, 0.0019) |
| 41 | 1.00 (0.98, 1.06) | -0.0001 (-0.0008, 0.0021) |
| 42 | 1.00 (0.98, 1.07) | 0.0000 (-0.0007, 0.0023) |
| 43 | 1.00 (0.99, 1.07) | 0.0002 (-0.0005, 0.0025) |
| 44 | 1.01 (0.99, 1.07) | 0.0003 (-0.0004, 0.0027) |
| 45 | 1.01 (0.99, 1.08) | 0.0004 (-0.0002, 0.0028) |
| 46 | 1.01 (1.00, 1.08) | 0.0005 (-0.0001, 0.0029) |
| 47 | 1.02 (1.00, 1.08) | 0.0007 (0.0000, 0.0030) |
| 48 | 1.02 (1.00, 1.08) | 0.0008 (0.0002, 0.0032) |
| 49 | 1.03 (1.01, 1.09) | 0.0010 (0.0003, 0.0034) |
| 50 | 1.03 (1.01, 1.09) | 0.0012 (0.0005, 0.0037) |
| 51 | 1.04 (1.02, 1.10) | 0.0015 (0.0007, 0.0040) |
| 52 | 1.04 (1.02, 1.11) | 0.0018 (0.0010, 0.0043) |

## Footnotes

1 Trivially, 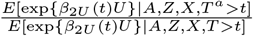 is exactly one under the null as *T* ^*a*^ = *T* for every individual.

2 To illustrate the difference between the scale on which equi-confounding is assumed and how the hazards themselves are measured: We could alternatively continue to assume equi-confounding on multiplicative scale but choose to model the individuals hazards *λ*_1_(*t* | *A, Z, X*) and *λ*_2_(*t* | *A, Z, X*) using an additive model specification via Aalen’s additive hazard model.

3 Li, Linderman, Shi, and Tchetgen [3] show that, under additive models, the location-shift model need only be assumed for *U*| *A, Z, X*, i.e. it does not need to persist across risk sets. However, we retain the stronger assumption here for symmetry with the prior section.

